# The impact of B_1_ ^+^ inhomogeneity on image quality metrics and morphometric statistical inferences at 7 T MRI

**DOI:** 10.64898/2026.06.08.26355136

**Authors:** Kai Liu, Kȃmil Uludaǧ, Irenaeus F.M. de Coo, Hubert J.M. Smeets, Jacobus F.A. Jansen, Elia Formisano, Benedikt A. Poser, Roy A.M. Haast, Dimo Ivanov

## Abstract

**Introduction:** Structural neuroimaging relies on T1-weighted (T1w) magnetic resonance imaging (MRI) for brain morphometry, yet at 7 Tesla (7 T) transmit field (B_1_ ^+^) inhomogeneity remains a major source of bias. Although Magnetization Prepared 2 Rapid Acquisition Gradient Echoes (MP2RAGE) improves the tissue contrast, residual B_1_ ^+^ effects may persist and may be exacerbated in aging or clinical populations, where anatomical and physiological factors further challenge image quality and preprocessing. The impact of B_1_ ^+^ inhomogeneity on automated quality assessment and morphometric statistical inference remains insufficiently understood.

**Methods:** Submillimeter 7 T MP2RAGE brain acquisitions from carriers of a mitochondrial gene mutation (m.3243A>G) and controls were retrieved from previous studies. Image quality before and after B_1_ ^+^ inhomogeneity correction was assessed by multiple automated pipelines. Case-control morphometric studies, including regional volume and mean cortical thickness, were analyzed in both registration-based and deep learning-based segmentation frameworks. Changes in image quality metrics (IQMs) and morphometric statistical significance were evaluated to determine the impact of B_1_ ^+^ inhomogeneity correction.

**Results:** Overall image quality rating and metrics sensitive to intensity non-uniformity and topological integrity consistently improved after B_1_ ^+^ inhomogeneity correction. However, its impact on morphometric statistical inferences was strongly method-dependent. Some pipelines showed redistribution of significant regions, whereas others predominantly demonstrated increased effects in sensitivity. Across methods, B_1_ ^+^ inhomogeneity correction altered the findings of morphometric analyses, particularly in cortical regions.

**Conclusion:** Residual B_1_ ^+^ inhomogeneity at 7 T substantially influences both image quality control and morphometric evaluations. Current automated quality control approaches can hardly capture these effects reliably. B_1_ ^+^ inhomogeneity correction will not only improve intensity uniformity, but also change sensitivity of morphometric statistical inferences. To establish reliable morphometric biomarkers at UHF strengths, explicit B_1_ ^+^ inhomogeneity correction and carefully chosen preprocessing strategies are practically necessary and highly recommended.

## 1. INTRODUCTION

Structural magnetic resonance imaging (MRI) is a key modality in modern neuroimaging protocols, enabling non-invasive characterization of brain anatomy and supporting a wide range of neuroscientific and clinical research applications (Sabuncu et al., 2015). In particular, T1-weighted (T1w) imaging has become the standard for assessing brain morphology due to its ability to provide strong contrast between cerebrospinal fluid (CSF), gray matter (GM), and white matter (WM), driven primarily by differences in longitudinal relaxation time (T1) (Nitz & Reimer, 1999). Over decades, sequence development, most notably the Magnetization Prepared Rapid Gradient Echo (MPRAGE), has optimized this contrast and became the clinical standard for three dimensional (3D) isotropic T1w imaging (Mugler & Brookeman, 1990).

Despite its widespread use, conventional T1w imaging at lower field strengths remains limited by relatively low spatial resolution and signal-to-noise ratio (SNR), constraining the detection of subtle anatomical variations (Opheim et al., 2021). The advent of ultra-high field (UHF) MRI, particularly 7 Tesla (7 T), has addressed these limitations by providing increased SNR and submillimeter resolution within reasonable scanning time, thereby enabling more precise characterization of cortical and subcortical structures (Van Der Zwaag et al., 2016). However, these benefits come at the cost of higher sensitivity to inhomogeneity of static magnetic (B_0_) and radiofrequency (RF) transmit and receive fields (Ibrahim et al., 2001; Kangarlu et al., 1999; Van de Moortele et al., 2005). Most notably, transmit field (B_1_^+^) inhomogeneities lead to spatially varying flip angles, resulting in non-uniform image intensity and contrast at 7 T (Collins et al., 2005). These effects are further exacerbated by tissue-dependent dielectric properties and anatomical factors, such as ventricular geometry, which introduce complex interference patterns (Brink et al., 2014; Sled & Pike, 1998). While receive field (B_1_^-^), spin density (M_0_) and “effective” transverse relaxation time (T2*) biases were reduced in specifically developed sequences such as the Magnetization Prepared 2 Rapid Acquisition Gradient Echoes (MP2RAGE), residual B_1_^+^ inhomogeneity effects may persist (Marques et al., 2010; Opheim et al., 2021; Van de Moortele et al., 2009). Post hoc correction with an additionally acquired B_1_^+^ map has therefore been proposed (Marques & Gruetter, 2013).

The impact of B_1_^+^ inhomogeneity on quantitative MRI measures is well established. Previous studies have demonstrated significant effects on both relaxometry and morphometry metrics, including biased cortical thickness and subcortical volume measurements, even in young healthy populations (Haast et al., 2018b, 2021). Consequently, failure to account for B_1_^+^ inhomogeneity may introduce systematic errors and confound downstream analyses. Despite this, the B_1_^+^ inhomogeneity correction is not consistently applied (Droby et al., 2021; Maynord et al., 2026).

Quality control (QC) is a critical step in neuroimaging pipelines, traditionally relying on expert visual inspection (Monereo-Sánchez et al., 2021), which is subjective and difficult to scale (Woodard & Carley-Spencer, 2006). Automated approaches, such as Magnetic Resonance Image Quality Control’s (MRIQC) binary classification (Esteban, 2017), Computational Anatomy Toolbox (CAT) structural image quality ratings (SIQR) (Dahnke et al., 2025), topological image quality proxies (Rosen et al., 2018), as well as post-cortical mesh reconstruction quality assessment, have emerged as scalable alternatives. While the influence of B_1_^+^ inhomogeneity on quantitative and morphometric measures is increasingly recognized, its effect on image quality metrics (IQMs) remains largely unexplored, which may restrict the application of these automated QC approaches on 7 T MP2RAGE data.

Moreover, sensitivity to B_1_ ^+^ inhomogeneities might become even more pronounced in clinical populations. Factors, such as increased head motion (Moqadam et al., 2024), ventricular enlargement (MacLennan et al., 2022), and pathological lesions like inflammation and tumors which alter focal tissue components (Brett et al., 2001) can further complicate preprocessing (Gilmore et al., 2021). It remains unclear whether B_1_ ^+^ inhomogeneity, and its correction, systematically influences morphometric statistical inferences in case-control studies.

In parallel, advances in neuroimaging analysis have shifted from traditional registration-based segmentation frameworks (Ashburner & Friston, 2005; Cohen et al., 1992; Kennedy et al., 1989; Klauschen et al., 2009; Kondrateva et al., 2025; Zhang et al., 2001) toward deep learning-based ones (Billot, Greve, et al., 2023; Billot, Magdamo, et al., 2023; Fortin et al., 2026; Henschel et al., 2022), offering faster processing speed and improved robustness. Nevertheless, their resilience to UHF-specific artifacts, including B_1_ ^+^ inhomogeneity, has not been fully established, raising the question of whether correction strategies remain critical in deep learning-based segmentation frameworks.

To address the aforementioned knowledge gaps, the present study investigates the impact of B_1_ ^+^ inhomogeneity correction on both automated QC approaches and morphometric analyses with 7 T MP2RAGE acquisition from both gene mutation carriers and healthy controls. By extending evaluation beyond healthy cohorts, this work aims to clarify the broader implications of B_1_ ^+^ inhomogeneity for neuroimaging research at UHF strengths.

## 2. METHODS

### 2.1. Participant recruitment

Participants were recruited for a m.3243A>G mitochondrial gene mutation cohort. To minimize confounding by severe pathology, gene mutation carriers with acute or severe phenotypes were excluded (Mickelsson et al., 2025, 2026). Identical to our previously published studies (Haast et al., 2018a, 2022), 22 carriers without detectable abnormalities on conventional 1.5 and 3 T MRI, as well as 15 controls matched for age and sex were included in our final sample (Supplementary Material 1). All participants provided written informed consent in accordance with the Declaration of Helsinki, and the study protocol was approved by the Medical Ethics Review Committee of the Maastricht University Medical Center.

### 2.2. MRI acquisition

All imaging was performed on a 7 T whole-body MRI scanner (Siemens Healthineers, Erlangen, Germany) using a 32-channel phased-array head coil (Nova Medical, Wilmington, MA, USA). Dielectric pads containing a 25% suspension of barium titanate in deuterated water were placed near the temporal regions to improve B_1_ ^+^ homogeneity locally (Teeuwisse et al., 2012). B_1_ ^+^ maps were obtained using the Saturation-prepared with 2 rapid gradient echoes (SA2RAGE) sequence (Eggenschwiler et al., 2012), high resolution structural images (T1w) were acquired using the MP2RAGE sequence (Marques et al. 2010), the “uniform” reconstruction was used to preserve the original signal properties, at the expense of residual “salt & pepper” air background noise (Supplementary Material 2).

### 2.3. B_1_^+^ inhomogeneity correction

All preprocessing (Figure 1) was performed on a dedicated workstation (Supplementary Material 3). Digital Imaging and Communications in Medicine (DICOM) data were converted to Neuroimaging Informatics Technology Initiative (NIfTI) format using *dcm2niix* (Rorden, 2025). SA2RAGE were rigidly registered to the MP2RAGE data using FreeSurfer’s *mri_robust_register* (Reuter et al. 2010). B_1_ ^+^ correction of MP2RAGE “uniform” (UNI) images was then applied using established MATLAB-based methods (Marques and Gruetter 2013) (Figure 1A). Gradient nonlinearity was corrected using the *mri_gradunwarp* tool with calibration parameters supplied by Siemens (Jovicich et al., 2006).

**Figure 1.**
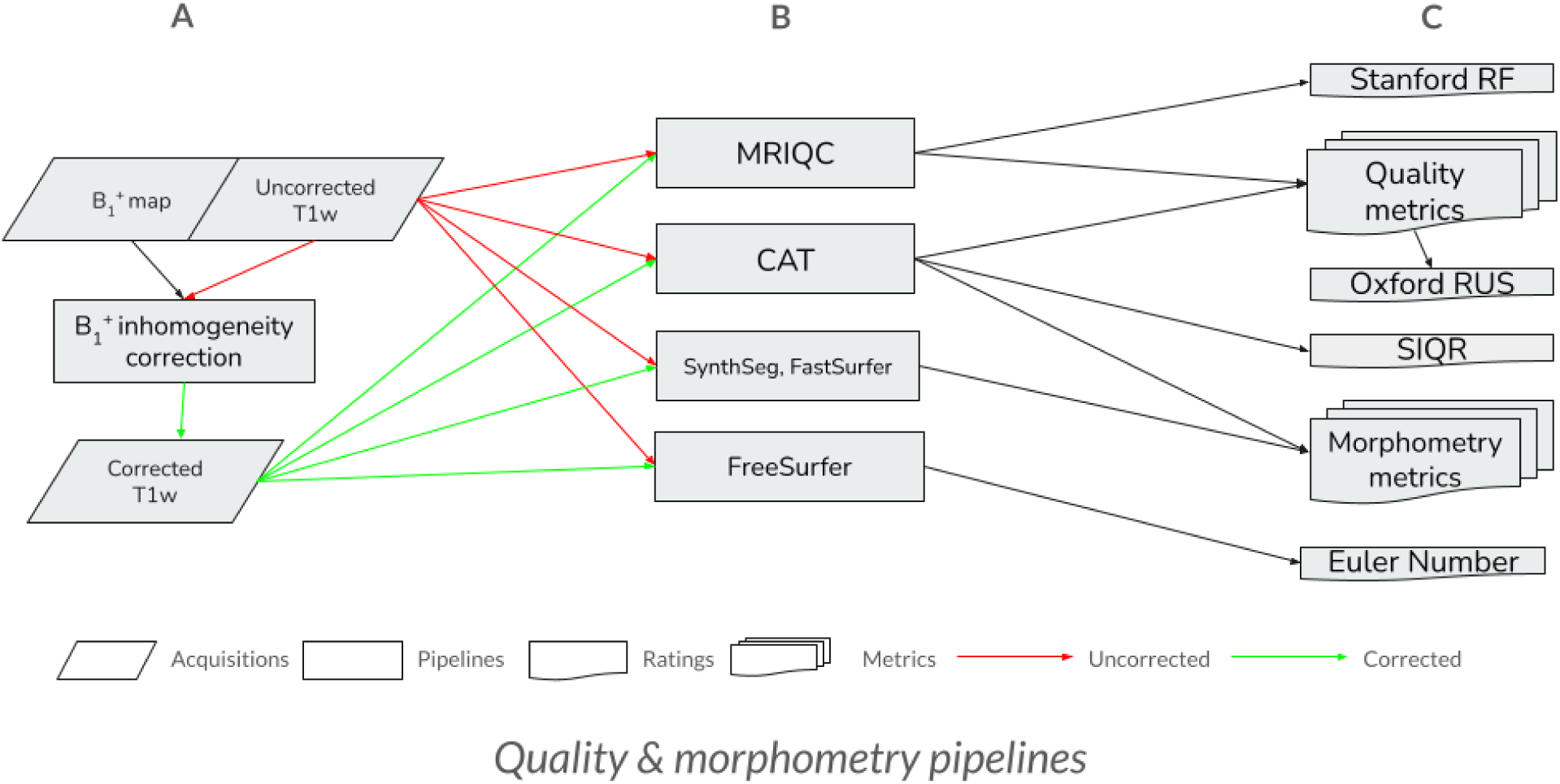
Schematic overview of the analysis pipelines used in this study. Image quality of preprocessed MP2RAGE data (A) was assessed using MRIQC, CAT, and FreeSurfer-derived Euler numbers. Morphometric analyses (B-C) were performed using registration-based (CAT) and deep learning-based (SynthSeg and FastSurfer) segmentation frameworks. FreeSurfer was only used for Euler number extraction, while FreeSurfer-compatible volumetric and cortical thickness measures were obtained from SynthSeg and FastSurfer.

### 2.4. Automated QC approaches

#### MRIQC (“Stanford RF”) classifier

MP2RAGE UNI images both before and after B_1_^+^ inhomogeneity correction were organized according to the Brain Imaging Data Structure (BIDS) (Gorgolewski et al., 2016) and processed using MRIQC (v0.15.1) in an Apptainer environment. The pipeline heavily depends on Advanced Normalization Tools Ecosystem (ANTsX) bias field correction (Sled et al. 1998; Tustison et al. 2010), Analysis of Functional NeuroImages (AFNI) skull stripping (Cox, 1996) and Functional Magnetic Resonance Imaging of the Brain Automated Segmentation Tool (FAST) segmentation (Zhang et al., 2001). MRIQC computes a broad set of IQMs, including protocol-derived parameters, statistical summaries, and artifact-related measures (e.g., noise, ghosting and bias, see also Supplementary Table 4). At the group level, 68 IQMs were aggregated and evaluated using a pretrained random forest (RF) classifier (*mriqc_clf*), here referred to as “Stanford RF” (Esteban, 2017).

#### CAT rating

MP2RAGE UNI images both before and after B_1_ ^+^ inhomogeneity correction were manually reoriented along the anterior commissure - posterior commissure (AC-PC) line in the Statistical Parametric Mapping (SPM) framework (Ashburner, 2009). Then complementary preprocessing was performed using SPM’s unified segmentation module and the CAT toolbox (Ashburner & Friston, 2005; Gaser et al., 2024), which jointly performs bias correction, skull stripping, tissue segmentation and cortical mesh reconstruction. Customized pipeline options were selected for our submillimeter UHF acquisition (i.e., *heavy* for “bias correction”, *native* for “resolution”). Similar to MRIQC, CAT computes component IQMs on noise, ghosting and bias, then composites them into a single SIQR (Appendix, Supplementary Material 4). Additionally, we tabulated 36 CAT IQMs for use in another binary classifier (“Oxford RUS”).

#### “Oxford RUS” classifier

To further assess image quality, MRIQC and CAT metrics were combined and evaluated using a pretrained robust undersampling (RUS) classifier. This model incorporates features from both pipelines, with CAT-derived noise metrics contributing prominently (Bhalerao et al., 2025). Predictions from this classifier are referred to as “Oxford RUS”.

#### FreeSurfer-derived topological proxy of image quality

Unlike CAT, which can directly process raw MP2RAGE UNI images, FreeSurfer and FastSurfer require prior removal of the characteristic MP2RAGE background noise. Therefore, images acquired before and after B_1_ ^+^ inhomogeneity correction were first converted to MPRAGE-like images using the AFNI-based *3dMPRAGEise* script (Kashyap, 2021). Hemispheric Euler numbers were subsequently extracted using FreeSurfer (v6.0.0) configured for submillimeter-resolution data (Fischl, 2012; Monereo-Sánchez et al., 2021; Rosen et al., 2018). To isolate topology estimates from downstream corrections, the reconstruction pipeline was intentionally terminated prior to automatic topology fixing. The arithmetic mean of the left and right hemispheric Euler numbers (AMHEN) was then calculated and used as a topology-based proxy of image quality:

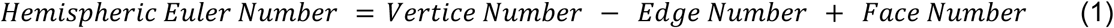

#### FastSurfer-derived post-reconstruction quality assessment

In parallel, cortical surface reconstruction was performed on the same MPRAGE-like images using FastSurfer (v2.4.2), a deep learning-based alternative to FreeSurfer (Henschel et al., 2020). Post-reconstruction quality assessment was performed using the *fsqc*, which combines conventional image quality metrics with shape- and symmetry-based measures derived from *LaPy* and *BrainPrint* (Reuter et al., 2006; Wachinger et al., 2015). Taking a normative model as reference, the number of structures exceeding the upper and lower bounds across these measurements was summarized as the Normative Outlier Number (NON), with higher values indicating greater deviation from the expected anatomical profile.

### 2.5. Morphometric analyses

#### Registration-based segmentation frameworks

Registration-based morphometric analyses were performed using CAT. From the surface pipeline, mean thickness was extracted for parcellations defined by the Desikan-Killiany (DK40) and Destrieux atlases (Desikan et al., 2006; Destrieux et al., 2010). From the volumetric pipeline, regional gray matter volumes were extracted using the Automated Anatomical Labeling atlas 3 (AAL3), together with total intracranial volume (TIV) estimates (Rolls et al., 2020).

#### Deep learning-based segmentation frameworks

To evaluate the robustness of deep learning approaches to residual B_1_ ^+^ inhomogeneity, morphometric analyses were additionally performed using FastSurferVINN, CerebNet, and SynthSeg. FastSurferVINN employs a voxel-size independent neural network architecture that preserves the advantages of submillimeter-resolution imaging by directly incorporating voxel size into the segmentation process (Henschel et al., 2022). Cortical thickness and GM volumes were extracted for the DK40 and Destrieux atlases from the FastSurfer surface pipeline.

Because some uncorrected acquisitions could not be fully processed by FastSurfer’s CerebNet (Faber et al., 2022), whole-brain segmentation and cortical parcellation were also performed using SynthSeg, a synthetic-data-trained convolutional neural network designed to be robust across image contrasts and resolutions (Billot, Greve, et al., 2023). Cerebellar parcellations were obtained using CerebNet, while SynthSeg provided whole-brain regional volumes and TIV estimates. SynthSeg-derived brainstem segmentations were further parcellated into medulla oblongata, pons, midbrain and superior cerebellar peduncle using the FreeSurfer *segment_subregions_brainstem* pipeline (Iglesias et al., 2015).

Notably, surface-based frameworks (FastSurfer) and voxel-based segmentation frameworks (CAT and SynthSeg) derive regional volumes using fundamentally different approaches (Winkler et al., 2010). Including both classes of methods enabled a comprehensive assessment of how B_1_ ^+^ inhomogeneity correction influences morphometric outcomes across analytical frameworks.

### 2.6. Statistical analyses & data visualization

Statistical analyses were conducted using jamovi (v2.7.30). The Wilcoxon signed rank test was used for composite SIQR percentage and NON count before and after B_1_ ^+^ inhomogeneity correction. The linear mixed model (LMM) was constructed in the module GAMLj3 (v3.6.5), to test component bias inhomogeneity metrics and AMHEN differences before and after B_1_ ^+^ inhomogeneity correction, taking participants’ group (carrier vs. control), sex and age as factors or covariate.

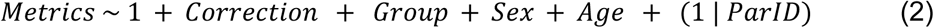

Considering our sample size, no random effect interactions between factors or covariates were explored, type III sum and restricted maximum likelihood were used.

Group difference (carriers < controls) in ROIs volume before and after B_1_^+^ inhomogeneity correction was assessed using permutation-based inference (Permutation Analysis of Linear Models, PALM) (Winkler, Webster, et al., 2016), with age, sex, and TIV included as nuisance variables. Group difference (carriers < controls) in regional mean cortical thickness (in DK40 and Destrieux parcellations) was assessed in the same way without TIV as a nuisance. To ensure accurate p-values for the large number of ROIs tested, Generalized Pareto Distribution (GPD) tail approximation (*-accel tail*) was applied (Winkler, Ridgway, et al., 2016). Statistical significance was determined using 5,000 permutations and corrected with the false discovery rate (FDR) method (Benjamini & Hochberg, 1995), which was done once for each atlas except CerebNet and *segment_subregions brainstem,* both pooled with SynthSeg and used the same TIV. Packages in Python (*xml, numpy, pandas, matplotlib, nibabel, PIL*) and R (*ggtext, ggplot2, ggseg, ggsegAAL, ggsegCerebellum, ggsegFreeSurfer, patchwork*) were utilized in data visualization (Mowinckel & Vidal-Piñeiro, 2020).

## 3. RESULTS

### 3.1. Automated QC approaches

#### Binary classifiers, SIQR and NON

The MP2RAGE UNI image from a carrier before B_1_^+^ inhomogeneity correction failed to complete MRIQC processing (Figure 2, Supplementary Material 5). Among the remaining datasets, the “Stanford RF” classifier labeled nearly all as “low” image quality, both before and after B_1_^+^ inhomogeneity correction. In contrast, the “Oxford RUS” classifier consistently labeled all as “high” image quality, irrespective of correction status. CAT-derived SIQR grades for uncorrected images ranged from C to A-, with all corrected images showing either improvement or no change in grade (Figure 3a). Quantitatively, SIQR percentages increased significantly following B_1_^+^ inhomogeneity correction in both carriers (Wilcoxon *W* = 14.0, *p* < 0.001) and controls (Wilcoxon *W* = 15.0, *p* = 0.008), indicating improved overall image quality. NON did not change in either carriers (Wilcoxon *W* = 39.5, *p* = 0.696) or controls (Wilcoxon *W* = 25.0, *p* = 0.810), showing the FastSurfer reconstruction pipeline was not sensitive to changes in image quality before and after B_1_ ^+^ inhomogeneity correction, either (Figure 3b).

**Figure 2.**
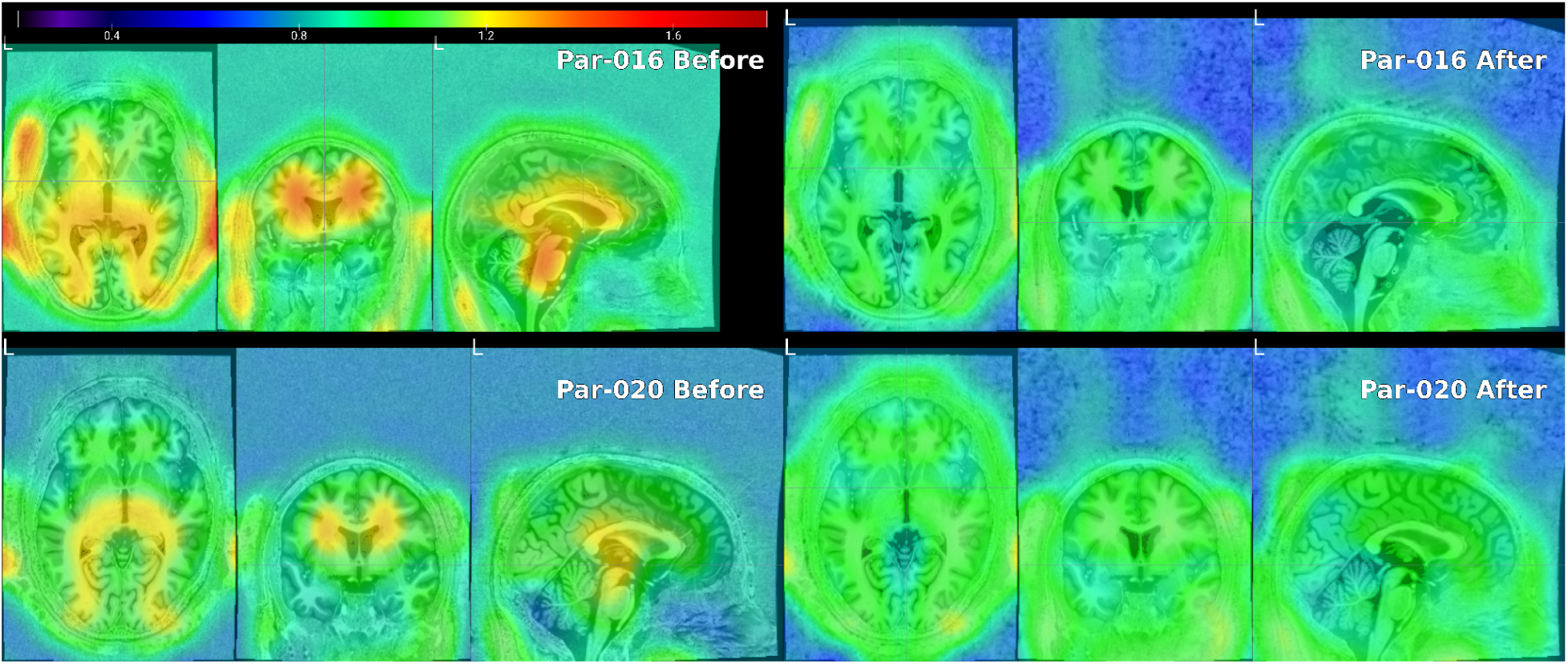
Bias field maps before and after B_1_^+^ inhomogeneity correction in a mutation carrier and a matched control. Bias fields estimated with ANTs’ *N4BiasFieldCorrection* (Tustison et al., 2010) are shown overlaid on MP2RAGE images. The upper row shows a carrier (par-016) whose acquisition before B_1_ ^+^ inhomogeneity failed the MRIQC pipeline, the lower row shows the age and sex matched control (par-020). Before correction (left column), the carrier exhibited greater intensity non-uniformity than the control. After correction (right column), bias field magnitude was substantially reduced and appeared comparable between participants.

**Figure 3.**
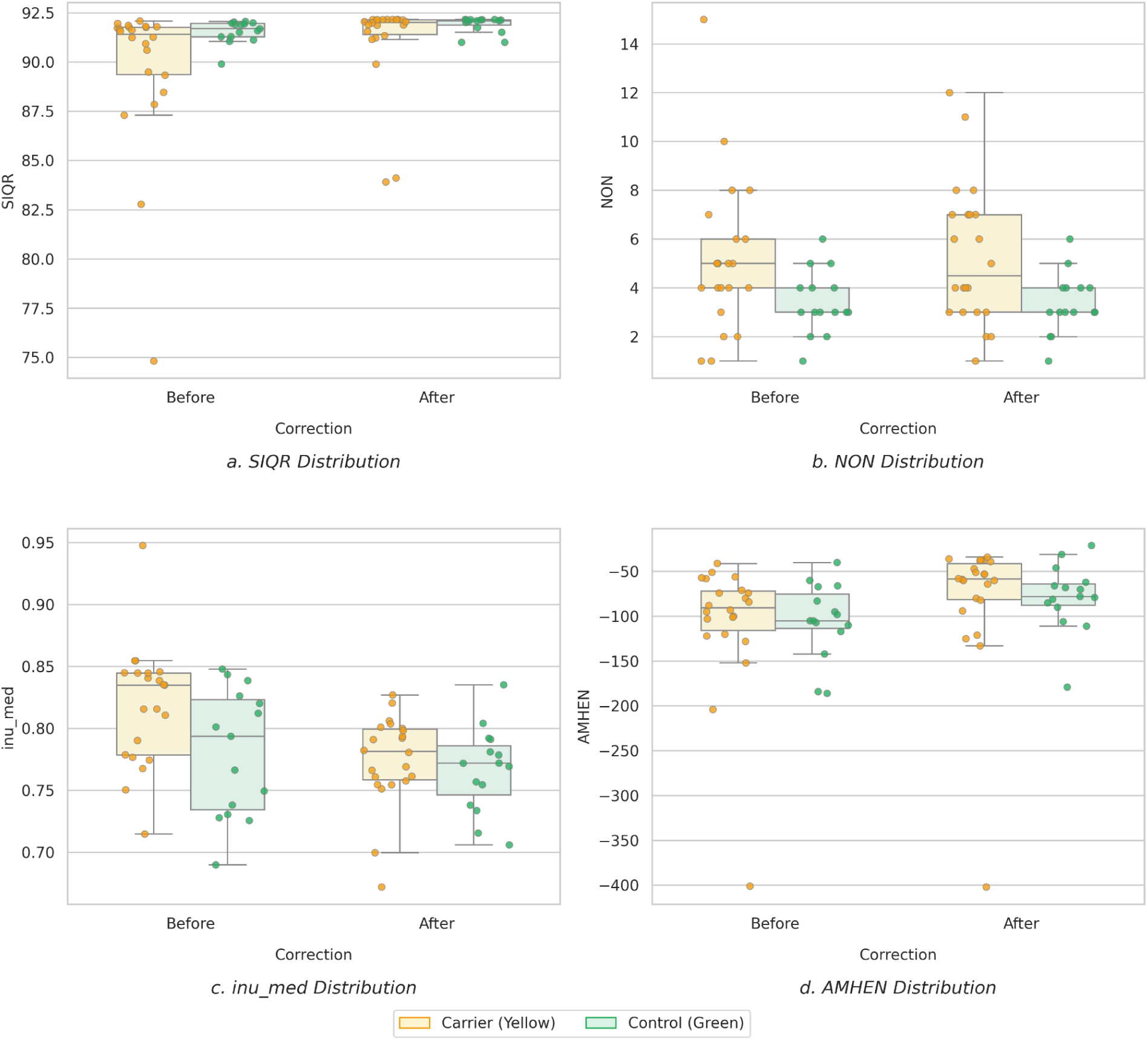
Image quality metrics before and after B_1_^+^ inhomogeneity correction. (a) CAT Structural Image Quality Rating (SIQR), (b) *fsqc* Normative Outlier Number (NON), (c) MRIQC Intensity Non-Uniformity field median (*inu_med*), and (d) FreeSurfer’s Arithmetic Mean of Hemispheric Euler Numbers (AMHEN). Each point represents an individual participant before and after B_1_^+^ inhomogeneity correction. Yellow markers denote mutation carriers and green markers denote healthy controls. B_1_ ^+^ inhomogeneity correction significantly improved SIQR, reduced residual intensity non-uniformity (*inu_med*), and increased AMHEN.

#### Inu_med and AMHEN

Although neither binary classifier (“Stanford RS” or “Oxford RUS”) was sensitive to B_1_^+^ inhomogeneity correction, several component metrics showed clear improvements in image quality. MRIQC-derived *inu_med*, a measure of residual intensity non-uniformity, decreased significantly (*p* < 0.001) after B_1_ ^+^ inhomogeneity correction, reflecting reduced bias field effects (Figure 3c). No significant effects of group (*p* = 0.134), age (*p* = 0.360) or sex (*p* = 0.744) were observed.

Similarly, the FreeSurfer-derived AMHEN increased significantly (*p* < 0.001) after B_1_ ^+^ inhomogeneity correction (Figure 3d). Higher AMHEN values indicate fewer topological defects during cortical surface reconstruction, suggesting improved reconstruction quality following correction. As with inu_med, no significant contributions from sex (*p* = 0.374), age (*p* = 0.639), or group (*p* = 0.803) were detected.

### 3.2. Morphometric analyses

B_1_ ^+^ inhomogeneity correction altered the number and distribution of significant ROIs (α = 0.05) across nearly all morphometric analyses, primarily by increasing the sensitivity of statistical inference in cortical parcellations (Figure 4).

**Figure 4.**
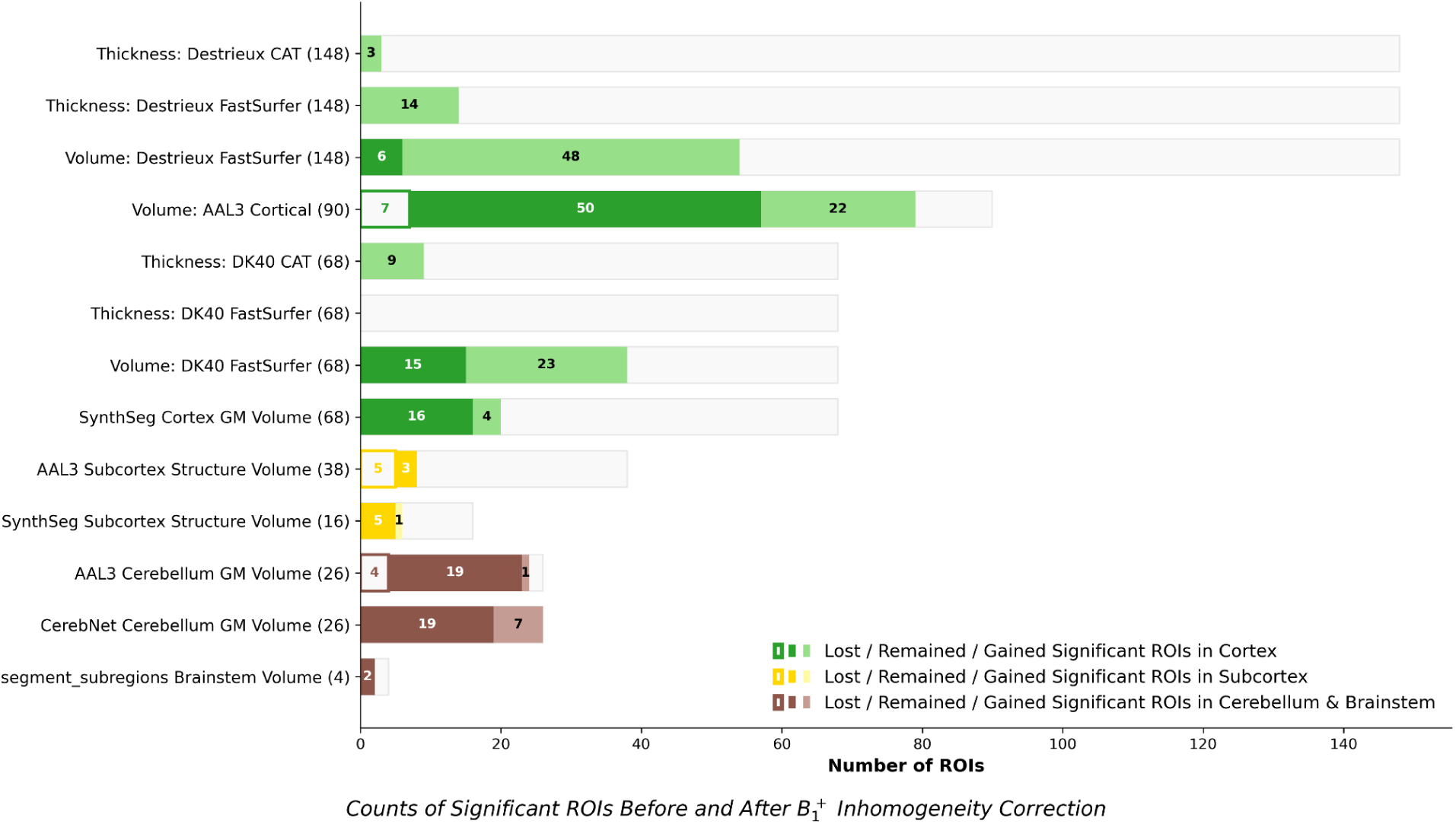
Method-dependent effects of B_1_^+^ inhomogeneity correction on morphometric findings. For each atlas or segmentation framework, the total number of evaluated ROIs is shown in parentheses. Blank boxes represent ROIs that lost significance after correction, solid boxes represent ROIs that remained significant before and after correction, and light-colored boxes represent ROIs that gained significance following correction. Differences in the distribution of lost, stable and gained ROIs highlight how B_1_ ^+^ inhomogeneity correction alters case-control inferences across morphometric pipelines.

#### CAT: mean cortical thickness analysis (DK40 and Destrieux)

Cortical thickness analysis demonstrated a substantial improvement in statistical sensitivity following B_1_^+^ inhomogeneity correction across both evaluated brain atlases (Figure 5, Supplementary Material 6a, 6e). Prior to B_1_ ^+^ inhomogeneity correction, no individual cortical region reached statistical significance in either atlas, while after B_1_ ^+^ inhomogeneity correction, multiple cortical parcellations gained statistical significance. Specifically, within the DK40 atlas, 9 cortical parcellations (6 occipital, 2 parietal and 1 cingulate) gained statistical significance post-correction. The Destrieux atlas mirrored the enhanced sensitivity pattern, where 3 cortical parcellations (2 occipital and 1 parietal) gained statistical significance following the B ^+^ inhomogeneity correction.

**Figure 5.**
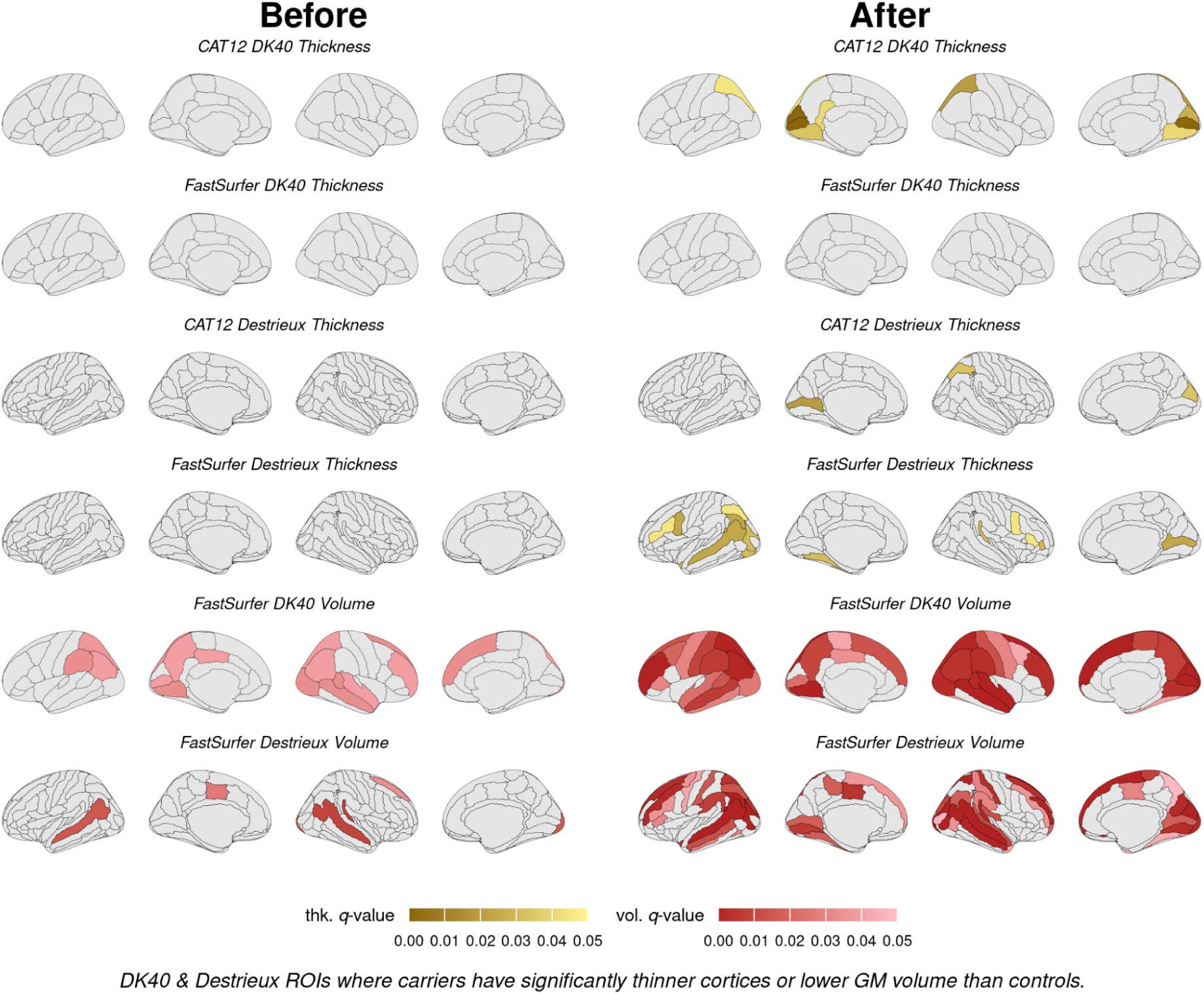
Impact of B_1_^+^ inhomogeneity correction on morphometric case-control findings. Significant regional differences in cortical thickness (thk.) and gray matter volume (vol.) between mutation carriers and controls are shown before and after B_1_^+^ inhomogeneity correction. Across nearly all atlases and segmentation frameworks, correction increased sensitivity to morphometric differences, although the magnitude and spatial distribution of effects varied by analysis method.

#### FastSurfer: mean cortical thickness analysis (DK40 and Destrieux)

The effect of B_1_ ^+^ inhomogeneity correction on FastSurfer cortical thickness estimates depended on atlas granularity (Figure 5, Supplementary Material 6b, 6f). Prior to B_1_ ^+^ inhomogeneity correction, no individual cortical parcellation reached statistical significance in either the DK40 or the Destrieux atlas. Following B_1_ ^+^ inhomogeneity correction, the DK40 atlas still did not yield significant ROIs, although the *q*-value of several frontal, parietal, and temporal lobes regions approached significance. Conversely, the more granular Destrieux atlas identified 14 distinct cortical parcellations that gained statistical significance, which were distributed across both hemispheres (5 frontal, 4 occipital, 3 temporal and 2 parietal).

#### FastSurfer: GM volume analysis (DK40 and Destrieux)

Cortical parcellations GM volume analysis demonstrated a substantial improvement in statistical sensitivity following B_1_ ^+^ inhomogeneity correction across both evaluated brain atlases (Figure 5, Supplementary Material 6c, 6g). In the DK40 atlas, no cortical parcellations lost significance, while 15 remained significant (5 frontal, 4 temporal, 4 parietal and 2 occipital), and 23 gained statistical significance (5 frontal, 6 temporal, 4 parietal, 6 occipital, 1 cingulate, and 1 insular). The Destrieux atlas amplified this enhanced sensitivity pattern. No cortical parcellations lost significance, 6 remained significant (2 frontal, 2 temporal and 2 occipital), and 48 gained statistical significance (14 frontal, 14 temporal, 7 parietal, 9 occipital, 2 limbic/cingulate, and 2 insular).

#### CAT: AAL3 whole-brain ROIs volume analysis

AAL3-based volumetric analyses demonstrated a redistribution of significant findings after B_1_ ^+^ inhomogeneity correction (Figure 6, Supplementary Material 6d, 6i, 6j). In the cerebral cortex, 7 parcellations lost significance (4 frontal and 3 temporal), 50 remained significant (14 frontal, 14 temporal, 10 occipital, 10 parietal, and 2 insula), and 22 gained significance (17 frontal, 3 temporal, and 2 parietal). In the subcortex, 5 regions lost significance, while 3 remained significant, and no regions gained significance. In the cerebellum, 4 subregions lost their statistical significance, 19 remained significant, and 1 gained significance after B ^+^ inhomogeneity correction.

**Figure 6.**
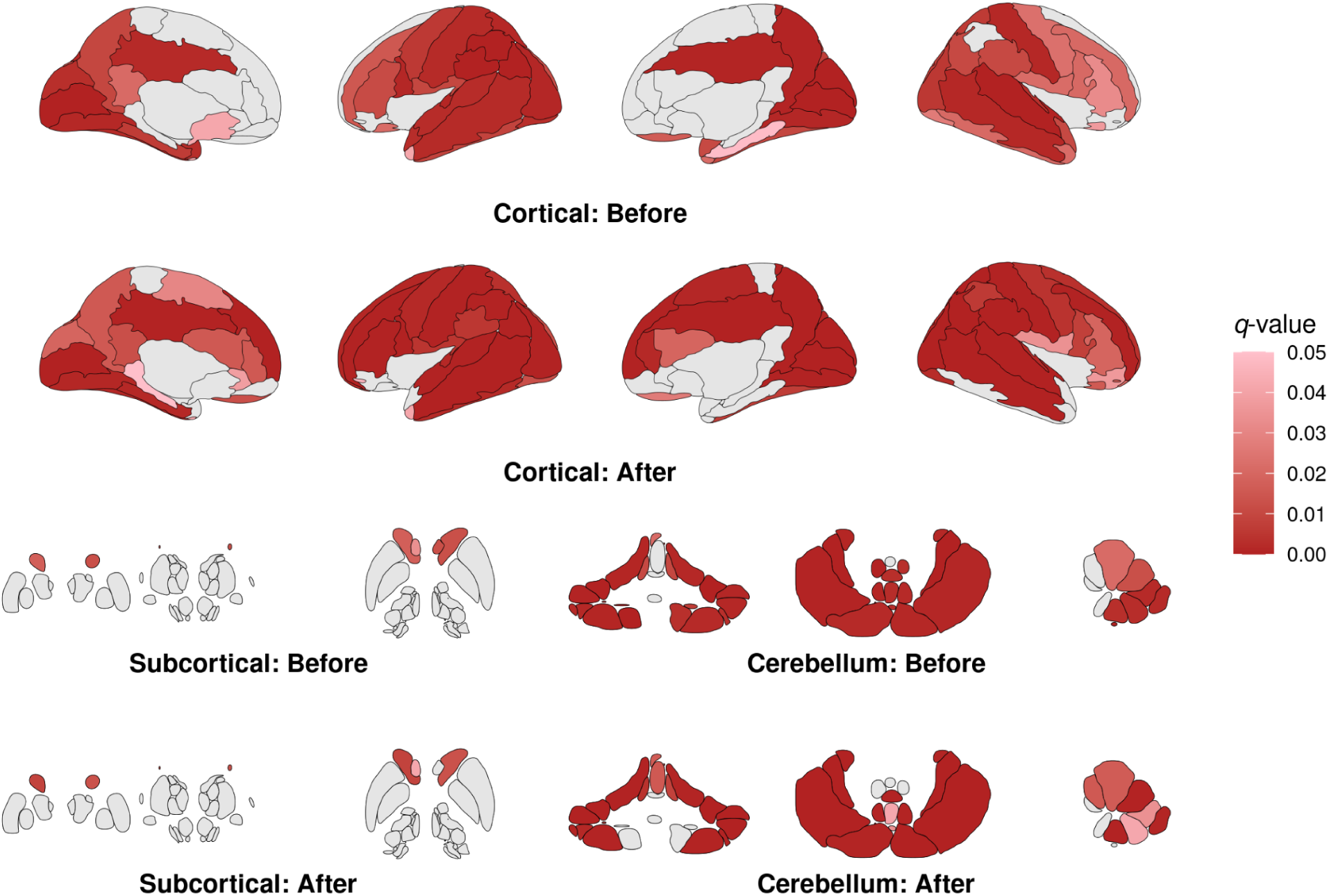
Significant ROIs of CAT AAL3 volume analysis. Before & After: before and after B_1_^+^ inhomogeneity correction. Across structures of the cortex, subcortex and cerebellum, the significant ROIs demonstrated a slight redistribution.

#### SynthSeg parcellation ROIs volume

SynthSeg demonstrated relatively robust and stable performance against B_1_^+^ inhomogeneity effects (Figure 7, Supplementary Material 6h). At the cortical level, 16 parcellations remained statistically significant after B_1_^+^ inhomogeneity correction, comprising 5 parietal regions (bilateral inferior parietal, bilateral postcentral, and right supramarginal), 4 temporal regions (left entorhinal, bilateral middle temporal, and right fusiform), 3 frontal regions (bilateral rostral middle frontal and right precentral), 3 occipital regions (left lingual, right cuneus, and right lateral occipital), and 1 cingulate region (left posterior cingulate). Additionally, 4 cortical regions gained significance after B_1_ ^+^ correction, including 2 temporal regions (left fusiform and left superior temporal) and 2 frontal regions (right paracentral and right superior frontal), while none lost significance. Subcortically, 5 specific structures remained significant (bilateral caudate, bilateral nucleus accumbens, and the right putamen), while 1 region gained significance (right ventral diencephalon), and no regions lost significance. For summary structures, bilateral cerebral cortex volumes (*q*_left-before_ = 0.0004, *q*_left-after_ < 0.0001; *q*_right-before_ < 0.0001, *q*_right-after_ < 0.0001) and brainstem as a whole (*q*_before_ = 0.0329, *q*_after_ = 0.0212) remained significant before and after B_1_ ^+^ inhomogeneity correction. Notably, bilateral cerebral cortex volumes changed from non-significant to marginal (*q*_left-before_ = 0.1088, *q*_right-before_ < 0.1614; *q*_left-after_ < 0.0564, *q*_right-after_ < 0.0772).

**Figure 7.**
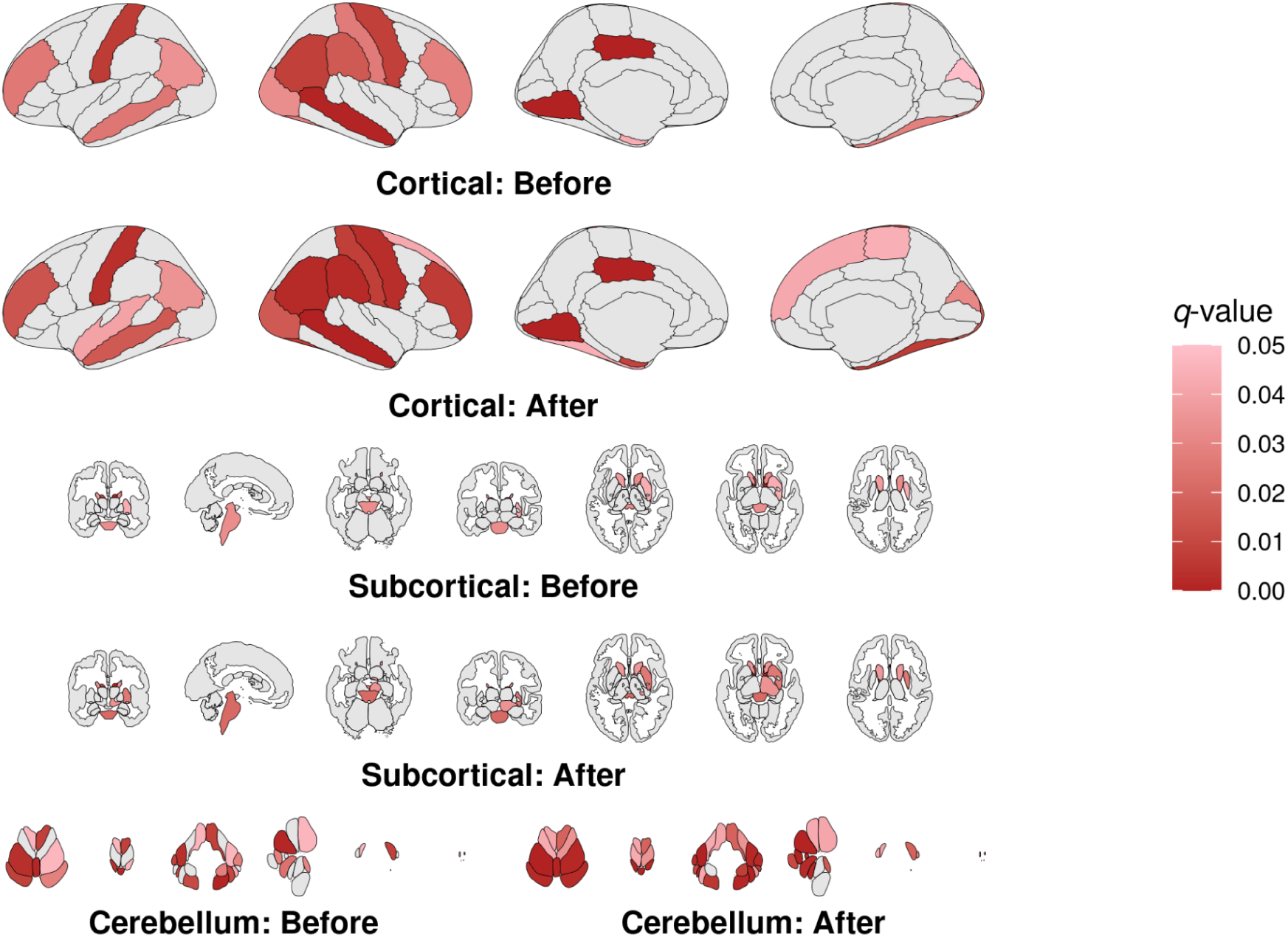
Significant ROIs of SynthSeg and CerebNet volume analysis. Before & After: before and after B_1_^+^ inhomogeneity correction. Across structures of cortex, subcortex and cerebellum, the sensitivity of morphometric statistical inference increased.

#### CerebNet and brainstem subregion analyses

CerebNet volume analysis demonstrated a clear gain in statistical sensitivity following B_1_ ^+^ inhomogeneity correction (Figure 7, Supplementary Material 6k). At the global level, bilateral cerebellar cortex and left cerebellar white matter volumes remained statistically significant, whereas right cerebellar white matter volume lost significance (*q*_before_ = 0.0306, *q*_after_ = 0.0564). At the cerebellar subregional level, no lobules lost significance: 19 subregions remained significant, and 7 newly gained significance after B_1_ ^+^ inhomogeneity correction (bilateral lobule V, left Crus II, left lobule VIIb, left lobule VIIIa, left lobule VIIIb, and left lobule X). For the *segment_subregions brainstem* segmentation, the medulla and pons volumes remained statistically significant, while the whole brainstem volume newly gained significance (*q*_before_ = 0.0526, *q*_after_ = 0.0398).

## 4. DISCUSSION

Translating structural MRI pipelines, including acquisition and preprocessing steps, from healthy controls to clinical populations requires careful consideration of both anatomical variability and underlying pathophysiology. In contrast to focal lesion disorders, where structural abnormalities are prominent but often disrupt preprocessing pipelines (Muthusivarajan et al., 2024), metabolic conditions, such as the m.3243A>G mitochondrial gene mutation, present a more subtle challenge. These populations typically exhibit diffuse and early-state alterations that precede macroscopic lesions (Alfaro et al., 2018; Ford, 2025; Hatch et al., 2023), providing a stringent test of both image quality control and morphometric sensitivity (de Jong et al., 2024). In this context, our findings demonstrate that pipelines validated in healthy populations do not necessarily generalize to subtle disease phenotypes.

Importantly, these findings directly address the key gaps outlined in the introduction. While the impact of B_1_ ^+^ inhomogeneity on qT1 and T1w MRI has been previously demonstrated (Haast et al., 2018b, 2021), our results extend this understanding in three ways. First, commonly used automated QC approaches fail to reliably capture these effects at 7 T. Second, the effects of B_1_ ^+^ inhomogeneity are strongly method-dependent, rather than uniform across pipelines. Third, and most critically, B_1_ ^+^ inhomogeneity can alter statistical inferences in case-control morphometric studies, thereby influencing the interpretation of neurobiological findings.

### 4.1. Automated QC approaches

#### Binary classifications by pretrained models

While the “Stanford RF” classifier labeled nearly all datasets as low quality, and failed for the uncorrected image from one carrier, the “Oxford RUS” classifier labeled all datasets as good quality, irrespective of correction. Such opposing classifications challenge the usage of current automated QC approaches in 7 T MP2RAGE acquisitions, which were largely trained on 3 T MPRAGE data (Esteban, 2017; Bhalerao et al., 2025). Their reliance on visual ground truth further limits sensitivity to subtle intensity non-uniformities, which may be visually inconspicuous but influential for downstream processing. In our dataset, pipeline failure proved more informative than classification output: the failure occurred in a mutation carrier and was resolved after B_1_ ^+^ inhomogeneity correction, suggesting the interaction between m.3243A>G carriers’ brain anatomy and the increased B_1_ ^+^ inhomogeneity in UHF was fatal for MRIQC preprocessing but not well represented in training data (MacLennan et al., 2022). Taken together, this discrepancy demonstrates that binary predictions alone are insufficient and potentially misleading.

#### Composite rating and component metrics

In contrast to the binary classifiers, increases in SIQR indicated overall improvements in image quality. Both component-level intensity non-uniformity metric (i.e., *inu_med*) and topological proxies of image quality (i.e., AMHEN) provided consistent and interpretable results. Despite ongoing debate regarding the interpretation of AMHEN (Gilmore et al., 2021), the observed increase provides additional evidence of reduced intensity non-uniformity. These findings reinforce that B_1_ ^+^ inhomogeneity correction meaningfully improves image quality, even when binary classifiers fail to detect such changes.

Interestingly, the absence of a significant change in the *fsqc* NON should not be interpreted as evidence that B_1_ ^+^ inhomogeneity is unnecessary. Rather, it likely reflects the relative robustness of modern deep learning-based preprocessing pipelines to residual intensity inhomogeneity. However, robustness of preprocessing does not necessarily translate into robustness of downstream analyses. As demonstrated by the morphometric results, and discussed in the following section, substantial changes in case-control inferences were observed despite largely stable *fsqc* scores. This distinction highlights an important point: quality control metrics may indicate successful image processing while remaining insensitive to biases that influence the final neurobiological conclusions.

### 4.2. Method-dependent effects on morphometric analyses

A key insight from this study is that B_1_ ^+^ inhomogeneity does not exert a uniform effect across the morphometric pipelines. Instead, its impact depends on the assumptions embedded within each processing framework, leading to method-dependent effects on morphometric analyses.

#### Surface resolution and atlas granularity, GM volume versus mean cortical thickness

To minimize confounding from partial volume effects, cortical morphometry was evaluated using both registration-based (CAT) and deep learning-based (FastSurfer) surface reconstruction at the native 0.7 mm isotropic resolution, while extracting GM volume and mean cortical thickness from identical atlases (DK40 and Destrieux) (Zaretskaya et al., 2018; Nian et al., 2023). Nevertheless, morphometric sensitivity remained dependent on both the selected metric and atlas granularity.

Consistent with previous observations, mean cortical thickness showed greater specificity and robustness than GM volume (Winkler et al., 2010). In addition, the more granular Destrieux atlas consistently revealed more significant regions than the coarser DK40 atlas, highlighting how atlas boundaries can influence detection of subtle effects. The relative location of significant clusters and atlas boundaries may conceal true positive findings. In this context, surface- and volume-based morphometry provides the highest granularity possible for mean cortical thickness and GM volume analyses, respectively (Dale et al., 1999; Fischl et al., 1999; Ashburner & Friston, 2000). Notably, the distribution of significant cortical thickness findings obtained with FastSurfer and the Destrieux atlas after B_1_ ^+^ inhomogeneity correction closely resembled our previously reported surface-based morphometry results (Haast et al., 2018a).

#### Registration-based versus deep learning-based pipelines

Registration-based whole-brain segmentation pipeline (CAT: AAL3) showed a redistribution of statistical significance. This reflected its reliance on the intensity distribution of its spatial priors, making it particularly sensitive to the presence of bias fields. In contrast, the relative robustness of deep learning-based pipelines (SynthSeg, CerebNet) was further demonstrated, although B_1_ ^+^ inhomogeneity remains a relevant confound.

#### Implications for case-control studies

Crucially, these method-dependent effects directly translate into differences in morphometric findings in case-control studies. Regions that were non-significant before became significant after B_1_ ^+^ inhomogeneity correction, and vice versa, depending on the pipeline. This demonstrates that B_1_ ^+^ inhomogeneity is not merely a technical nuisance, but a factor that can alter morphometric statistical interference. As such, it may contribute to inconsistencies across studies, particularly in clinical populations with subtle pathologies.

### 4.3. Limitations

This study has several limitations. First, analyses were restricted to 7 T “uniform MP2RAGE” acquisitions, and findings may not generalize to other sequence variants (O’Brien et al., 2014; Tanner et al., 2012). Second, B_1_ ^+^ correction was performed using SA2RAGE-based mapping, while alternative correction strategies may yield different results (Kanakaraj et al., 2024; Pawade et al., 2025; Ruijters et al., 2025). Third, legacy MRIQC configurations were used for comparability with prior studies, which may underestimate the performance of newer implementations (Esteban et al., 2019). Finally, statistical analyses were not designed for definitive biomarker discovery, but rather to illustrate the impact of B_1_ ^+^ inhomogeneity.

## 5. CONCLUSIONS

By focusing on a mitochondrial gene mutation cohort with subtle structural alterations, this study provides a more stringent evaluation of neuroimaging pipelines than traditional validations in healthy populations. In such settings, where effect sizes are small and spatially diffuse, methodological biases become proportionally more influential. Our findings therefore demonstrate that validation in healthy cohorts alone is insufficient, and that clinical populations are essential for revealing hidden sensitivities in both image quality assessment and morphometric pipelines. Within this framework, B_1_^+^ inhomogeneity emerges as a critical, yet often underappreciated, confound in 7 T MP2RAGE imaging. While correction improves signal uniformity and generally increases sensitivity to group differences, its effects are strongly method-dependent and can alter morphometric statistical interference. As a result, B_1_ ^+^ inhomogeneity is not merely a technical nuisance, but a factor that can directly influence research conclusions and contribute to variability across studies. Importantly, current automated QC approaches do not reliably capture these effects, and even advanced deep learning pipelines do not fully compensate for an uncorrected bias. This underscores the need for both improved QC strategies and explicit B_1_ ^+^ inhomogeneity correction tailored to UHF imaging.

In summary, robust and reproducible neuroimaging at 7 T requires careful handling of B_1_ ^+^ inhomogeneity, alongside thoughtful selection of preprocessing methods. This is particularly critical for studies targeting subtle clinical phenotypes. To establish reliable morphometric biomarkers at UHF strengths, explicit B_1_ ^+^ inhomogeneity correction and carefully chosen preprocessing strategies are practically necessary and highly recommended.

### Take-home messages

- *Residual B ^+^ inhomogeneity at 7 T affects more than image appearance. It influences image quality metrics, preprocessing performance, and downstream morphometric analyses, even in datasets that appear visually acceptable and successfully complete automated processing pipelines*.
- *The impact of B_1_ ^+^ inhomogeneity is strongly method-dependent. Registration-based and deep learning-based frameworks respond differently to residual bias fields, resulting in changes in the number, location, and statistical significance of morphometric findings. Consequently, preprocessing choices can directly influence neurobiological interpretations and case-control inferences*.
- *Explicit B ^+^ correction should be considered best practice for 7 T MP2RAGE morphometry. Although modern deep learning pipelines improve robustness to image imperfections, they do not fully compensate for residual B_1_ ^+^ bias. Correction remains important for achieving reliable, reproducible, and biologically meaningful results, particularly in clinical populations and studies targeting subtle structural differences*.

## 6. ENDING SECTIONS

### 6.1. Data and Code Availability

Due to institutional privacy restrictions, raw and processed participant data are not publicly available. MRIQC, CAT, and FastSurfer pipelines were implemented using either default settings or options as specified in Methods. Custom shell scripts were utilized to convert SynthSeg outputs for CerebNet and brainstem subregion analyses (https://github.com/ADM-3A/M-BIC_S_2.022).

### 6.2. Author Contributions

Conceptualization: K.L., R.A.M.H., D.I.;

Methodology: K.L., R.A.M.H., D.I.;

Software: K.L., R.A.M.H.;

Formal analysis: K.L., R.A.M.H., D.I.;

Investigation: K.U., I.F.M.d.C., H.J.M.S., J.F.A.J.,

Resources: K.U., I.F.M.d.C., H.J.M.S., J.F.A.J., E.F., B.A.P.;

Writing – original draft: K.L., R.A.M.H., D.I.;

Writing – review & editing: All authors;

Supervision: R.A.M.H., D.I., B.A.P.;

Funding acquisition: K.U., E.F., I.F.M.d.C., B.A.P..

### 6.3. Funding

This work was supported by Maastricht University, the Netherlands Organization for Scientific Research (NWO; VIDI grant 452-11-002 to K.U.), Technology Foundation STW (12724 to E.F.) and Ride4Kids, Join4Energy and NeMo (to I.F.M.d.C.). K.L. was receiving the State Scholarship Fund (CSC 202208620032) as a living stipend from the Chinese Scholarship Council.

### 6.4. Declaration of Competing Interests

The authors report no competing interests.

## Data Availability

Due to institutional privacy restrictions, raw and processed participant data are not publicly available. MRIQC, CAT, and FastSurfer pipelines were implemented using either default settings or options as specified in Methods. Custom shell scripts were utilized to convert SynthSeg outputs for CerebNet and brainstem subregion analyses are available online at

https://github.com/ADM-3A/M-BIC_S_2.022

## 6.5. Acknowledgements

The authors are indebted to Prof. Dr. Andrew Webb (Leiden University Medical Centre, Leiden, Netherlands), Dr. José Marques (Donders Institute for Brain, Cognition and Behaviour, Nijmegen, Netherlands) and Dr. Florence van Tienen for providing the dielectric pads, the MATLAB code to perform the T1 correction and help with the genetic analyses used in this study, respectively.

## Appendix

*CAT SIQR*

CAT IQMs included :

- Noise-to-Contrast Ratio (NCR): assessed within optimized CSF and WM masks, estimates image noise by calculating the lowest average local standard deviation of voxel intensities in the bias-corrected image.
- Inhomogeneity-to-Contrast Ratio (ICR): assessed within optimized WM masks, evaluates intensity variations across the image by calculating the global standard deviation of smoothed intensities.
- Resolution Score (RES): accounts for distortions due to anisotropic resolution, directly computed using the root mean square (RMS) equation.
- Edge-to-Contrast Ratio (ECR): additional measure which captures the average slope of intensity changes at the GM/WM boundary, assesses the sharpness of tissue interfaces.
- Full-brain Euler Characteristic (FEC): This metric quantifies the topological integrity of the WM brain interface, helping to detect potential distortions caused by noise and artifacts.

These metrics were combined into a composite SIQR, reported as both percentage and categorical grade (Dahnke et al., 2025).

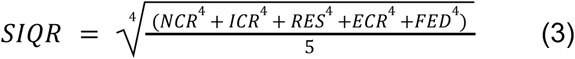

**Supplementary Material 1.** Demographics of participants.

Demographics of participants.
|  | Mutation Carrier | Healthy Control |
| --- | --- | --- |
| Sample size | 22 | 15 |
| Sex (female : male) ratio | 18 : 4 | 11 : 4 |
| Age ($\mu \pm \sigma$) years | 42.0 ± 10.9 | 38.4 ± 13.5 |
| Body Mass Index ($\text{kg} \cdot \text{m}^{-2}$) | 23.04 ± 3.51 | 24.43 ± 4.10 |
| Fasting Plasma Glucose (mmol/L) | 6.56 ± 1.66 | 4.99 ± 0.43 |
| Glycerated Hemoglobin (%) | 6.55 ± 1.21 | 5.02 ± 0.62 |

**Supplementary Material 2.**
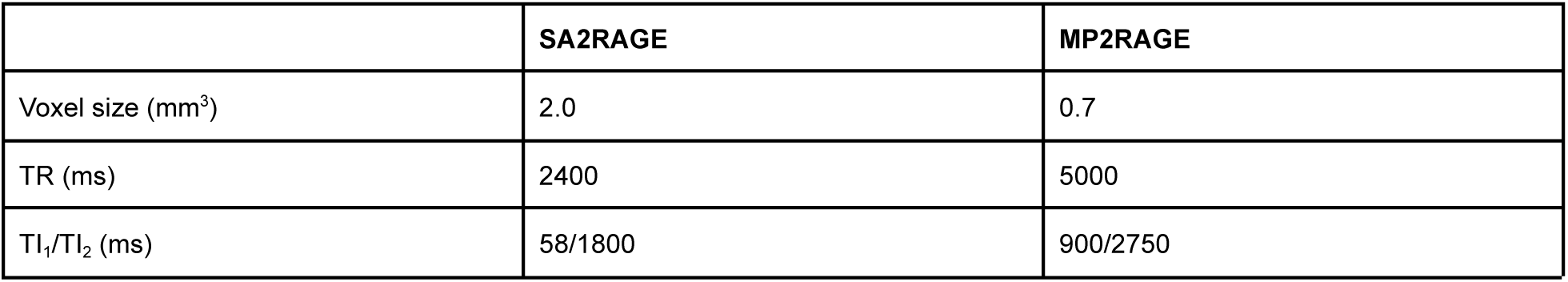
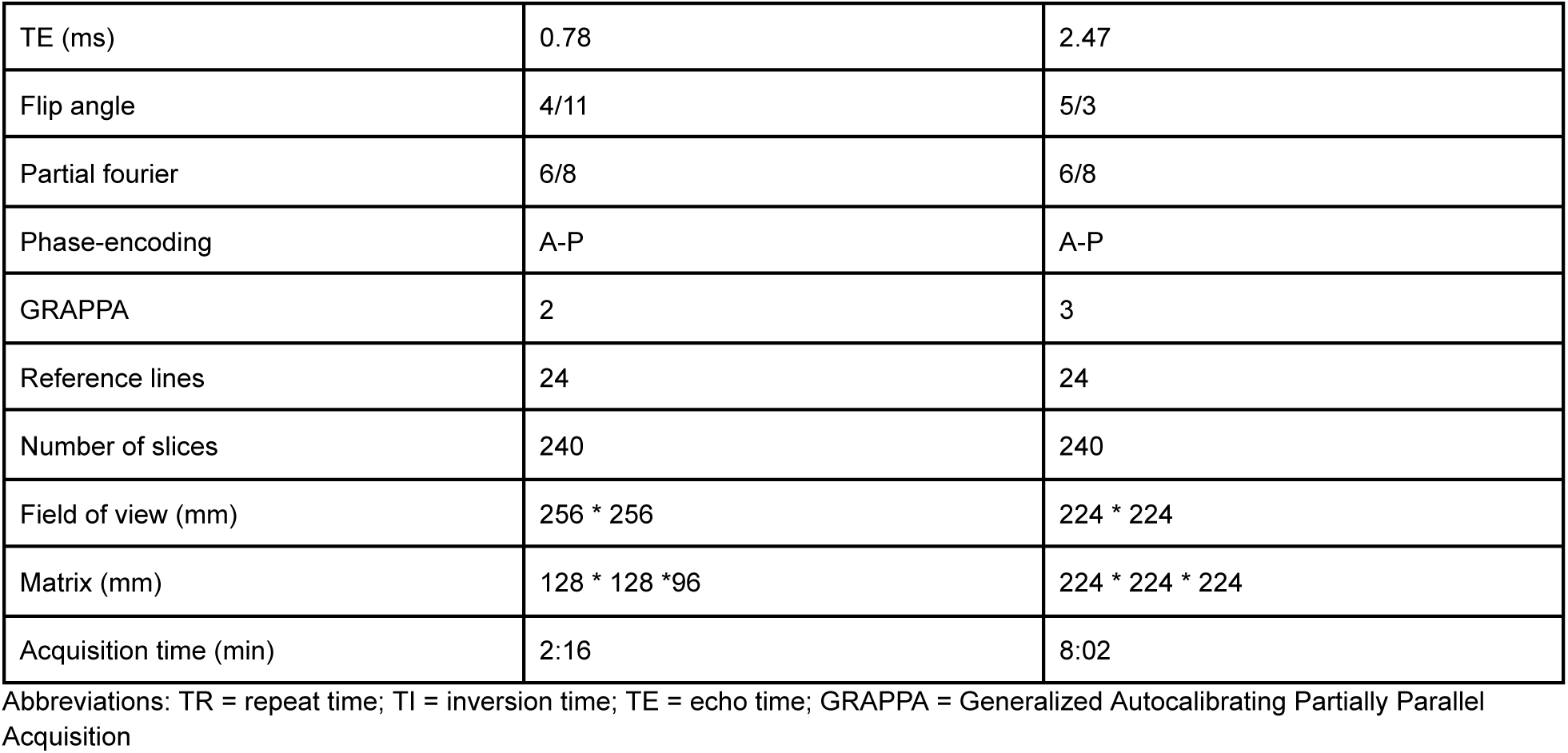
MRI sequence parameters.

**Supplementary Material 3.**
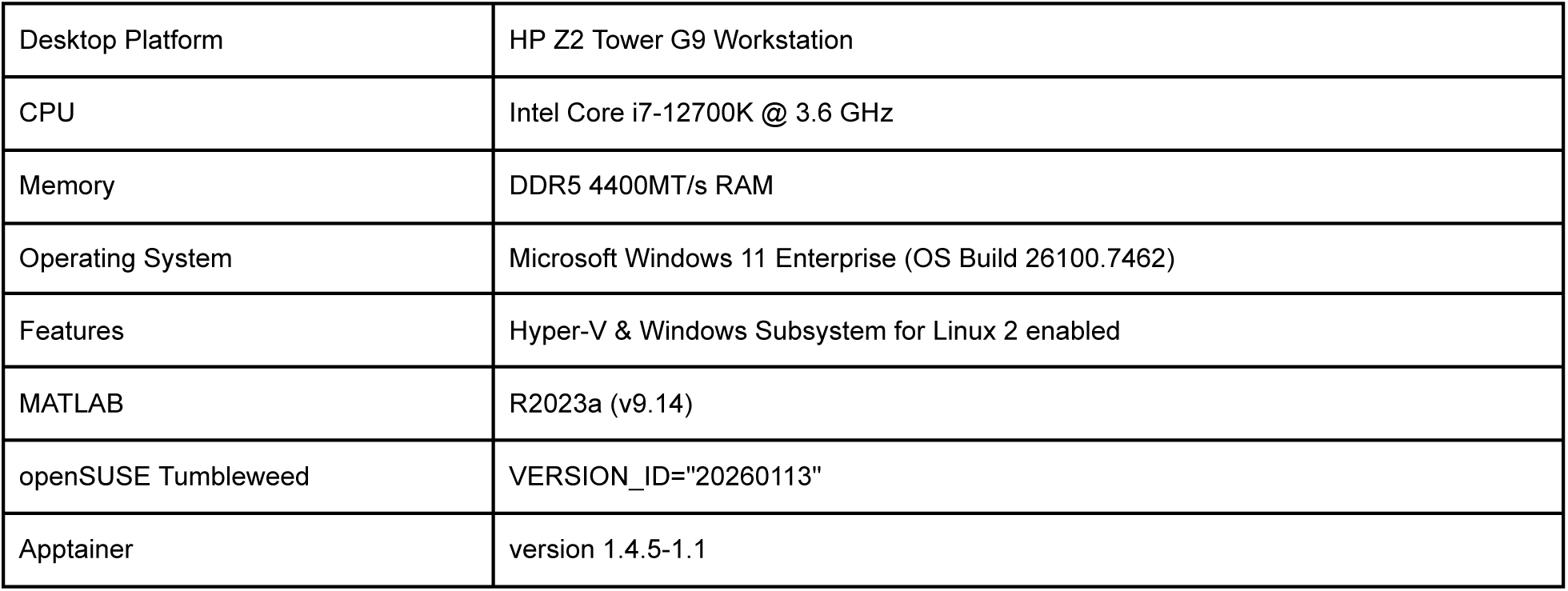
Preprocessing workstation specifications.

Preprocessing workstation specifications.
|  |  |
| --- | --- |
| Desktop Platform | HP Z2 Tower G9 Workstation |
| CPU | Intel Core i7-12700K @ 3.6 GHz |
| Memory | DDR5 4400MT/s RAM |
| Operating System | Microsoft Windows 11 Enterprise (OS Build 26100.7462) |
| Features | Hyper-V & Windows Subsystem for Linux 2 enabled |
| MATLAB | R2023a (v9.14) |
| openSUSE Tumbleweed | VERSION_ID="20260113" |
| Apptainer | version 1.4.5-1.1 |

**Supplementary Material 4.**
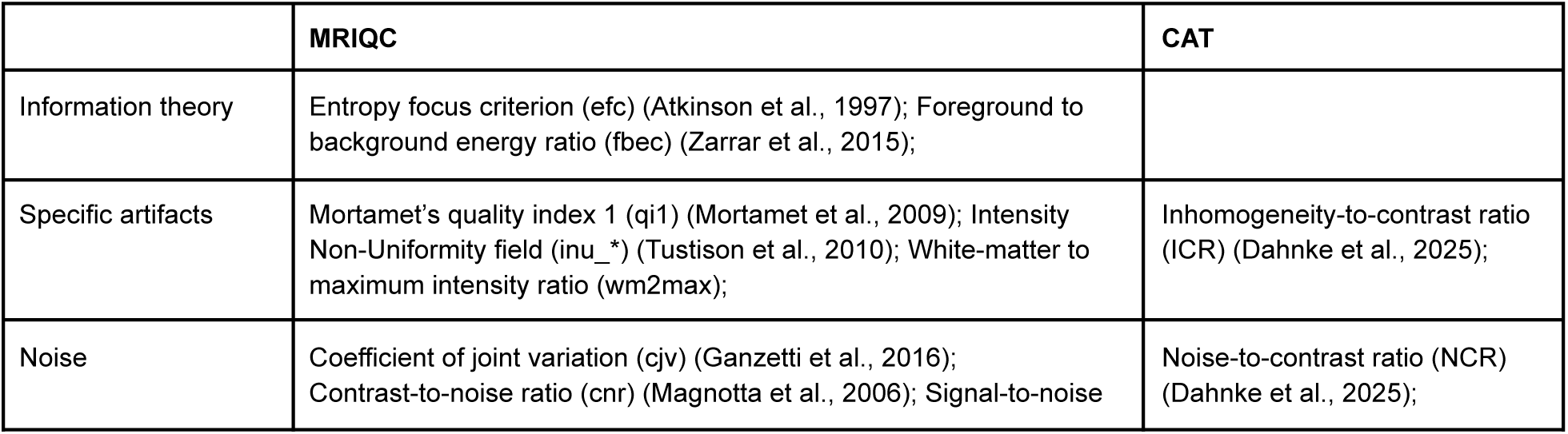
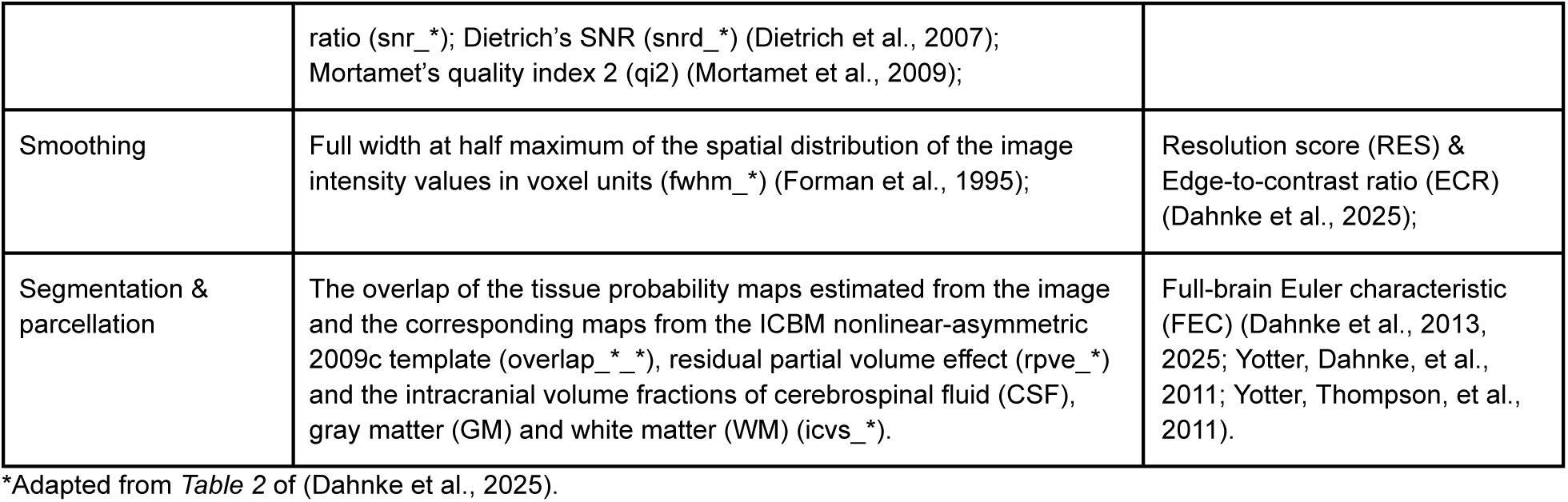
Homologous MRIQC & CAT image quality metrics*

**Supplementary Material 5.**
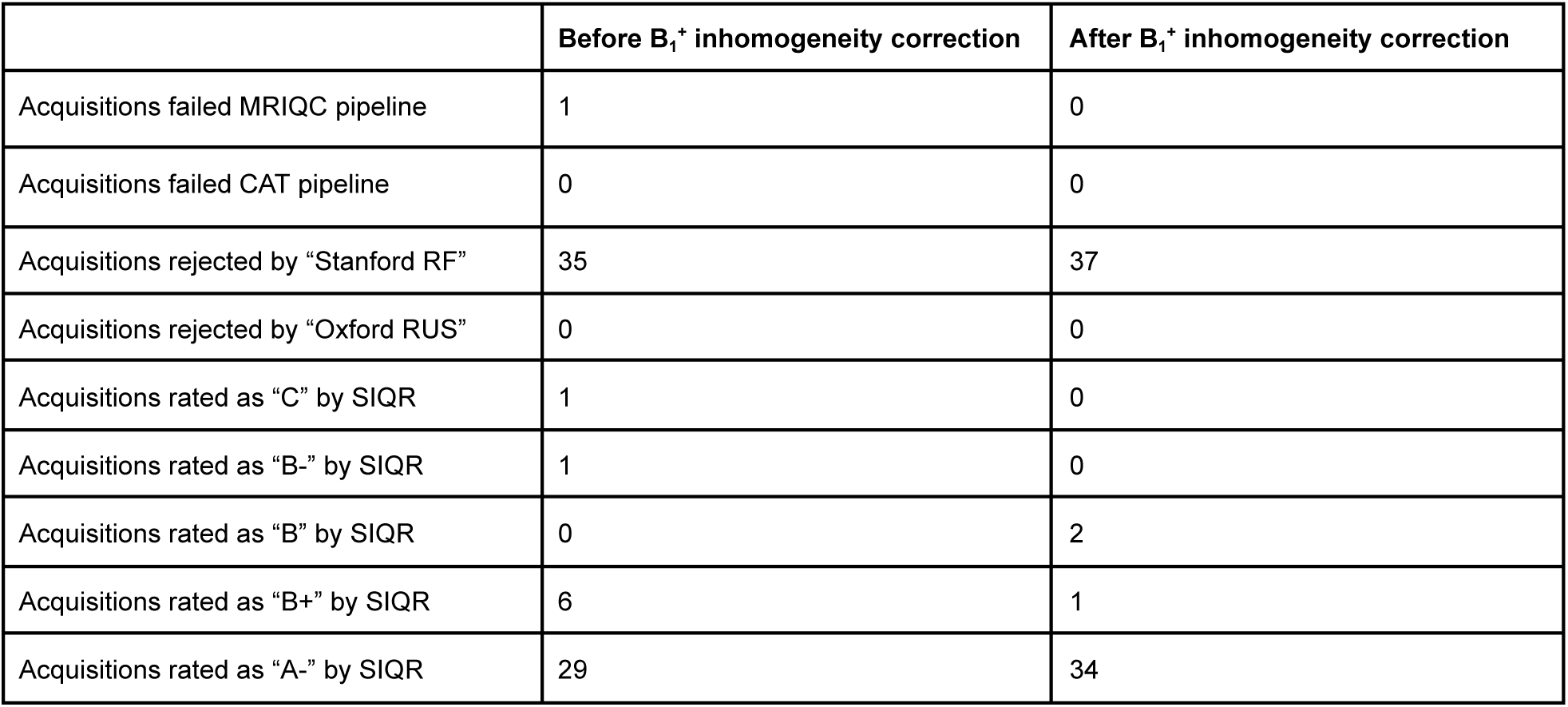
Pipeline failure cases.

**Supplementary Material 6a.**
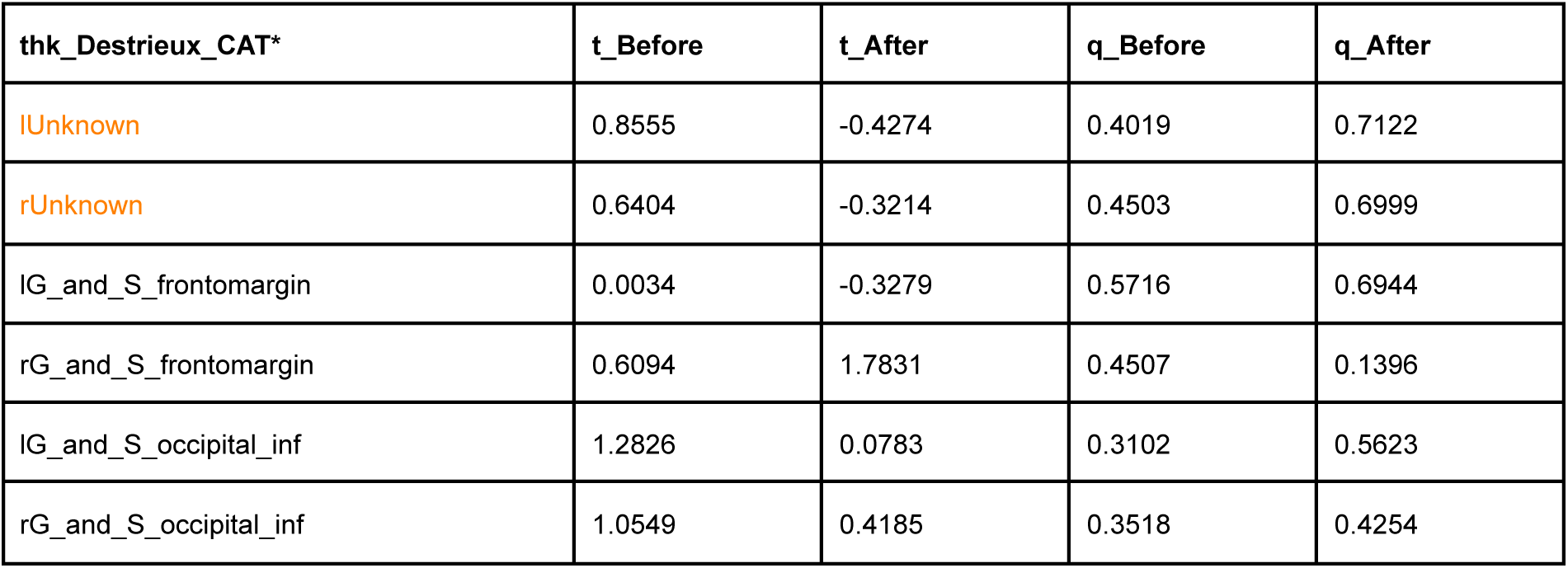
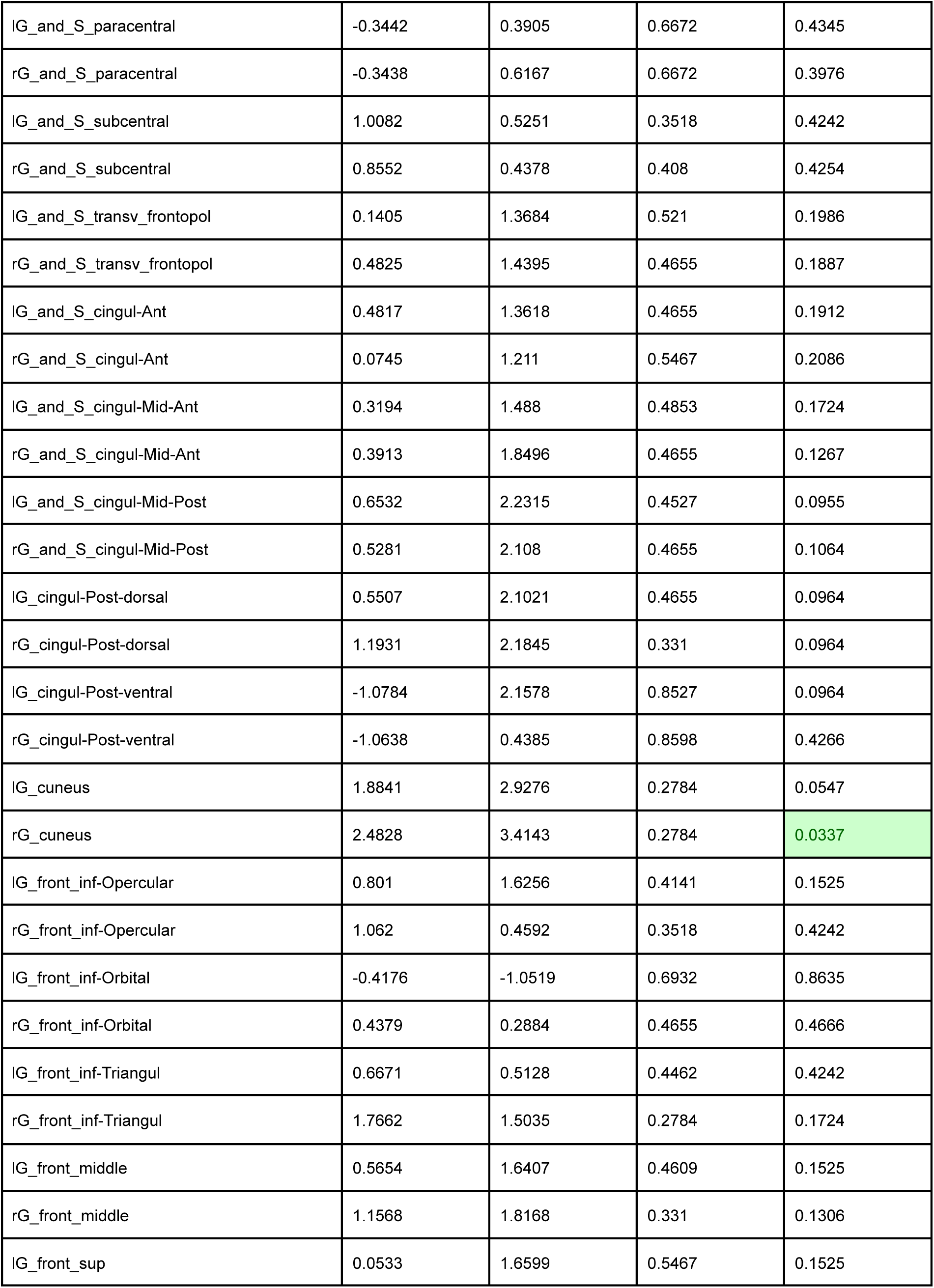
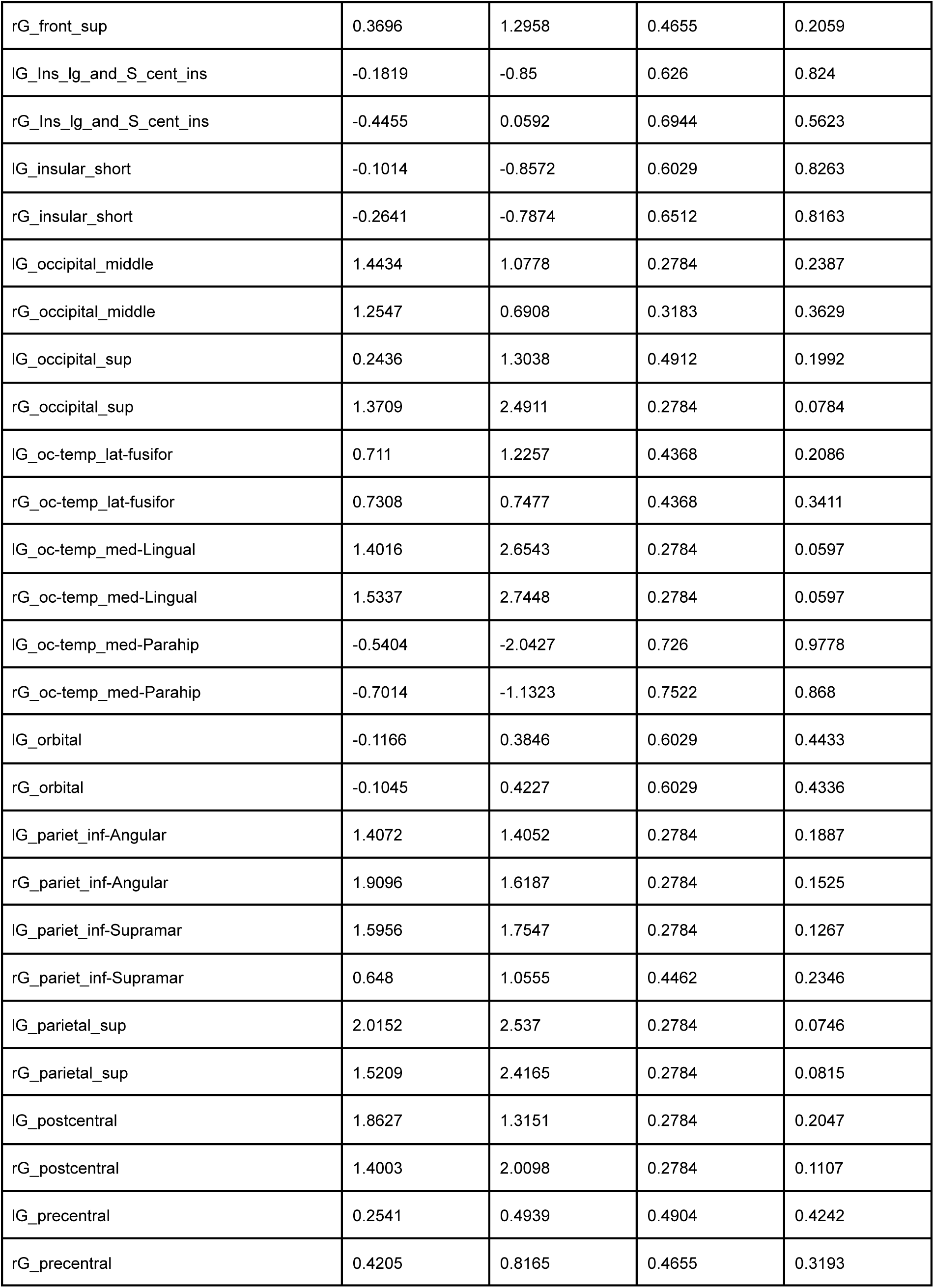
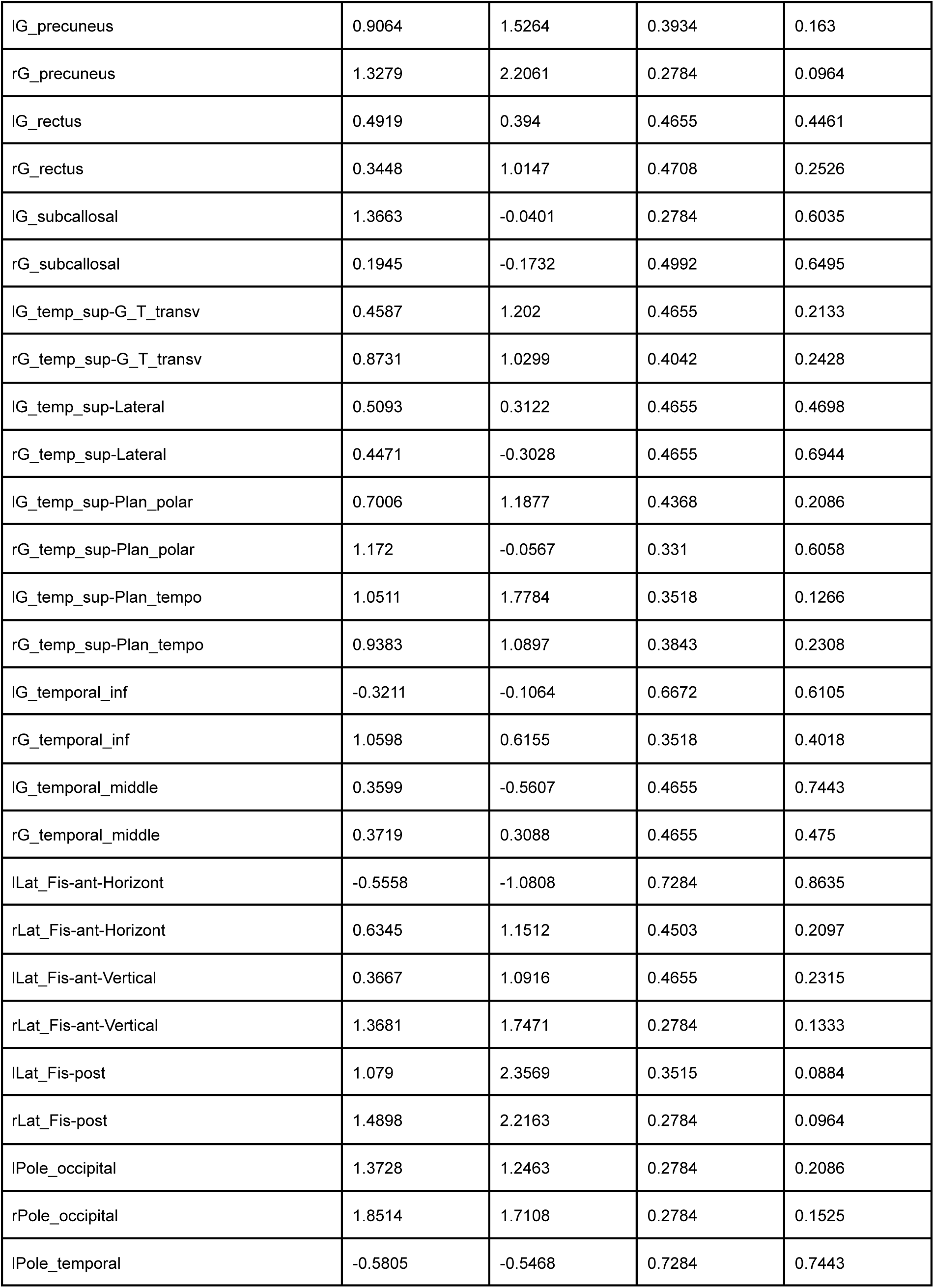
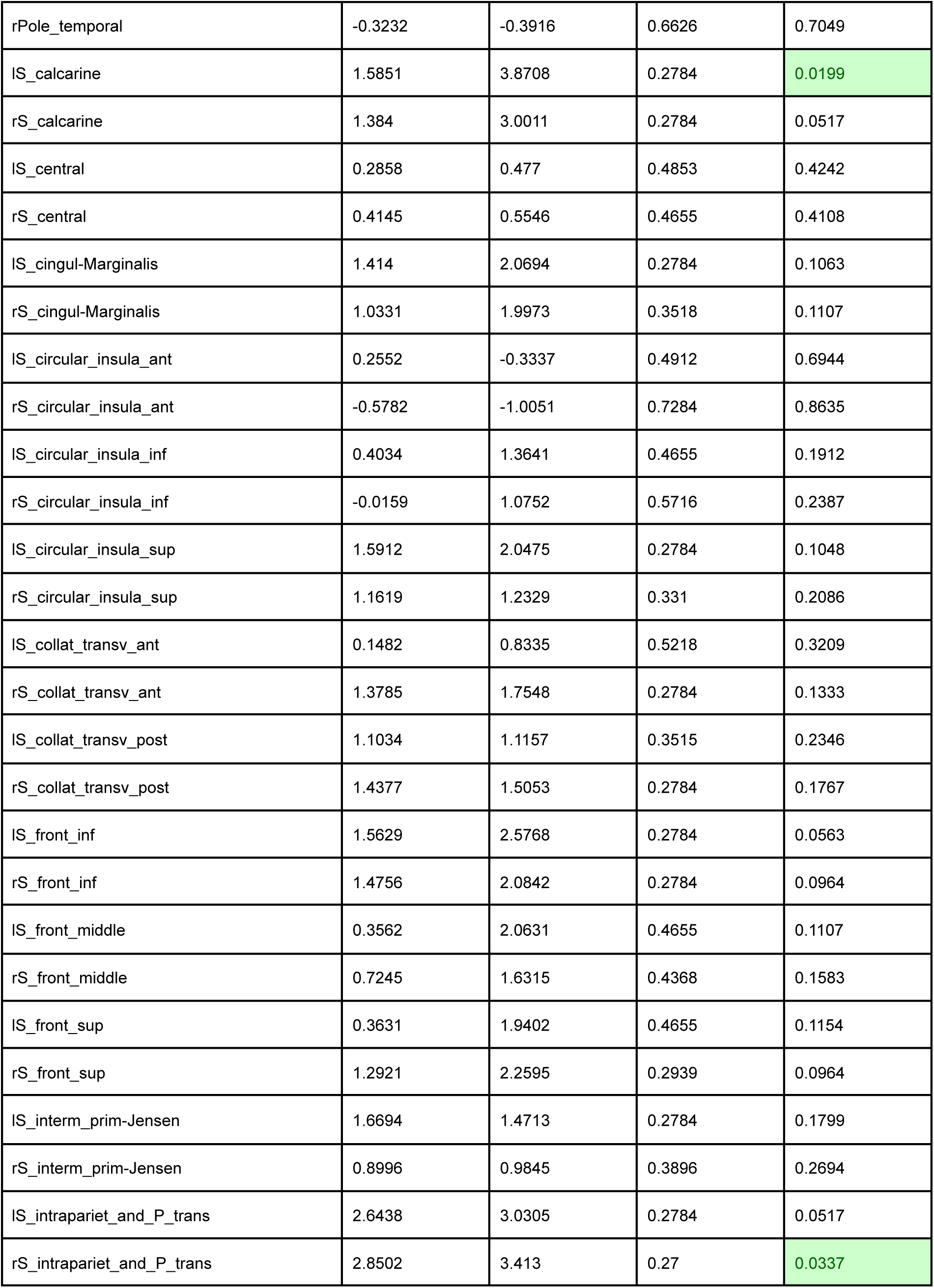
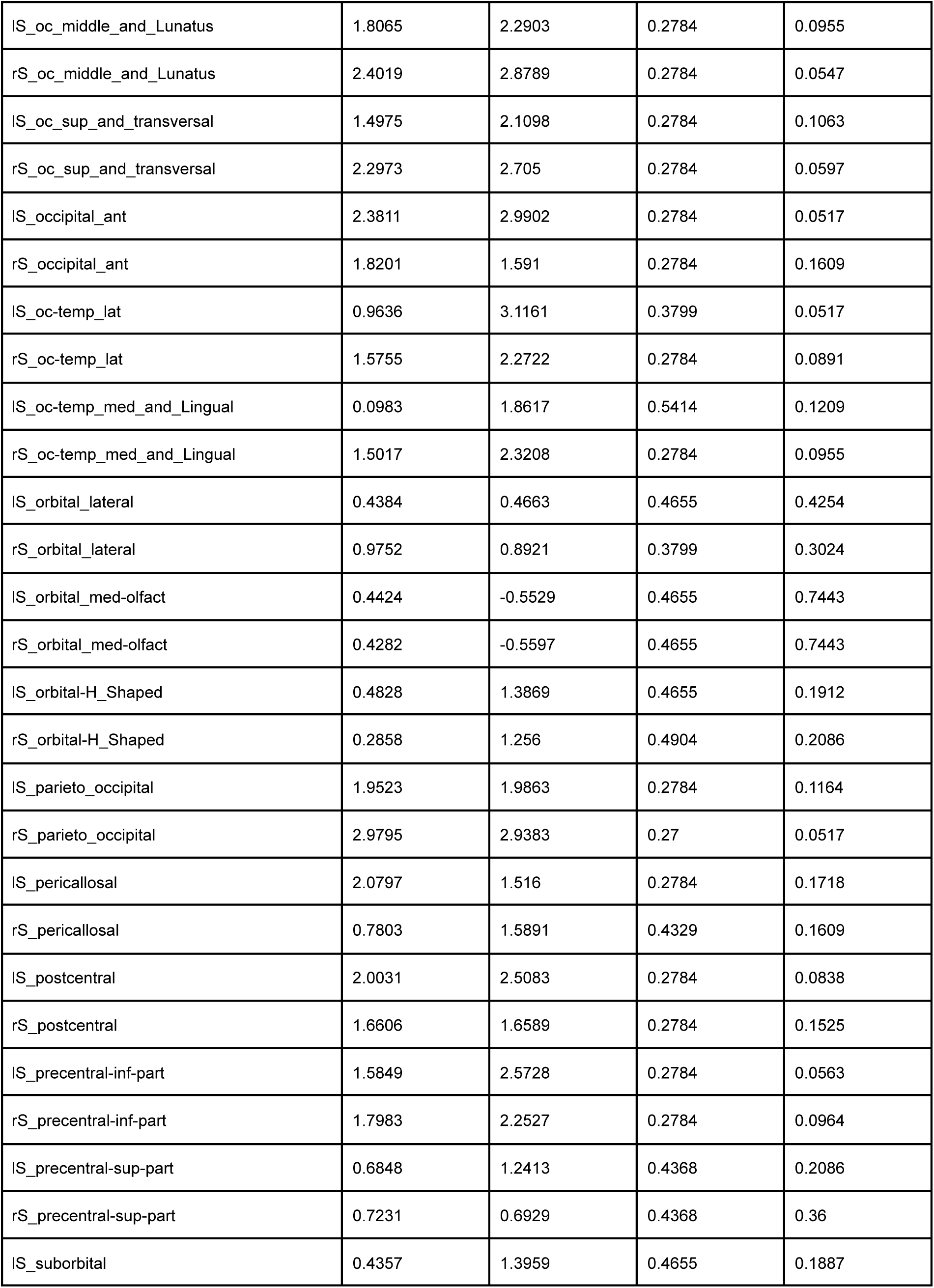
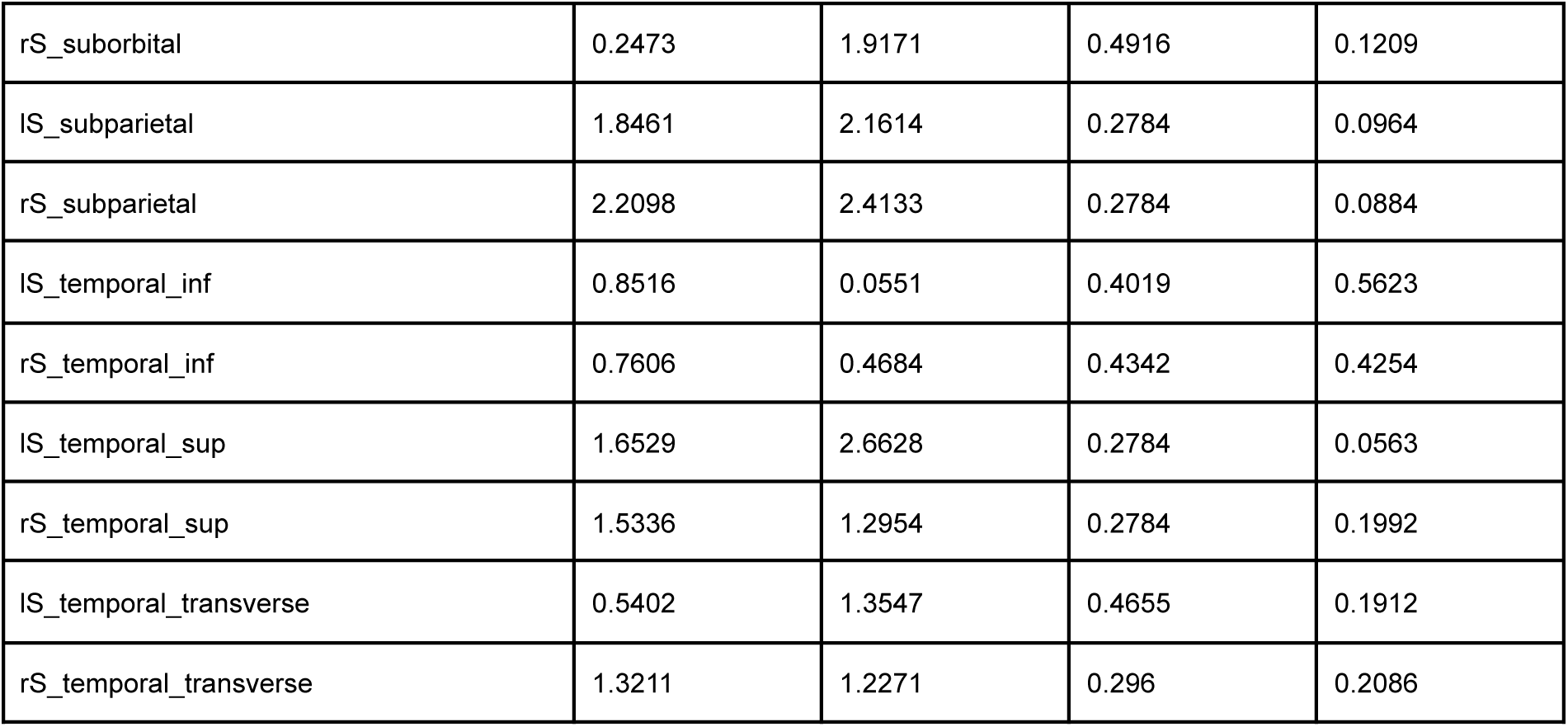

**Supplementary Material 6b.**
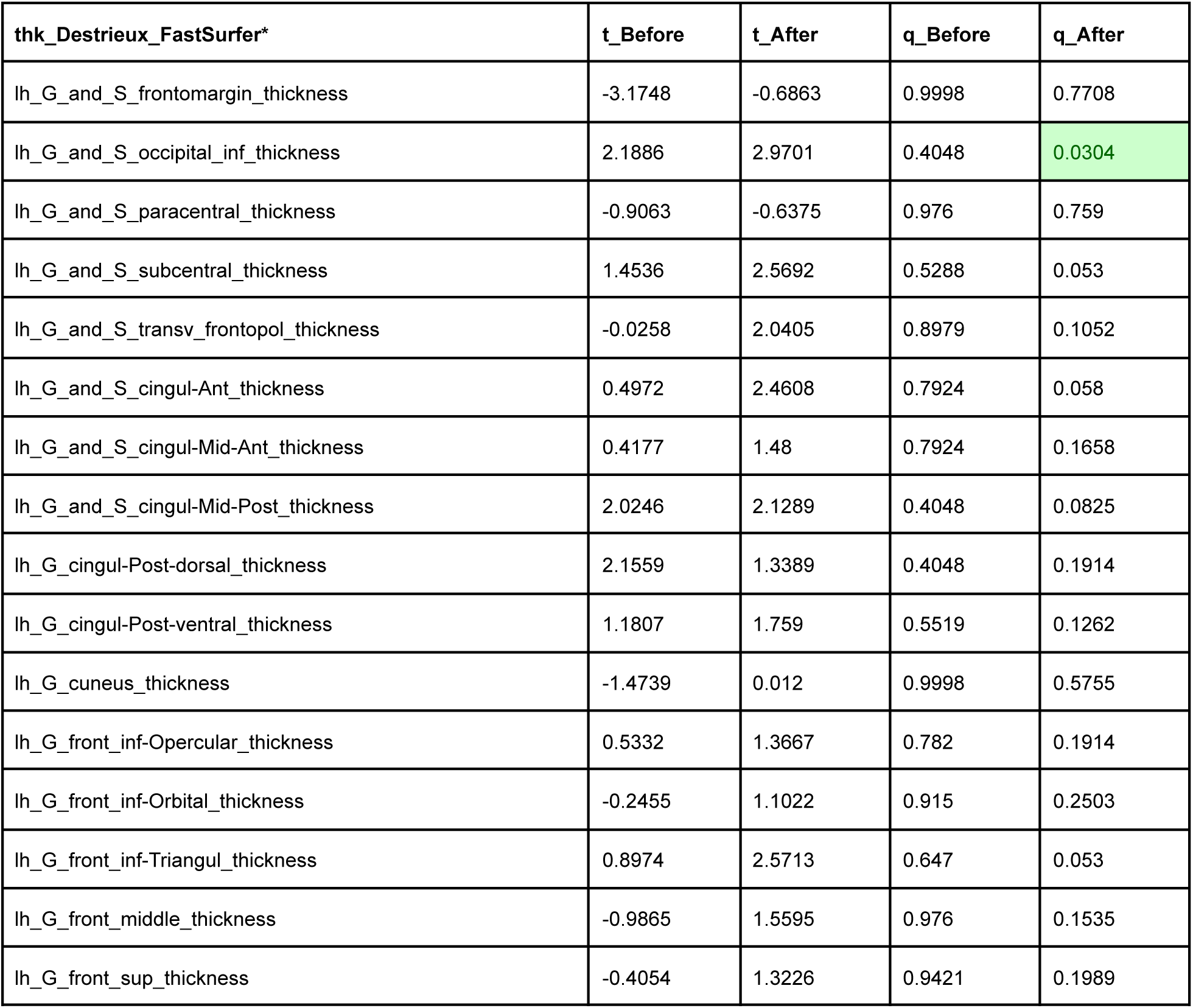
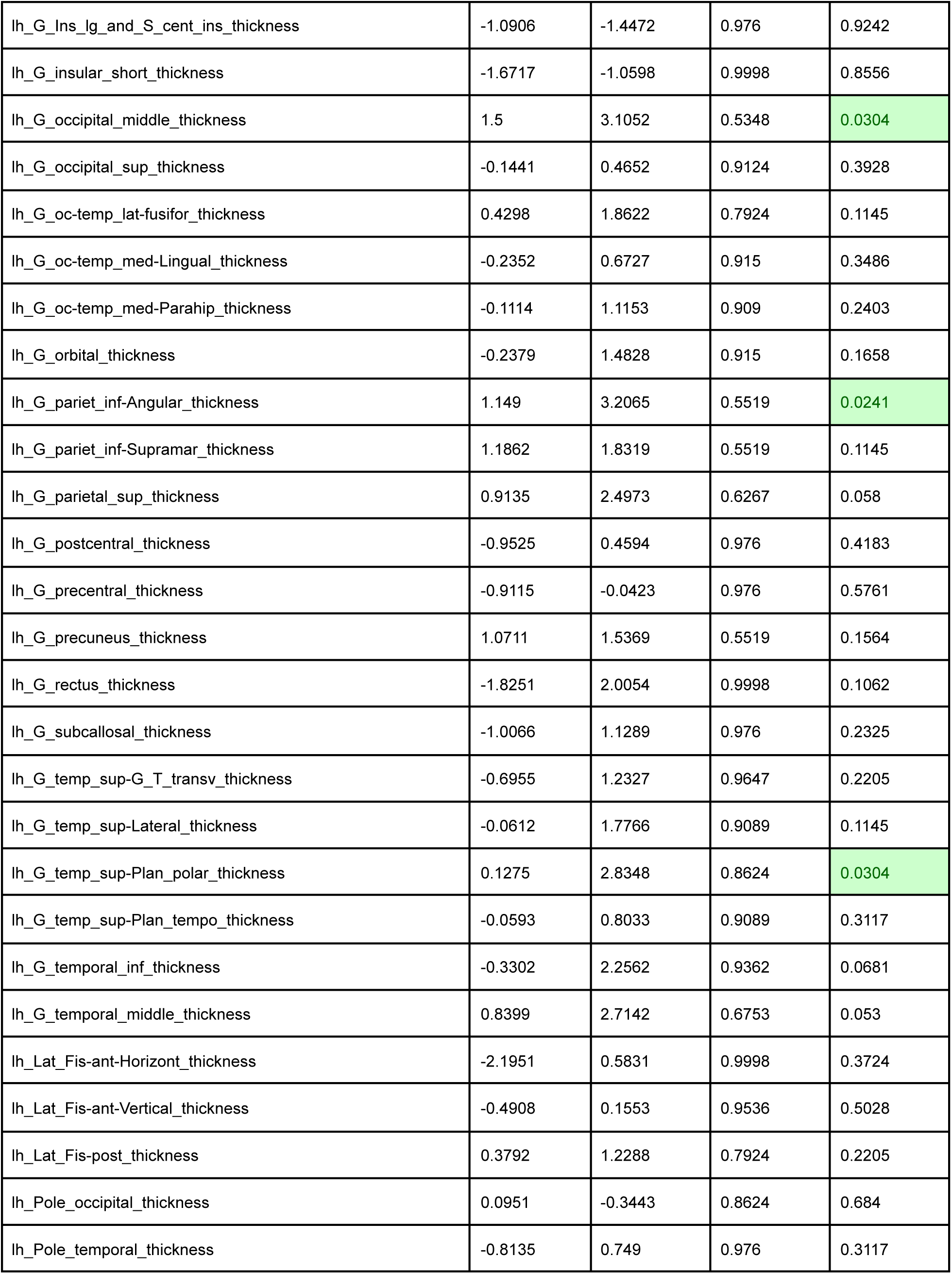
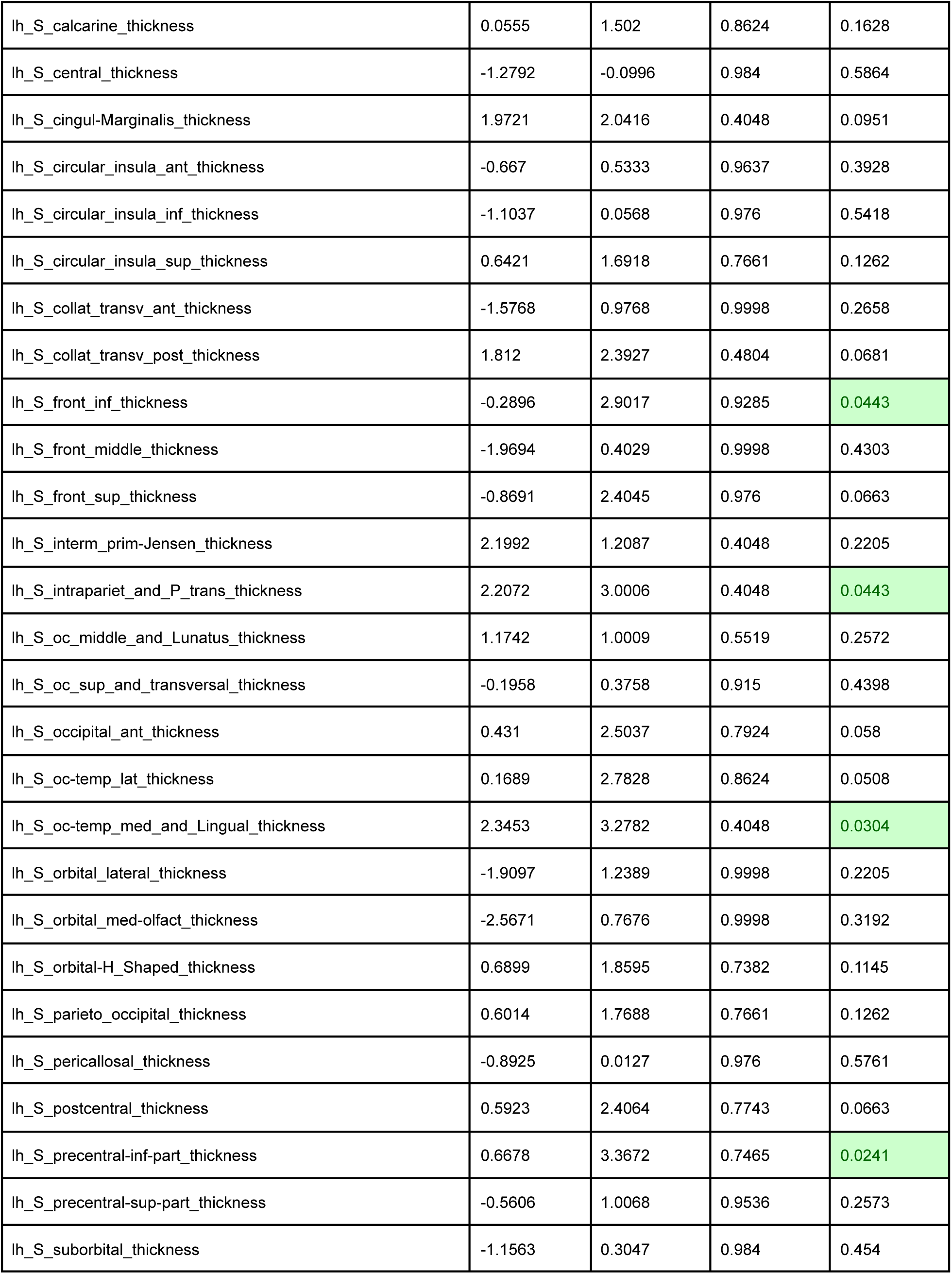
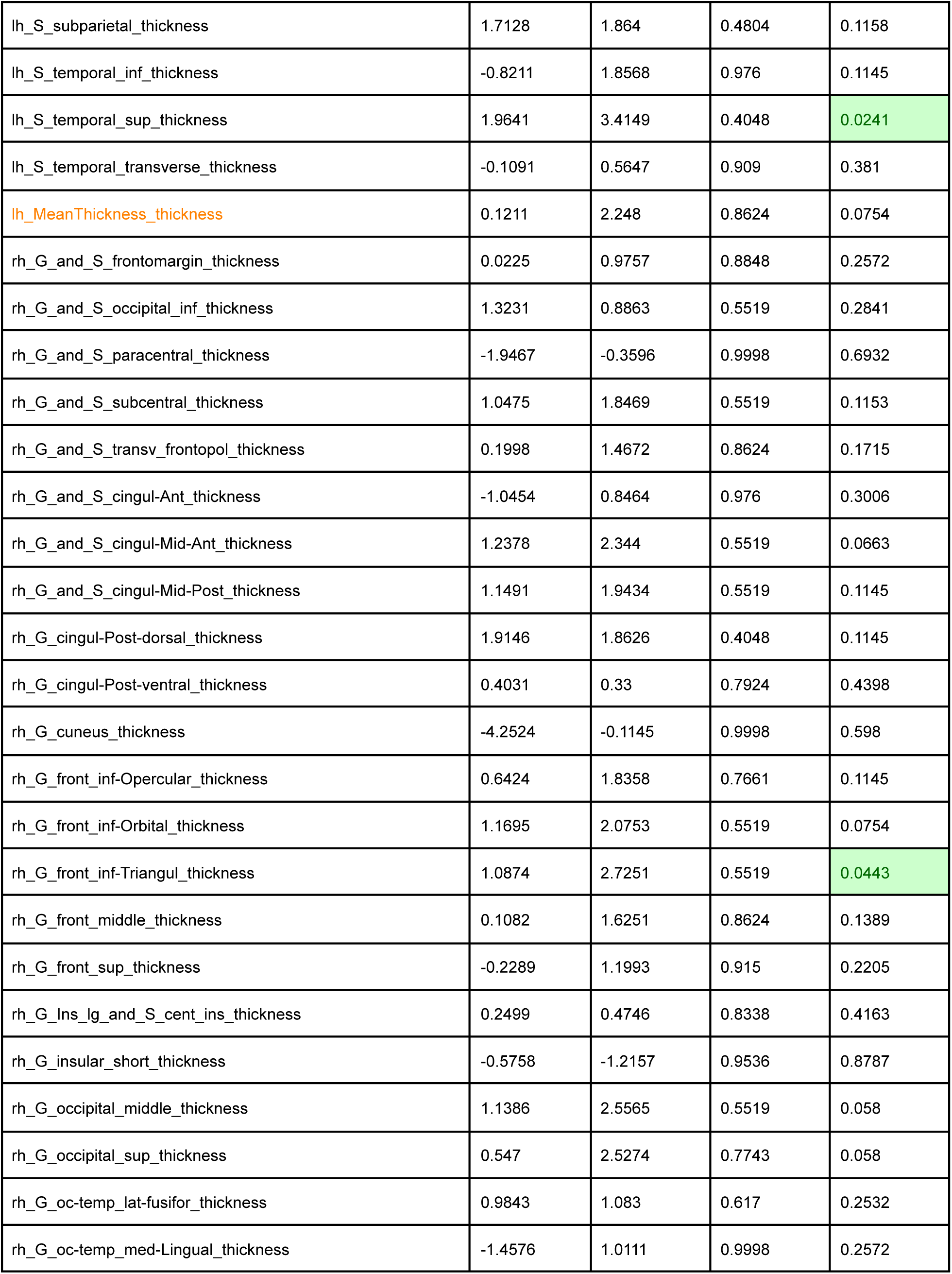
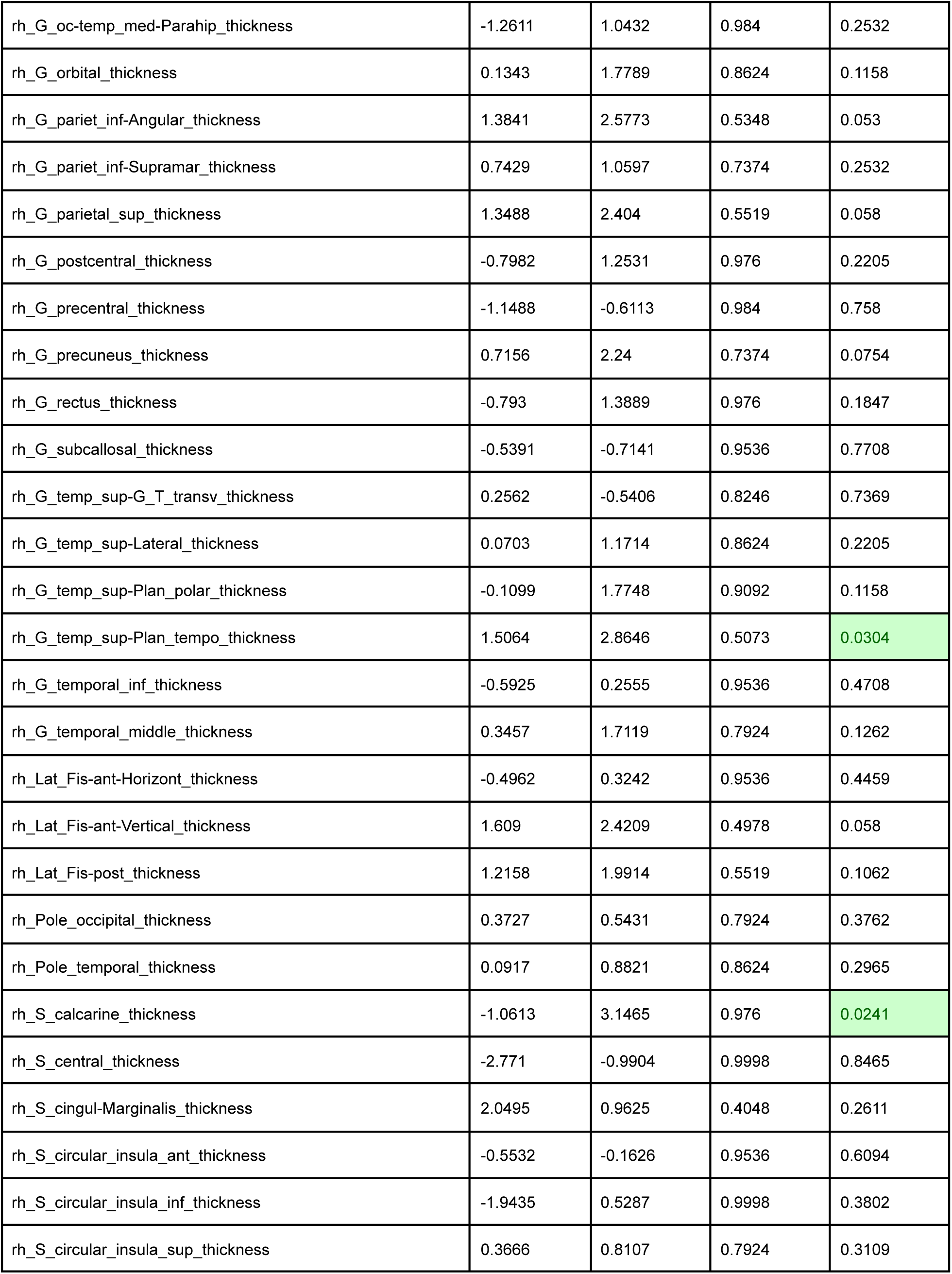
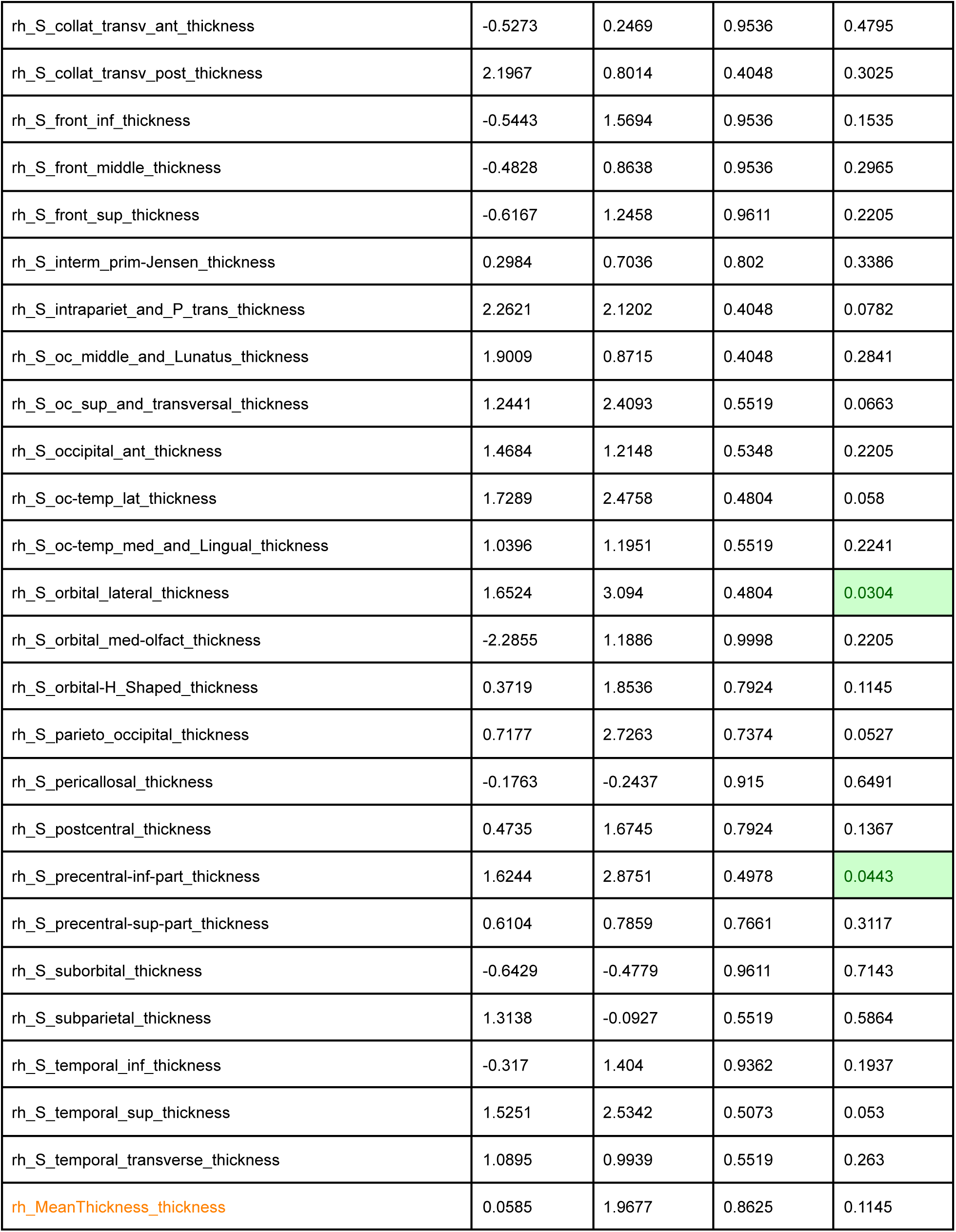

| thk_Destrieux_FastSurfer* | t_Before | t_After | q_Before | q_After |
| --- | --- | --- | --- | --- |
| lh_G_and_S_frontomargin_thickness | -3.1748 | -0.6863 | 0.9998 | 0.7708 |
| lh_G_and_S_occipital_inf_thickness | 2.1886 | 2.9701 | 0.4048 | 0.0304 |
| lh_G_and_S_paracentral_thickness | -0.9063 | -0.6375 | 0.976 | 0.759 |
| lh_G_and_S_subcentral_thickness | 1.4536 | 2.5692 | 0.5288 | 0.053 |
| lh_G_and_S_transv_frontopol_thickness | -0.0258 | 2.0405 | 0.8979 | 0.1052 |
| lh_G_and_S_cingul-Ant_thickness | 0.4972 | 2.4608 | 0.7924 | 0.058 |
| lh_G_and_S_cingul-Mid-Ant_thickness | 0.4177 | 1.48 | 0.7924 | 0.1658 |
| lh_G_and_S_cingul-Mid-Post_thickness | 2.0246 | 2.1289 | 0.4048 | 0.0825 |
| lh_G_cingul-Post-dorsal_thickness | 2.1559 | 1.3389 | 0.4048 | 0.1914 |
| lh_G_cingul-Post-ventral_thickness | 1.1807 | 1.759 | 0.5519 | 0.1262 |
| lh_G_cuneus_thickness | -1.4739 | 0.012 | 0.9998 | 0.5755 |
| lh_G_front_inf-Opercular_thickness | 0.5332 | 1.3667 | 0.782 | 0.1914 |
| lh_G_front_inf-Orbital_thickness | -0.2455 | 1.1022 | 0.915 | 0.2503 |
| lh_G_front_inf-Triangul_thickness | 0.8974 | 2.5713 | 0.647 | 0.053 |
| lh_G_front_middle_thickness | -0.9865 | 1.5595 | 0.976 | 0.1535 |
| lh_G_front_sup_thickness | -0.4054 | 1.3226 | 0.9421 | 0.1989 |
| lh_G_ins_lg_and_S_cent_ins_thickness | -1.0906 | -1.4472 | 0.976 | 0.9242 |
| lh_G_insular_short_thickness | -1.6717 | -1.0598 | 0.9998 | 0.8556 |
| lh_G_occipital_middle_thickness | 1.5 | 3.1052 | 0.5348 | 0.0304 |
| lh_G_occipital_sup_thickness | -0.1441 | 0.4652 | 0.9124 | 0.3928 |
| lh_G_oc-temp_lat-fusiform_thickness | 0.4298 | 1.8622 | 0.7924 | 0.1145 |
| lh_G_oc-temp_med-Lingual_thickness | -0.2352 | 0.6727 | 0.915 | 0.3486 |
| lh_G_oc-temp_med-Parahipp_thickness | -0.1114 | 1.1153 | 0.909 | 0.2403 |
| lh_G_orbital_thickness | -0.2379 | 1.4828 | 0.915 | 0.1658 |
| lh_G_pariet_inf-Angular_thickness | 1.149 | 3.2065 | 0.5519 | 0.0241 |
| lh_G_pariet_inf-Supramar_thickness | 1.1862 | 1.8319 | 0.5519 | 0.1145 |
| lh_G_parietal_sup_thickness | 0.9135 | 2.4973 | 0.6267 | 0.058 |
| lh_G_postcentral_thickness | -0.9525 | 0.4594 | 0.976 | 0.4183 |
| lh_G_precentral_thickness | -0.9115 | -0.0423 | 0.976 | 0.5761 |
| lh_G_precuneus_thickness | 1.0711 | 1.5369 | 0.5519 | 0.1564 |
| lh_G_rectus_thickness | -1.8251 | 2.0054 | 0.9998 | 0.1062 |
| lh_G_subcallosal_thickness | -1.0066 | 1.1289 | 0.976 | 0.2325 |
| lh_G_temp_sup-G_T_transv_thickness | -0.6955 | 1.2327 | 0.9647 | 0.2205 |
| lh_G_temp_sup-Lateral_thickness | -0.0612 | 1.7766 | 0.9089 | 0.1145 |
| lh_G_temp_sup-Plan_polar_thickness | 0.1275 | 2.8348 | 0.8624 | 0.0304 |
| lh_G_temp_sup-Plan_tempo_thickness | -0.0593 | 0.8033 | 0.9089 | 0.3117 |
| lh_G_temporal_inf_thickness | -0.3302 | 2.2562 | 0.9362 | 0.0681 |
| lh_G_temporal_middle_thickness | 0.8399 | 2.7142 | 0.6753 | 0.053 |
| lh_Lat_Fis-ant-Horizont_thickness | -2.1951 | 0.5831 | 0.9998 | 0.3724 |
| lh_Lat_Fis-ant-Vertical_thickness | -0.4908 | 0.1553 | 0.9536 | 0.5028 |
| lh_Lat_Fis-post_thickness | 0.3792 | 1.2288 | 0.7924 | 0.2205 |
| lh_Pole_occipital_thickness | 0.0951 | -0.3443 | 0.8624 | 0.684 |
| lh_Pole_temporal_thickness | -0.8135 | 0.749 | 0.976 | 0.3117 |
| lh_S_calcarine_thickness | 0.0555 | 1.502 | 0.8624 | 0.1628 |
| lh_S_central_thickness | -1.2792 | -0.0996 | 0.984 | 0.5864 |
| lh_S_cingul-Marginalis_thickness | 1.9721 | 2.0416 | 0.4048 | 0.0951 |
| lh_S_circular_insula_ant_thickness | -0.667 | 0.5333 | 0.9637 | 0.3928 |
| lh_S_circular_insula_inf_thickness | -1.1037 | 0.0568 | 0.976 | 0.5418 |
| lh_S_circular_insula_sup_thickness | 0.6421 | 1.6918 | 0.7661 | 0.1262 |
| lh_S_collat_transv_ant_thickness | -1.5768 | 0.9768 | 0.9998 | 0.2658 |
| lh_S_collat_transv_post_thickness | 1.812 | 2.3927 | 0.4804 | 0.0681 |
| lh_S_front_inf_thickness | -0.2896 | 2.9017 | 0.9285 | 0.0443 |
| lh_S_front_middle_thickness | -1.9694 | 0.4029 | 0.9998 | 0.4303 |
| lh_S_front_sup_thickness | -0.8691 | 2.4045 | 0.976 | 0.0663 |
| lh_S_interm_prim-Jensen_thickness | 2.1992 | 1.2087 | 0.4048 | 0.2205 |
| lh_S_intrapariet_and_P_trans_thickness | 2.2072 | 3.0006 | 0.4048 | 0.0443 |
| lh_S_oc_middle_and_Lunatus_thickness | 1.1742 | 1.0009 | 0.5519 | 0.2572 |
| lh_S_oc_sup_and_transversal_thickness | -0.1958 | 0.3758 | 0.915 | 0.4398 |
| lh_S_occipital_ant_thickness | 0.431 | 2.5037 | 0.7924 | 0.058 |
| lh_S_oc-temp_lat_thickness | 0.1689 | 2.7828 | 0.8624 | 0.0508 |
| lh_S_oc-temp_med_and_Lingual_thickness | 2.3453 | 3.2782 | 0.4048 | 0.0304 |
| lh_S_orbital_lateral_thickness | -1.9097 | 1.2389 | 0.9998 | 0.2205 |
| lh_S_orbital_med-olfact_thickness | -2.5671 | 0.7676 | 0.9998 | 0.3192 |
| lh_S_orbital-H_Shaped_thickness | 0.6899 | 1.8595 | 0.7382 | 0.1145 |
| lh_S_parieto_occipital_thickness | 0.6014 | 1.7688 | 0.7661 | 0.1262 |
| lh_S_pericallosal_thickness | -0.8925 | 0.0127 | 0.976 | 0.5761 |
| lh_S_postcentral_thickness | 0.5923 | 2.4064 | 0.7743 | 0.0663 |
| lh_S_precentral-inf-part_thickness | 0.6678 | 3.3672 | 0.7465 | 0.0241 |
| lh_S_precentral-sup-part_thickness | -0.5606 | 1.0068 | 0.9536 | 0.2573 |
| lh_S_suborbital_thickness | -1.1563 | 0.3047 | 0.984 | 0.454 |
| lh_S_subparietal_thickness | 1.7128 | 1.864 | 0.4804 | 0.1158 |
| lh_S_temporal_inf_thickness | -0.8211 | 1.8568 | 0.976 | 0.1145 |
| lh_S_temporal_sup_thickness | 1.9641 | 3.4149 | 0.4048 | 0.0241 |
| lh_S_temporal_transverse_thickness | -0.1091 | 0.5647 | 0.909 | 0.381 |
| lh_MeanThickness_thickness | 0.1211 | 2.248 | 0.8624 | 0.0754 |
| rh_G_and_S_frontomargin_thickness | 0.0225 | 0.9757 | 0.8848 | 0.2572 |
| rh_G_and_S_occipital_inf_thickness | 1.3231 | 0.8863 | 0.5519 | 0.2841 |
| rh_G_and_S_paracentral_thickness | -1.9467 | -0.3596 | 0.9998 | 0.6932 |
| rh_G_and_S_subcentral_thickness | 1.0475 | 1.8469 | 0.5519 | 0.1153 |
| rh_G_and_S_transv_frontopol_thickness | 0.1998 | 1.4672 | 0.8624 | 0.1715 |
| rh_G_and_S_cingul-Ant_thickness | -1.0454 | 0.8464 | 0.976 | 0.3006 |
| rh_G_and_S_cingul-Mid-Ant_thickness | 1.2378 | 2.344 | 0.5519 | 0.0663 |
| rh_G_and_S_cingul-Mid-Post_thickness | 1.1491 | 1.9434 | 0.5519 | 0.1145 |
| rh_G_cingul-Post-dorsal_thickness | 1.9146 | 1.8626 | 0.4048 | 0.1145 |
| rh_G_cingul-Post-ventral_thickness | 0.4031 | 0.33 | 0.7924 | 0.4398 |
| rh_G_cuneus_thickness | -4.2524 | -0.1145 | 0.9998 | 0.598 |
| rh_G_front_inf-Opercular_thickness | 0.6424 | 1.8358 | 0.7661 | 0.1145 |
| rh_G_front_inf-Orbital_thickness | 1.1695 | 2.0753 | 0.5519 | 0.0754 |
| rh_G_front_inf-Triangul_thickness | 1.0874 | 2.7251 | 0.5519 | 0.0443 |
| rh_G_front_middle_thickness | 0.1082 | 1.6251 | 0.8624 | 0.1389 |
| rh_G_front_sup_thickness | -0.2289 | 1.1993 | 0.915 | 0.2205 |
| rh_G_Ins_Ig_and_S_cent_ins_thickness | 0.2499 | 0.4746 | 0.8338 | 0.4163 |
| rh_G_insular_short_thickness | -0.5758 | -1.2157 | 0.9536 | 0.8787 |
| rh_G_occipital_middle_thickness | 1.1386 | 2.5565 | 0.5519 | 0.058 |
| rh_G_occipital_sup_thickness | 0.547 | 2.5274 | 0.7743 | 0.058 |
| rh_G_oc-temp_lat-fusifor_thickness | 0.9843 | 1.083 | 0.617 | 0.2532 |
| rh_G_oc-temp_med-Lingual_thickness | -1.4576 | 1.0111 | 0.9998 | 0.2572 |
| rh_G_oc-temp_med-Parahip_thickness | -1.2611 | 1.0432 | 0.984 | 0.2532 |
| rh_G_orbital_thickness | 0.1343 | 1.7789 | 0.8624 | 0.1158 |
| rh_G_pariet_inf-Angular_thickness | 1.3841 | 2.5773 | 0.5348 | 0.053 |
| rh_G_pariet_inf-Supramar_thickness | 0.7429 | 1.0597 | 0.7374 | 0.2532 |
| rh_G_parietal_sup_thickness | 1.3488 | 2.404 | 0.5519 | 0.058 |
| rh_G_postcentral_thickness | -0.7982 | 1.2531 | 0.976 | 0.2205 |
| rh_G_precentral_thickness | -1.1488 | -0.6113 | 0.984 | 0.758 |
| rh_G_precuneus_thickness | 0.7156 | 2.24 | 0.7374 | 0.0754 |
| rh_G_rectus_thickness | -0.793 | 1.3889 | 0.976 | 0.1847 |
| rh_G_subcallosal_thickness | -0.5391 | -0.7141 | 0.9536 | 0.7708 |
| rh_G_temp_sup-G_T_transv_thickness | 0.2562 | -0.5406 | 0.8246 | 0.7369 |
| rh_G_temp_sup-Lateral_thickness | 0.0703 | 1.1714 | 0.8624 | 0.2205 |
| rh_G_temp_sup-Plan_polar_thickness | -0.1099 | 1.7748 | 0.9092 | 0.1158 |
| rh_G_temp_sup-Plan_tempo_thickness | 1.5064 | 2.8646 | 0.5073 | 0.0304 |
| rh_G_temporal_inf_thickness | -0.5925 | 0.2555 | 0.9536 | 0.4708 |
| rh_G_temporal_middle_thickness | 0.3457 | 1.7119 | 0.7924 | 0.1262 |
| rh_Lat_Fis-ant-Horizont_thickness | -0.4962 | 0.3242 | 0.9536 | 0.4459 |
| rh_Lat_Fis-ant-Vertical_thickness | 1.609 | 2.4209 | 0.4978 | 0.058 |
| rh_Lat_Fis-post_thickness | 1.2158 | 1.9914 | 0.5519 | 0.1062 |
| rh_Pole_occipital_thickness | 0.3727 | 0.5431 | 0.7924 | 0.3762 |
| rh_Pole_temporal_thickness | 0.0917 | 0.8821 | 0.8624 | 0.2965 |
| rh_S_calcarine_thickness | -1.0613 | 3.1465 | 0.976 | 0.0241 |
| rh_S_central_thickness | -2.771 | -0.9904 | 0.9998 | 0.8465 |
| rh_S_cingul-Marginalis_thickness | 2.0495 | 0.9625 | 0.4048 | 0.2611 |
| rh_S_circular_insula_ant_thickness | -0.5532 | -0.1626 | 0.9536 | 0.6094 |
| rh_S_circular_insula_inf_thickness | -1.9435 | 0.5287 | 0.9998 | 0.3802 |
| rh_S_circular_insula_sup_thickness | 0.3666 | 0.8107 | 0.7924 | 0.3109 |
| rh_S_collat_transv_ant_thickness | -0.5273 | 0.2469 | 0.9536 | 0.4795 |
| rh_S_collat_transv_post_thickness | 2.1967 | 0.8014 | 0.4048 | 0.3025 |
| rh_S_front_inf_thickness | -0.5443 | 1.5694 | 0.9536 | 0.1535 |
| rh_S_front_middle_thickness | -0.4828 | 0.8638 | 0.9536 | 0.2965 |
| rh_S_front_sup_thickness | -0.6167 | 1.2458 | 0.9611 | 0.2205 |
| rh_S_interm_prim-Jensen_thickness | 0.2984 | 0.7036 | 0.802 | 0.3386 |
| rh_S_intrapariet_and_P_trans_thickness | 2.2621 | 2.1202 | 0.4048 | 0.0782 |
| rh_S_oc_middle_and_Lunatus_thickness | 1.9009 | 0.8715 | 0.4048 | 0.2841 |
| rh_S_oc_sup_and_transversal_thickness | 1.2441 | 2.4093 | 0.5519 | 0.0663 |
| rh_S_occipital_ant_thickness | 1.4684 | 1.2148 | 0.5348 | 0.2205 |
| rh_S_oc-temp_lat_thickness | 1.7289 | 2.4758 | 0.4804 | 0.058 |
| rh_S_oc-temp_med_and_Lingual_thickness | 1.0396 | 1.1951 | 0.5519 | 0.2241 |
| rh_S_orbital_lateral_thickness | 1.6524 | 3.094 | 0.4804 | 0.0304 |
| rh_S_orbital_med-olfact_thickness | -2.2855 | 1.1886 | 0.9998 | 0.2205 |
| rh_S_orbital-H_Shaped_thickness | 0.3719 | 1.8536 | 0.7924 | 0.1145 |
| rh_S_parieto_occipital_thickness | 0.7177 | 2.7263 | 0.7374 | 0.0527 |
| rh_S_pericallosal_thickness | -0.1763 | -0.2437 | 0.915 | 0.6491 |
| rh_S_postcentral_thickness | 0.4735 | 1.6745 | 0.7924 | 0.1367 |
| rh_S_precentral-inf-part_thickness | 1.6244 | 2.8751 | 0.4978 | 0.0443 |
| rh_S_precentral-sup-part_thickness | 0.6104 | 0.7859 | 0.7661 | 0.3117 |
| rh_S_suborbital_thickness | -0.6429 | -0.4779 | 0.9611 | 0.7143 |
| rh_S_subparietal_thickness | 1.3138 | -0.0927 | 0.5519 | 0.5864 |
| rh_S_temporal_inf_thickness | -0.317 | 1.404 | 0.9362 | 0.1937 |
| rh_S_temporal_sup_thickness | 1.5251 | 2.5342 | 0.5073 | 0.053 |
| rh_S_temporal_transverse_thickness | 1.0895 | 0.9939 | 0.5519 | 0.263 |
| rh_MeanThickness_thickness | 0.0585 | 1.9677 | 0.8625 | 0.1145 |

**Supplementary Material 6c.**
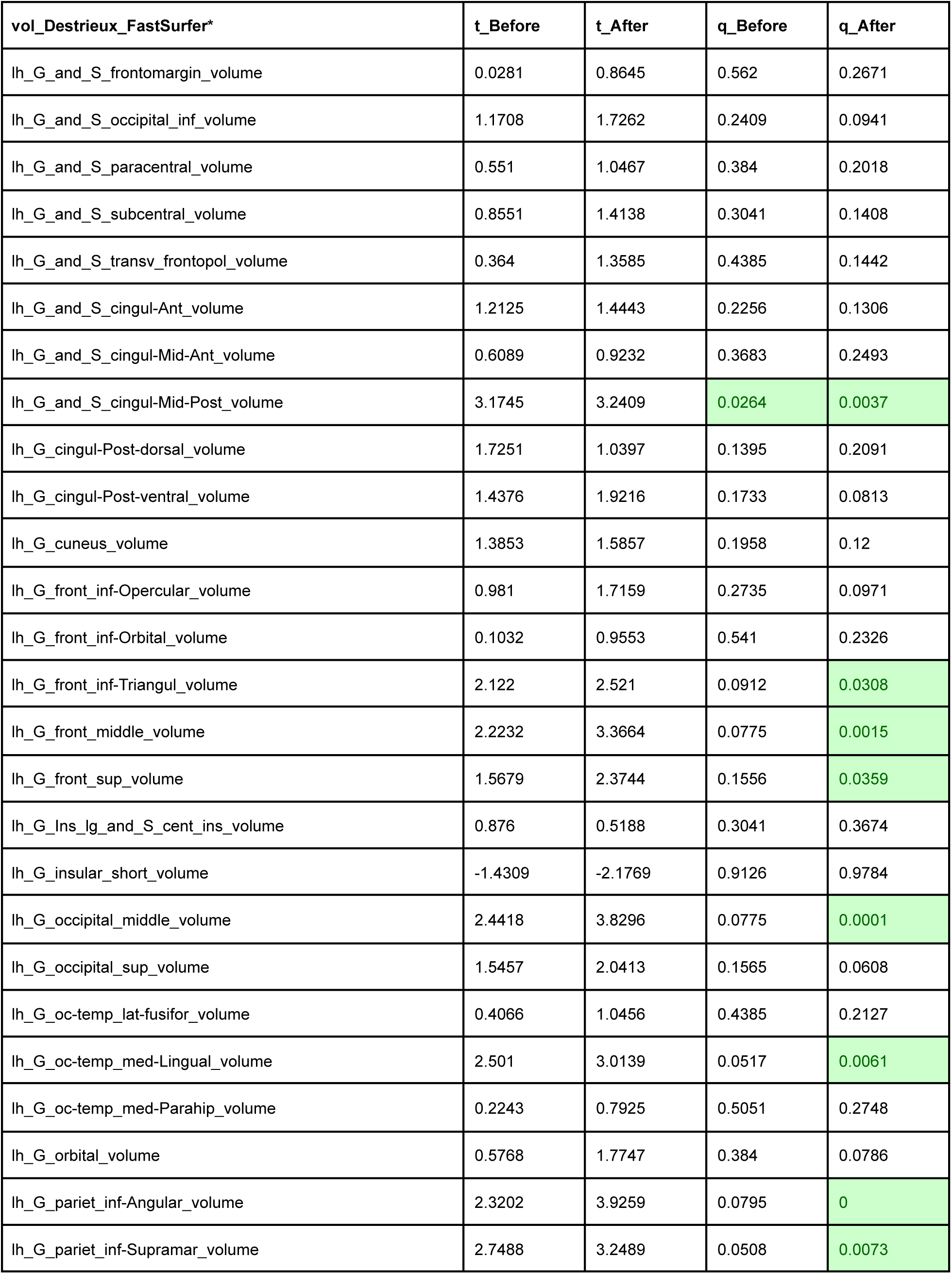
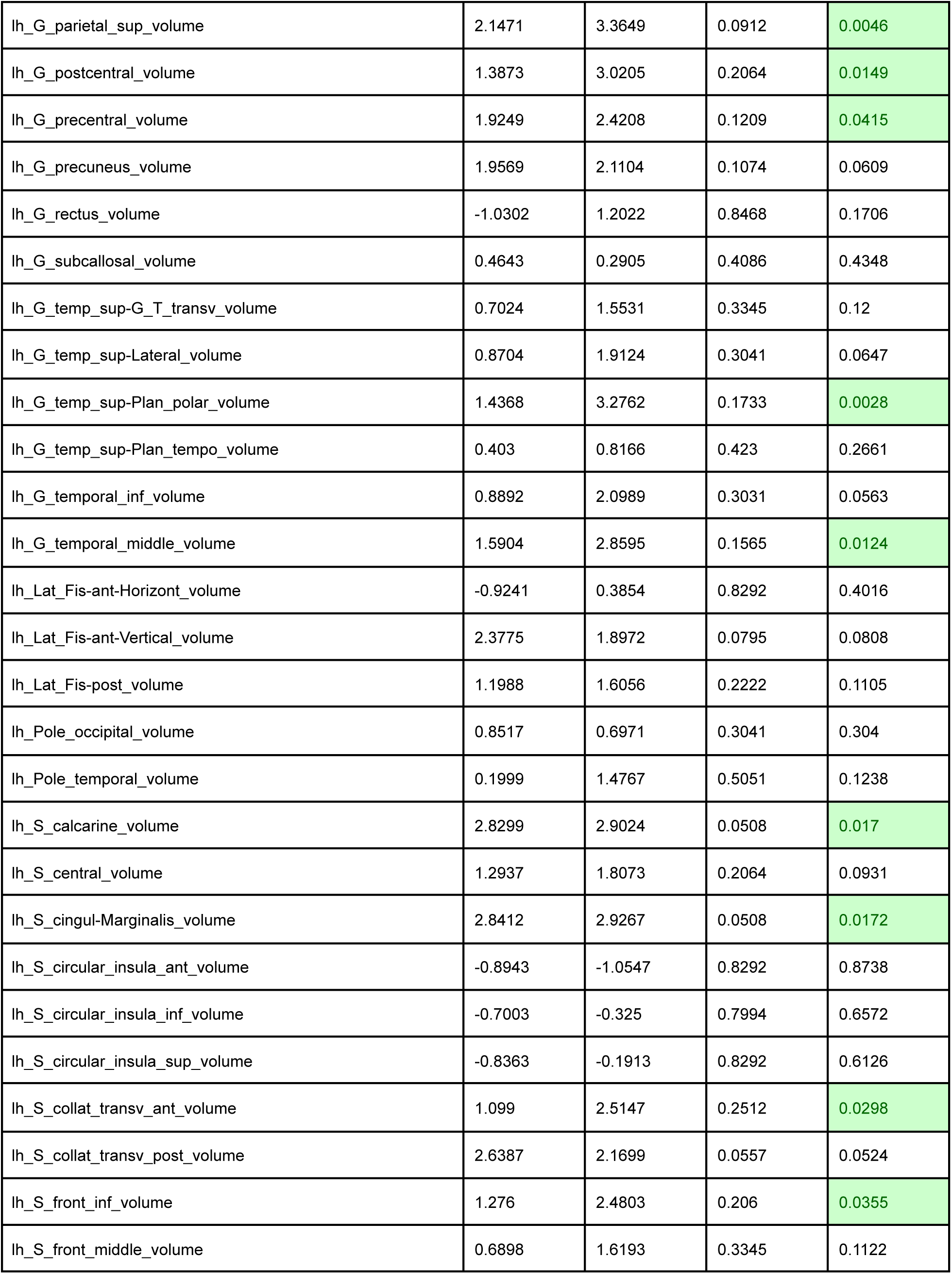
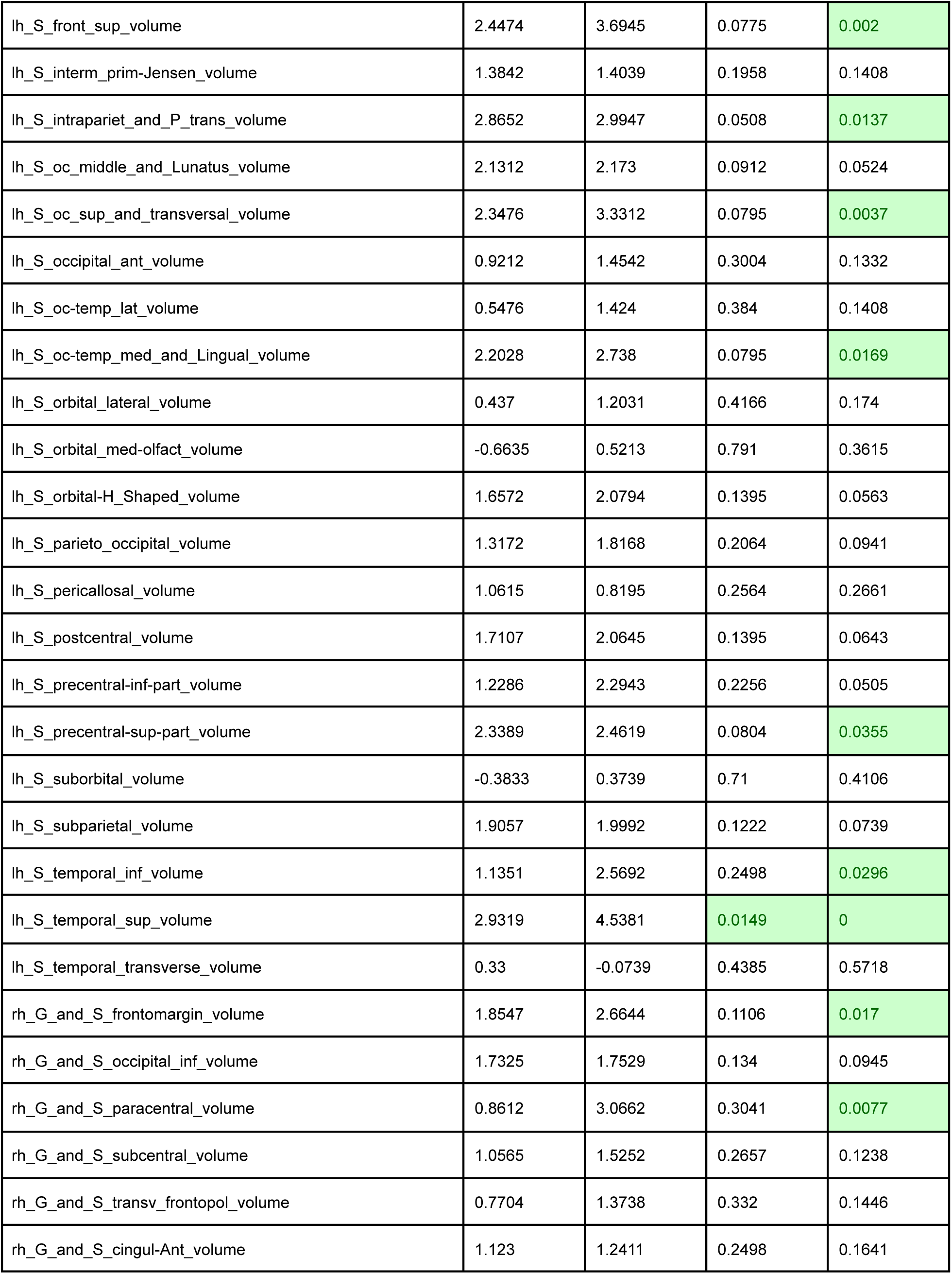
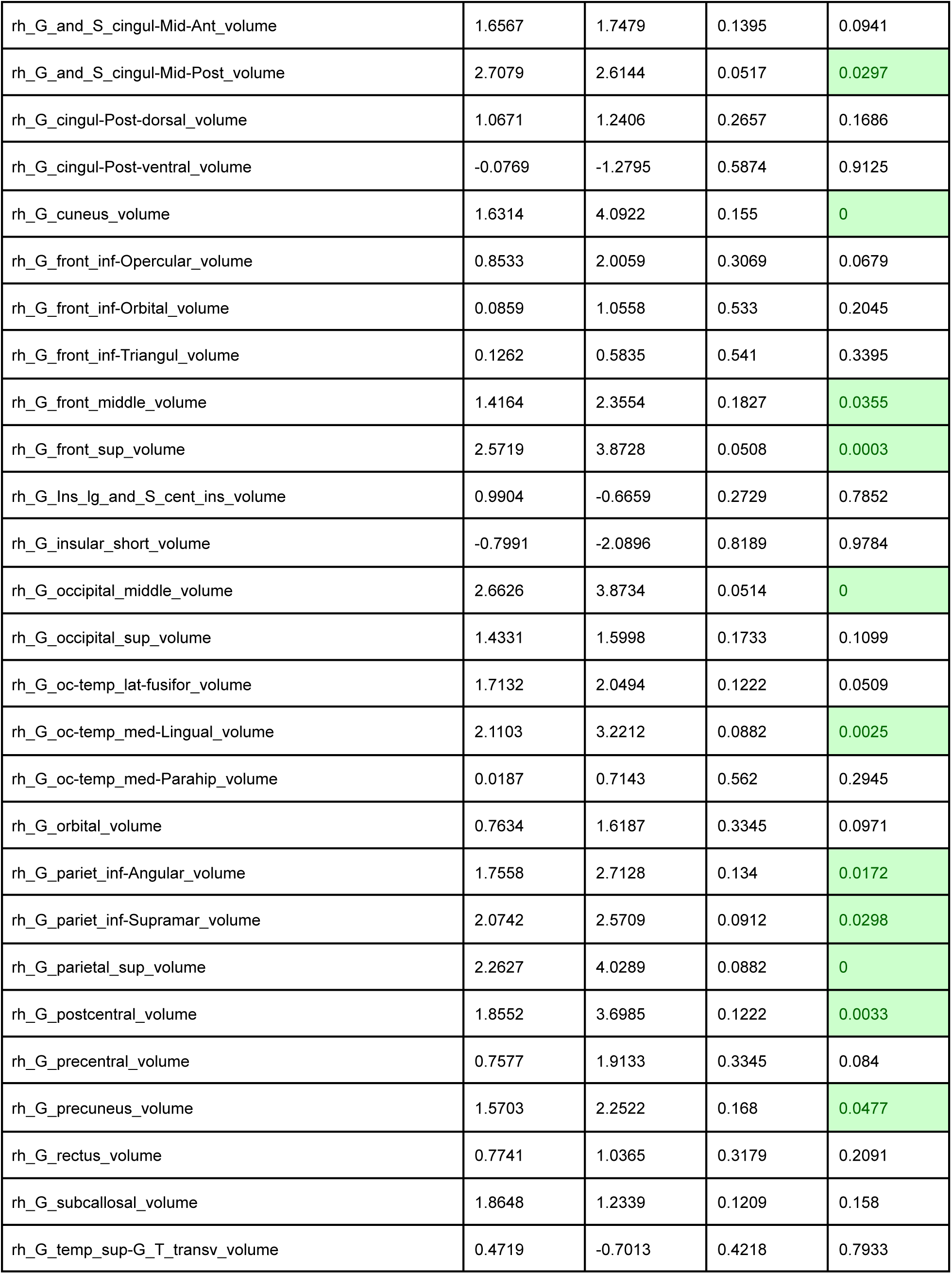
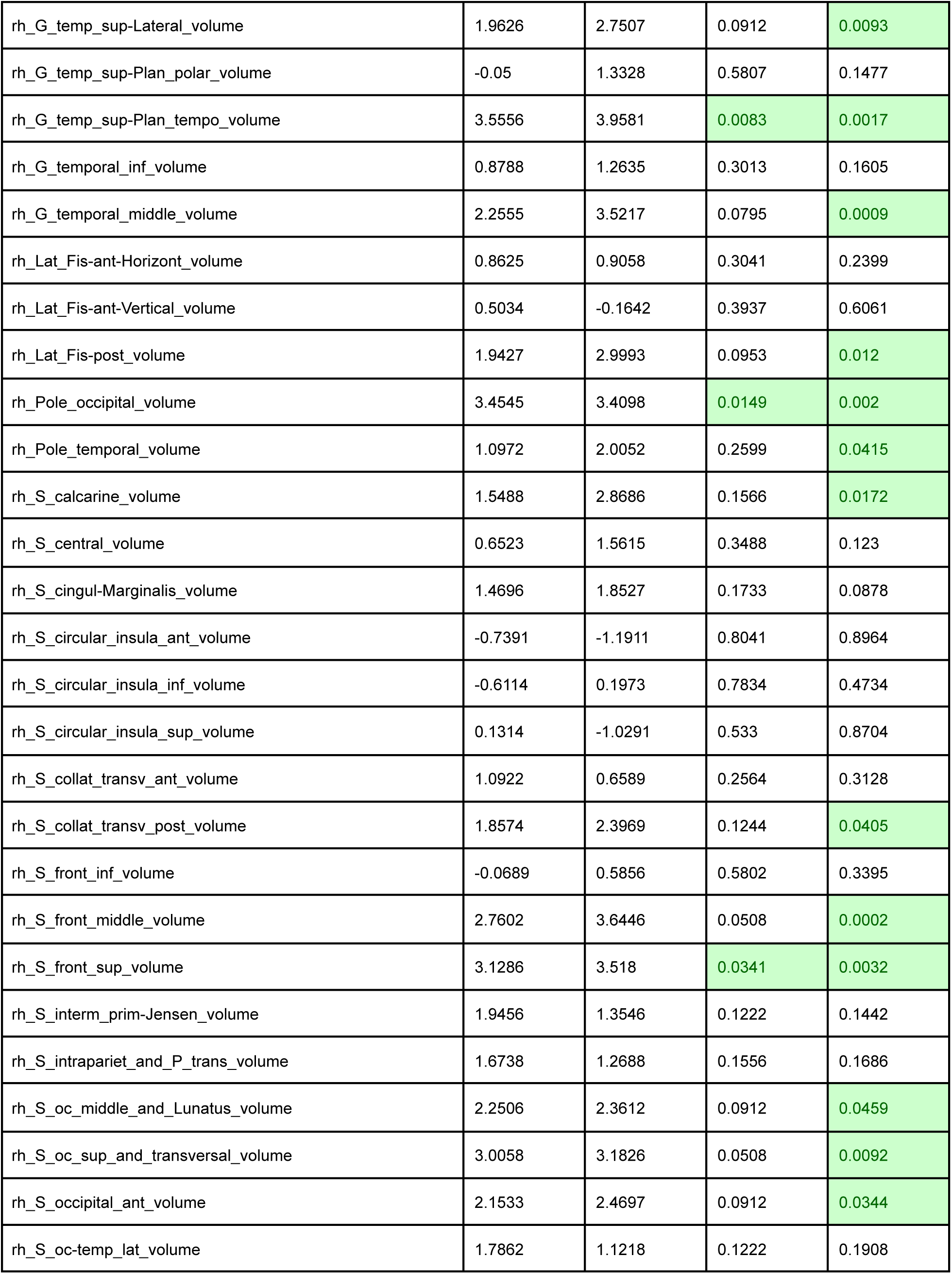
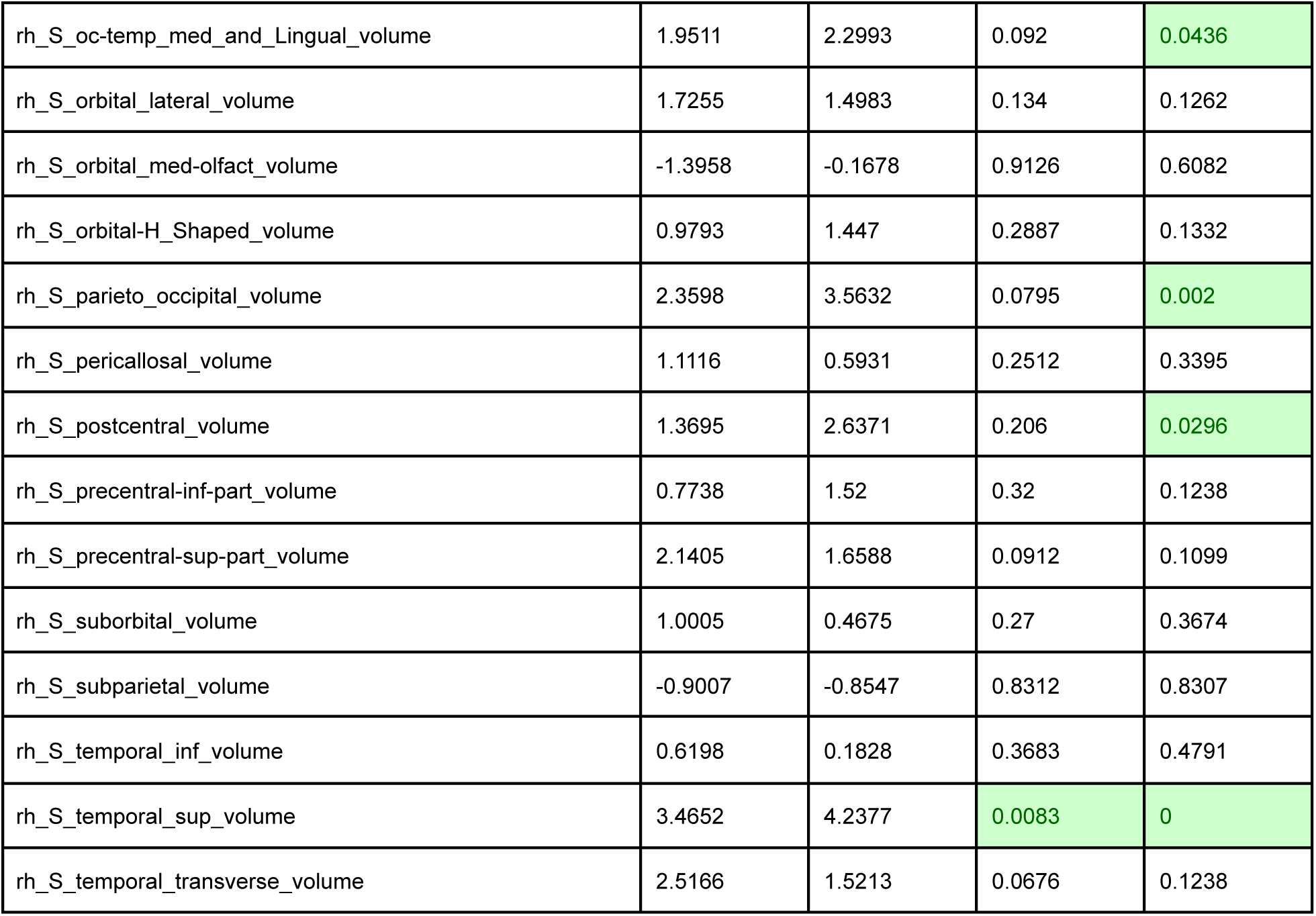

| **vol_Destrieux_FastSurfer\*** | **t_Before** | **t_After** | **q_Before** | **q_After** |
| --- | --- | --- | --- | --- |
| lh_G_and_S_frontomargin_volume | 0.0281 | 0.8645 | 0.562 | 0.2671 |
| lh_G_and_S_occipital_inf_volume | 1.1708 | 1.7262 | 0.2409 | 0.0941 |
| lh_G_and_S_paracentral_volume | 0.551 | 1.0467 | 0.384 | 0.2018 |
| lh_G_and_S_subcentral_volume | 0.8551 | 1.4138 | 0.3041 | 0.1408 |
| lh_G_and_S_transv_frontopol_volume | 0.364 | 1.3585 | 0.4385 | 0.1442 |
| lh_G_and_S_cingul-Ant_volume | 1.2125 | 1.4443 | 0.2256 | 0.1306 |
| lh_G_and_S_cingul-Mid-Ant_volume | 0.6089 | 0.9232 | 0.3683 | 0.2493 |
| lh_G_and_S_cingul-Mid-Post_volume | 3.1745 | 3.2409 | 0.0264 | 0.0037 |
| lh_G_cingul-Post-dorsal_volume | 1.7251 | 1.0397 | 0.1395 | 0.2091 |
| lh_G_cingul-Post-ventral_volume | 1.4376 | 1.9216 | 0.1733 | 0.0813 |
| lh_G_cuneus_volume | 1.3853 | 1.5857 | 0.1958 | 0.12 |
| lh_G_front_inf-Opercular_volume | 0.981 | 1.7159 | 0.2735 | 0.0971 |
| lh_G_front_inf-Orbital_volume | 0.1032 | 0.9553 | 0.541 | 0.2326 |
| lh_G_front_inf-Triangul_volume | 2.122 | 2.521 | 0.0912 | 0.0308 |
| lh_G_front_middle_volume | 2.2232 | 3.3664 | 0.0775 | 0.0015 |
| lh_G_front_sup_volume | 1.5679 | 2.3744 | 0.1556 | 0.0359 |
| lh_G_Ins_lg_and_S_cent_ins_volume | 0.876 | 0.5188 | 0.3041 | 0.3674 |
| lh_G_insular_short_volume | -1.4309 | -2.1769 | 0.9126 | 0.9784 |
| lh_G_occipital_middle_volume | 2.4418 | 3.8296 | 0.0775 | 0.0001 |
| lh_G_occipital_sup_volume | 1.5457 | 2.0413 | 0.1565 | 0.0608 |
| lh_G_oc-temp_lat-fusifor_volume | 0.4066 | 1.0456 | 0.4385 | 0.2127 |
| lh_G_oc-temp_med-Lingual_volume | 2.501 | 3.0139 | 0.0517 | 0.0061 |
| lh_G_oc-temp_med-Parahip_volume | 0.2243 | 0.7925 | 0.5051 | 0.2748 |
| lh_G_orbital_volume | 0.5768 | 1.7747 | 0.384 | 0.0786 |
| lh_G_pariet_inf-Angular_volume | 2.3202 | 3.9259 | 0.0795 | 0 |
| lh_G_pariet_inf-Supramar_volume | 2.7488 | 3.2489 | 0.0508 | 0.0073 |
| lh_G_parietal_sup_volume | 2.1471 | 3.3649 | 0.0912 | 0.0046 |
| lh_G_postcentral_volume | 1.3873 | 3.0205 | 0.2064 | 0.0149 |
| lh_G_precentral_volume | 1.9249 | 2.4208 | 0.1209 | 0.0415 |
| lh_G_precuneus_volume | 1.9569 | 2.1104 | 0.1074 | 0.0609 |
| lh_G_rectus_volume | -1.0302 | 1.2022 | 0.8468 | 0.1706 |
| lh_G_subcallosal_volume | 0.4643 | 0.2905 | 0.4086 | 0.4348 |
| lh_G_temp_sup-G_T_transv_volume | 0.7024 | 1.5531 | 0.3345 | 0.12 |
| lh_G_temp_sup-Lateral_volume | 0.8704 | 1.9124 | 0.3041 | 0.0647 |
| lh_G_temp_sup-Plan_polar_volume | 1.4368 | 3.2762 | 0.1733 | 0.0028 |
| lh_G_temp_sup-Plan_tempo_volume | 0.403 | 0.8166 | 0.423 | 0.2661 |
| lh_G_temporal_inf_volume | 0.8892 | 2.0989 | 0.3031 | 0.0563 |
| lh_G_temporal_middle_volume | 1.5904 | 2.8595 | 0.1565 | 0.0124 |
| lh_Lat_Fis-ant-Horizont_volume | -0.9241 | 0.3854 | 0.8292 | 0.4016 |
| lh_Lat_Fis-ant-Vertical_volume | 2.3775 | 1.8972 | 0.0795 | 0.0808 |
| lh_Lat_Fis-post_volume | 1.1988 | 1.6056 | 0.2222 | 0.1105 |
| lh_Pole_occipital_volume | 0.8517 | 0.6971 | 0.3041 | 0.304 |
| lh_Pole_temporal_volume | 0.1999 | 1.4767 | 0.5051 | 0.1238 |
| lh_S_calcarine_volume | 2.8299 | 2.9024 | 0.0508 | 0.017 |
| lh_S_central_volume | 1.2937 | 1.8073 | 0.2064 | 0.0931 |
| lh_S_cingul-Marginalis_volume | 2.8412 | 2.9267 | 0.0508 | 0.0172 |
| lh_S_circular_insula_ant_volume | -0.8943 | -1.0547 | 0.8292 | 0.8738 |
| lh_S_circular_insula_inf_volume | -0.7003 | -0.325 | 0.7994 | 0.6572 |
| lh_S_circular_insula_sup_volume | -0.8363 | -0.1913 | 0.8292 | 0.6126 |
| lh_S_collat_transv_ant_volume | 1.099 | 2.5147 | 0.2512 | 0.0298 |
| lh_S_collat_transv_post_volume | 2.6387 | 2.1699 | 0.0557 | 0.0524 |
| lh_S_front_inf_volume | 1.276 | 2.4803 | 0.206 | 0.0355 |
| lh_S_front_middle_volume | 0.6898 | 1.6193 | 0.3345 | 0.1122 |
| lh_S_front_sup_volume | 2.4474 | 3.6945 | 0.0775 | 0.002 |
| lh_S_interm_prim-Jensen_volume | 1.3842 | 1.4039 | 0.1958 | 0.1408 |
| lh_S_intrapariet_and_P_trans_volume | 2.8652 | 2.9947 | 0.0508 | 0.0137 |
| lh_S_oc_middle_and_Lunatus_volume | 2.1312 | 2.173 | 0.0912 | 0.0524 |
| lh_S_oc_sup_and_transversal_volume | 2.3476 | 3.3312 | 0.0795 | 0.0037 |
| lh_S_occipital_ant_volume | 0.9212 | 1.4542 | 0.3004 | 0.1332 |
| lh_S_oc-temp_lat_volume | 0.5476 | 1.424 | 0.384 | 0.1408 |
| lh_S_oc-temp_med_and_Lingual_volume | 2.2028 | 2.738 | 0.0795 | 0.0169 |
| lh_S_orbital_lateral_volume | 0.437 | 1.2031 | 0.4166 | 0.174 |
| lh_S_orbital_med-olfact_volume | -0.6635 | 0.5213 | 0.791 | 0.3615 |
| lh_S_orbital-H_Shaped_volume | 1.6572 | 2.0794 | 0.1395 | 0.0563 |
| lh_S_parieto_occipital_volume | 1.3172 | 1.8168 | 0.2064 | 0.0941 |
| lh_S_pericallosal_volume | 1.0615 | 0.8195 | 0.2564 | 0.2661 |
| lh_S_postcentral_volume | 1.7107 | 2.0645 | 0.1395 | 0.0643 |
| lh_S_precentral-inf-part_volume | 1.2286 | 2.2943 | 0.2256 | 0.0505 |
| lh_S_precentral-sup-part_volume | 2.3389 | 2.4619 | 0.0804 | 0.0355 |
| lh_S_suborbital_volume | -0.3833 | 0.3739 | 0.71 | 0.4106 |
| lh_S_subparietal_volume | 1.9057 | 1.9992 | 0.1222 | 0.0739 |
| lh_S_temporal_inf_volume | 1.1351 | 2.5692 | 0.2498 | 0.0296 |
| lh_S_temporal_sup_volume | 2.9319 | 4.5381 | 0.0149 | 0 |
| lh_S_temporal_transverse_volume | 0.33 | -0.0739 | 0.4385 | 0.5718 |
| rh_G_and_S_frontomargin_volume | 1.8547 | 2.6644 | 0.1106 | 0.017 |
| rh_G_and_S_occipital_inf_volume | 1.7325 | 1.7529 | 0.134 | 0.0945 |
| rh_G_and_S_paracentral_volume | 0.8612 | 3.0662 | 0.3041 | 0.0077 |
| rh_G_and_S_subcentral_volume | 1.0565 | 1.5252 | 0.2657 | 0.1238 |
| rh_G_and_S_transv_frontopol_volume | 0.7704 | 1.3738 | 0.332 | 0.1446 |
| rh_G_and_S_cingul-Ant_volume | 1.123 | 1.2411 | 0.2498 | 0.1641 |
| rh_G_and_S_cingul-Mid-Ant_volume | 1.6567 | 1.7479 | 0.1395 | 0.0941 |
| rh_G_and_S_cingul-Mid-Post_volume | 2.7079 | 2.6144 | 0.0517 | 0.0297 |
| rh_G_cingul-Post-dorsal_volume | 1.0671 | 1.2406 | 0.2657 | 0.1686 |
| rh_G_cingul-Post-ventral_volume | -0.0769 | -1.2795 | 0.5874 | 0.9125 |
| rh_G_cuneus_volume | 1.6314 | 4.0922 | 0.155 | 0 |
| rh_G_front_inf-Opercular_volume | 0.8533 | 2.0059 | 0.3069 | 0.0679 |
| rh_G_front_inf-Orbital_volume | 0.0859 | 1.0558 | 0.533 | 0.2045 |
| rh_G_front_inf-Triangul_volume | 0.1262 | 0.5835 | 0.541 | 0.3395 |
| rh_G_front_middle_volume | 1.4164 | 2.3554 | 0.1827 | 0.0355 |
| rh_G_front_sup_volume | 2.5719 | 3.8728 | 0.0508 | 0.0003 |
| rh_G_Ins_lg_and_S_cent_ins_volume | 0.9904 | -0.6659 | 0.2729 | 0.7852 |
| rh_G_insular_short_volume | -0.7991 | -2.0896 | 0.8189 | 0.9784 |
| rh_G_occipital_middle_volume | 2.6626 | 3.8734 | 0.0514 | 0 |
| rh_G_occipital_sup_volume | 1.4331 | 1.5998 | 0.1733 | 0.1099 |
| rh_G_oc-temp_lat-fusiform_volume | 1.7132 | 2.0494 | 0.1222 | 0.0509 |
| rh_G_oc-temp_med-Lingual_volume | 2.1103 | 3.2212 | 0.0882 | 0.0025 |
| rh_G_oc-temp_med-Parahip_volume | 0.0187 | 0.7143 | 0.562 | 0.2945 |
| rh_G_orbital_volume | 0.7634 | 1.6187 | 0.3345 | 0.0971 |
| rh_G_pariet_inf-Angular_volume | 1.7558 | 2.7128 | 0.134 | 0.0172 |
| rh_G_pariet_inf-Supramar_volume | 2.0742 | 2.5709 | 0.0912 | 0.0298 |
| rh_G_parietal_sup_volume | 2.2627 | 4.0289 | 0.0882 | 0 |
| rh_G_postcentral_volume | 1.8552 | 3.6985 | 0.1222 | 0.0033 |
| rh_G_precentral_volume | 0.7577 | 1.9133 | 0.3345 | 0.084 |
| rh_G_precuneus_volume | 1.5703 | 2.2522 | 0.168 | 0.0477 |
| rh_G_rectus_volume | 0.7741 | 1.0365 | 0.3179 | 0.2091 |
| rh_G_subcallosal_volume | 1.8648 | 1.2339 | 0.1209 | 0.158 |
| rh_G_temp_sup-G_T_transv_volume | 0.4719 | -0.7013 | 0.4218 | 0.7933 |
| rh_G_temp_sup-Lateral_volume | 1.9626 | 2.7507 | 0.0912 | 0.0093 |
| rh_G_temp_sup-Plan_polar_volume | -0.05 | 1.3328 | 0.5807 | 0.1477 |
| rh_G_temp_sup-Plan_tempo_volume | 3.5556 | 3.9581 | 0.0083 | 0.0017 |
| rh_G_temporal_inf_volume | 0.8788 | 1.2635 | 0.3013 | 0.1605 |
| rh_G_temporal_middle_volume | 2.2555 | 3.5217 | 0.0795 | 0.0009 |
| rh_Lat_Fis-ant-Horizont_volume | 0.8625 | 0.9058 | 0.3041 | 0.2399 |
| rh_Lat_Fis-ant-Vertical_volume | 0.5034 | -0.1642 | 0.3937 | 0.6061 |
| rh_Lat_Fis-post_volume | 1.9427 | 2.9993 | 0.0953 | 0.012 |
| rh_Pole_occipital_volume | 3.4545 | 3.4098 | 0.0149 | 0.002 |
| rh_Pole_temporal_volume | 1.0972 | 2.0052 | 0.2599 | 0.0415 |
| rh_S_calcarine_volume | 1.5488 | 2.8686 | 0.1566 | 0.0172 |
| rh_S_central_volume | 0.6523 | 1.5615 | 0.3488 | 0.123 |
| rh_S_cingul-Marginalis_volume | 1.4696 | 1.8527 | 0.1733 | 0.0878 |
| rh_S_circular_insula_ant_volume | -0.7391 | -1.1911 | 0.8041 | 0.8964 |
| rh_S_circular_insula_inf_volume | -0.6114 | 0.1973 | 0.7834 | 0.4734 |
| rh_S_circular_insula_sup_volume | 0.1314 | -1.0291 | 0.533 | 0.8704 |
| rh_S_collat_transv_ant_volume | 1.0922 | 0.6589 | 0.2564 | 0.3128 |
| rh_S_collat_transv_post_volume | 1.8574 | 2.3969 | 0.1244 | 0.0405 |
| rh_S_front_inf_volume | -0.0689 | 0.5856 | 0.5802 | 0.3395 |
| rh_S_front_middle_volume | 2.7602 | 3.6446 | 0.0508 | 0.0002 |
| rh_S_front_sup_volume | 3.1286 | 3.518 | 0.0341 | 0.0032 |
| rh_S_interm_prim-Jensen_volume | 1.9456 | 1.3546 | 0.1222 | 0.1442 |
| rh_S_intrapariet_and_P_trans_volume | 1.6738 | 1.2688 | 0.1556 | 0.1686 |
| rh_S_oc_middle_and_Lunatus_volume | 2.2506 | 2.3612 | 0.0912 | 0.0459 |
| rh_S_oc_sup_and_transversal_volume | 3.0058 | 3.1826 | 0.0508 | 0.0092 |
| rh_S_occipital_ant_volume | 2.1533 | 2.4697 | 0.0912 | 0.0344 |
| rh_S_oc-temp_lat_volume | 1.7862 | 1.1218 | 0.1222 | 0.1908 |
| rh_S_oc-temp_med_and_Lingual_volume | 1.9511 | 2.2993 | 0.092 | 0.0436 |
| rh_S_orbital_lateral_volume | 1.7255 | 1.4983 | 0.134 | 0.1262 |
| rh_S_orbital_med-olfact_volume | -1.3958 | -0.1678 | 0.9126 | 0.6082 |
| rh_S_orbital-H_Shaped_volume | 0.9793 | 1.447 | 0.2887 | 0.1332 |
| rh_S_parieto_occipital_volume | 2.3598 | 3.5632 | 0.0795 | 0.002 |
| rh_S_pericallosal_volume | 1.1116 | 0.5931 | 0.2512 | 0.3395 |
| rh_S_postcentral_volume | 1.3695 | 2.6371 | 0.206 | 0.0296 |
| rh_S_precentral-inf-part_volume | 0.7738 | 1.52 | 0.32 | 0.1238 |
| rh_S_precentral-sup-part_volume | 2.1405 | 1.6588 | 0.0912 | 0.1099 |
| rh_S_suborbital_volume | 1.0005 | 0.4675 | 0.27 | 0.3674 |
| rh_S_subparietal_volume | -0.9007 | -0.8547 | 0.8312 | 0.8307 |
| rh_S_temporal_inf_volume | 0.6198 | 0.1828 | 0.3683 | 0.4791 |
| rh_S_temporal_sup_volume | 3.4652 | 4.2377 | 0.0083 | 0 |
| rh_S_temporal_transverse_volume | 2.5166 | 1.5213 | 0.0676 | 0.1238 |

**Supplementary Material 6d.**

| CAT_AAL3_cortical* | t_Before | t_After | q_Before | q_After |
| --- | --- | --- | --- | --- |
| lh_Precentral | 3.2443 | 3.5722 | 0.0058 | 0.0015 |
| rh_Precentral | 2.4778 | 3.2309 | 0.0232 | 0.0042 |
| lh_Frontal_Sup_2 | 1.2359 | 3.2895 | 0.144 | 0.001 |
| rh_Frontal_Sup_2 | 1.4894 | 3.1517 | 0.1044 | 0.0035 |
| lh_Frontal_Mid_2 | 2.6632 | 3.8483 | 0.0151 | 0.0002 |
| rh_Frontal_Mid_2 | 2.4664 | 3.4424 | 0.0208 | 0.0002 |
| lh_Frontal_Inf_Oper | 3.5976 | 3.8299 | 0.0012 | 0 |
| rh_Frontal_Inf_Oper | 2.5344 | 2.8704 | 0.0208 | 0.0077 |
| lh_Frontal_Inf_Tri | 2.7031 | 3.5009 | 0.0112 | 0.0002 |
| rh_Frontal_Inf_Tri | 2.2317 | 2.4259 | 0.0323 | 0.0193 |
| lh_Frontal_Inf_Orb_2 | 1.1445 | 0.9744 | 0.1638 | 0.2049 |
| rh_Frontal_Inf_Orb_2 | 1.3386 | 2.052 | 0.1273 | 0.0367 |
| lh_Rolandic_Oper | 2.8476 | 2.9418 | 0.0098 | 0.0047 |
| rh_Rolandic_Oper | 2.3957 | 2.1543 | 0.0218 | 0.0342 |
| lh_Supp_Motor_Area | 1.0565 | 2.2875 | 0.1759 | 0.0305 |
| rh_Supp_Motor_Area | 1.3577 | 3.4465 | 0.1221 | 0.0012 |
| lh_Olfactory | 2.1322 | 1.7958 | 0.0415 | 0.0626 |
| rh_Olfactory | 1.878 | 1.6421 | 0.0677 | 0.0827 |
| lh_Frontal_Sup_Medial | 1.7153 | 3.8616 | 0.0753 | 0 |
| rh_Frontal_Sup_Medial | 1.2819 | 3.204 | 0.1357 | 0.0002 |
| lh_PFC_Vent_Med | 1.1288 | 2.6321 | 0.1642 | 0.0156 |
| rh_PFC_Vent_Med | 0.8405 | 2.2009 | 0.2318 | 0.0342 |
| lh_Rectus | 1.793 | 2.6701 | 0.0753 | 0.0124 |
| rh_Rectus | 2.7501 | 2.3473 | 0.0164 | 0.0266 |
| lh_OFCmed | 2.7558 | 2.2292 | 0.0175 | 0.0362 |
| rh_OFCmed | 1.8111 | 2.861 | 0.0813 | 0.0071 |
| lh_OFCant | 1.8422 | 2.4824 | 0.0746 | 0.0156 |
| rh_OFCant | 1.4609 | 2.3795 | 0.1201 | 0.0199 |
| lh_OFCpost | 2.5027 | 0.8454 | 0.0213 | 0.2395 |
| rh_OFCpost | 2.2865 | 2.5726 | 0.0373 | 0.0157 |
| lh_OFClat | 1.984 | 1.9585 | 0.0513 | 0.047 |
| rh_OFClat | 1.1212 | 2.4866 | 0.1819 | 0.0157 |
| lh_Insula | 1.3075 | 0.894 | 0.1285 | 0.2308 |
| rh_Insula | 1.6032 | 1.1354 | 0.0872 | 0.1625 |
| lh_Cingulate_Mid | 3.5451 | 3.8821 | 0.0019 | 0.0003 |
| rh_Cingulate_Mid | 4.2566 | 4.7727 | 0.0006 | 0 |
| lh_Cingulate_Post | 2.3975 | 2.6852 | 0.0211 | 0.0124 |
| rh_Cingulate_Post | 0.4933 | 1.7533 | 0.3589 | 0.0766 |
| lh_Hippocampus | 1.7405 | 1.9233 | 0.0715 | 0.0498 |
| rh_Hippocampus | 1.4455 | 0.7169 | 0.1112 | 0.2781 |
| lh_ParaHippocampal | 3.1081 | 3.0713 | 0.0066 | 0.0013 |
| rh_ParaHippocampal | 2.0623 | 0.9477 | 0.0488 | 0.2118 |
| lh_Amygdala | 1.5405 | 1.8231 | 0.0992 | 0.0615 |
| rh_Amygdala | 1.2653 | 1.5052 | 0.144 | 0.1038 |
| lh_Calcarine | 4.2523 | 3.8244 | 0.0001 | 0 |
| rh_Calcarine | 4.9479 | 4.679 | 0 | 0 |
| lh_Cuneus | 3.1563 | 2.4632 | 0.003 | 0.0194 |
| rh_Cuneus | 3.9718 | 3.9063 | 0.0001 | 0 |
| lh_Lingual | 3.5047 | 3.3693 | 0.0013 | 0.001 |
| rh_Lingual | 3.5397 | 3.3454 | 0.0012 | 0.0002 |
| lh_Occipital_Sup | 2.7431 | 2.781 | 0.0118 | 0.0119 |
| rh_Occipital_Sup | 2.9917 | 3.4377 | 0.0072 | 0.001 |
| lh_Occipital_Mid | 3.6462 | 3.5719 | 0.0012 | 0.0002 |
| rh_Occipital_Mid | 3.2081 | 5.0369 | 0.0054 | 0 |
| lh_Occipital_Inf | 3.2712 | 2.6031 | 0.0031 | 0.0124 |
| rh_Occipital_Inf | 2.3894 | 3.2033 | 0.0268 | 0.0024 |
| lh_Fusiform | 3.189 | 3.023 | 0.002 | 0.0011 |
| rh_Fusiform | 2.6462 | 2.5685 | 0.0098 | 0.0068 |
| lh_Postcentral | 3.7101 | 4.0927 | 0.002 | 0.0002 |
| rh_Postcentral | 3.7036 | 5.0716 | 0.0013 | 0.0002 |
| lh_Parietal_Sup | 3.8031 | 3.026 | 0.0013 | 0.0035 |
| rh_Parietal_Sup | 3.0499 | 3.7018 | 0.0063 | 0.0001 |
| lh_Parietal_Inf | 6.121 | 5.3536 | 0 | 0.0003 |
| rh_Parietal_Inf | 1.924 | 3.0336 | 0.0589 | 0.0061 |
| lh_SupraMarginal | 3.7693 | 2.9791 | 0.0001 | 0.0051 |
| rh_SupraMarginal | 2.8855 | 4.0426 | 0.0105 | 0 |
| lh_Angular | 4.864 | 4.3661 | 0 | 0.0002 |
| rh_Angular | 2.8396 | 4.0039 | 0.0105 | 0.0001 |
| lh_Precuneus | 2.9851 | 2.6455 | 0.0098 | 0.0156 |
| rh_Precuneus | 4.1208 | 4.1187 | 0.0005 | 0 |
| lh_Paracentral_Lobule | 0.6307 | 1.3338 | 0.3129 | 0.1331 |
| rh_Paracentral_Lobule | 0.8099 | 1.8996 | 0.2419 | 0.0565 |
| lh_Heschl | 3.8095 | 3.3248 | 0.0008 | 0.0013 |
| rh_Heschl | 3.1822 | 3.3443 | 0.0032 | 0.0016 |
| lh_Temporal_Sup | 3.1667 | 3.1525 | 0.0024 | 0.0002 |
| rh_Temporal_Sup | 3.6863 | 4.3904 | 0 | 0 |
| lh_Temporal_Pole_Sup | 3.4587 | 1.4465 | 0.0026 | 0.1038 |
| rh_Temporal_Pole_Sup | 2.7737 | 1.3341 | 0.0105 | 0.1269 |
| lh_Temporal_Mid | 3.2198 | 3.6642 | 0.002 | 0 |
| rh_Temporal_Mid | 3.3228 | 3.6151 | 0.0013 | 0 |
| lh_Temporal_Pole_Mid | 2.1495 | 2.0854 | 0.0415 | 0.0412 |
| rh_Temporal_Pole_Mid | 2.4956 | 1.8359 | 0.0218 | 0.0565 |
| lh_Temporal_Inf | 2.82 | 3.0705 | 0.0086 | 0.0017 |
| rh_Temporal_Inf | 2.438 | 1.838 | 0.0208 | 0.0565 |
| lh_ACC_sub | 1.8225 | 2.144 | 0.0653 | 0.0384 |
| rh_ACC_sub | 0.2099 | 0.2629 | 0.4498 | 0.4216 |
| lh_ACC_pre | 1.4174 | 2.5615 | 0.1106 | 0.0156 |
| rh_ACC_pre | 1.6273 | 3.1999 | 0.0834 | 0.0016 |
| lh_ACC_sup | 1.5902 | 2.5022 | 0.0885 | 0.0156 |
| rh_ACC_sup | 1.4258 | 2.4817 | 0.1106 | 0.0194 |

**Supplementary Material 6e.**
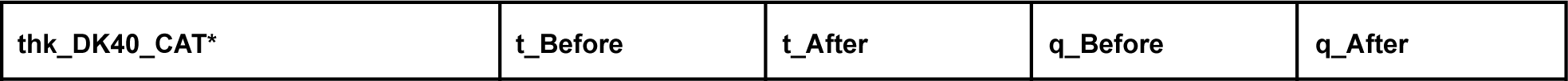
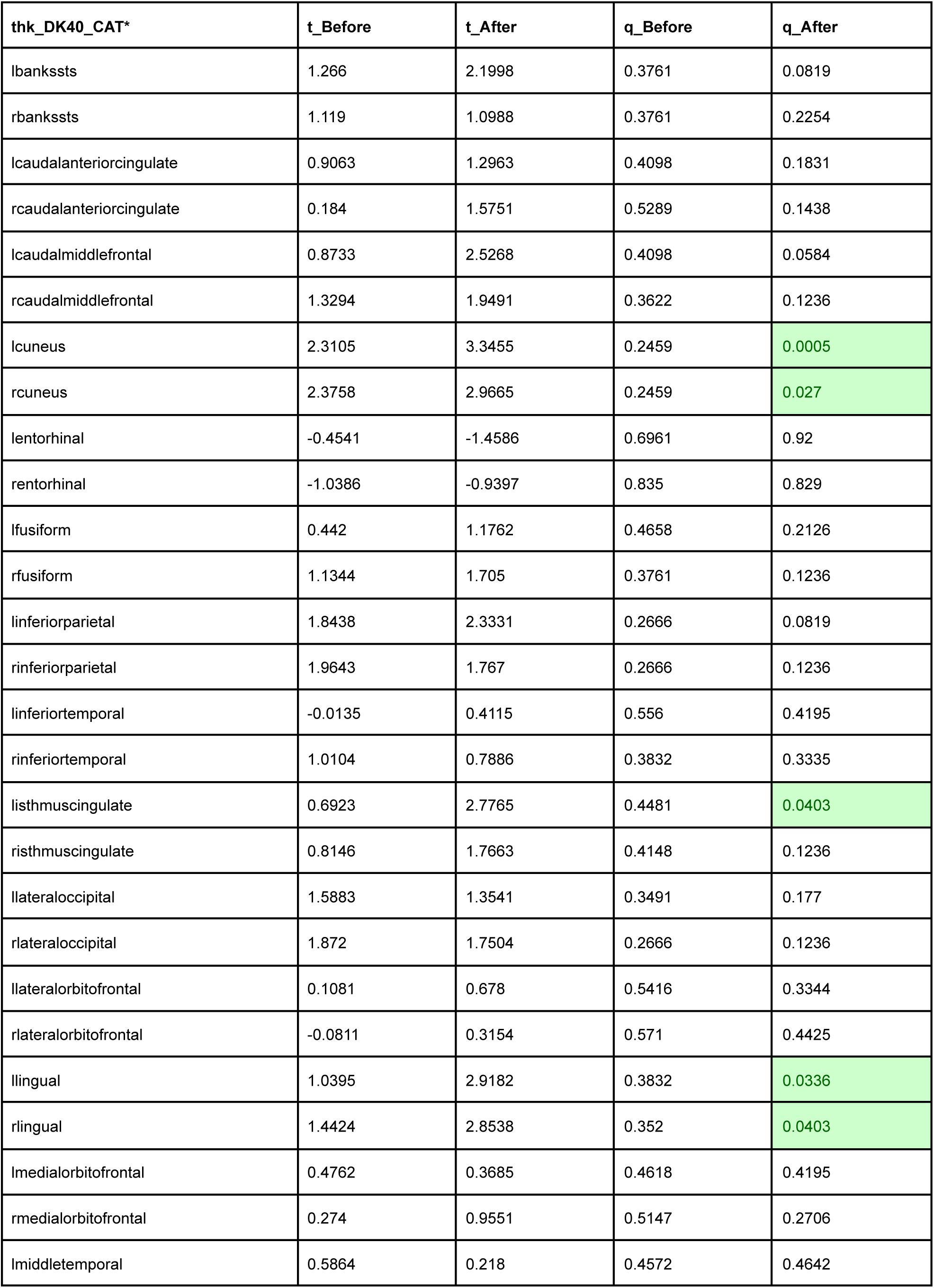
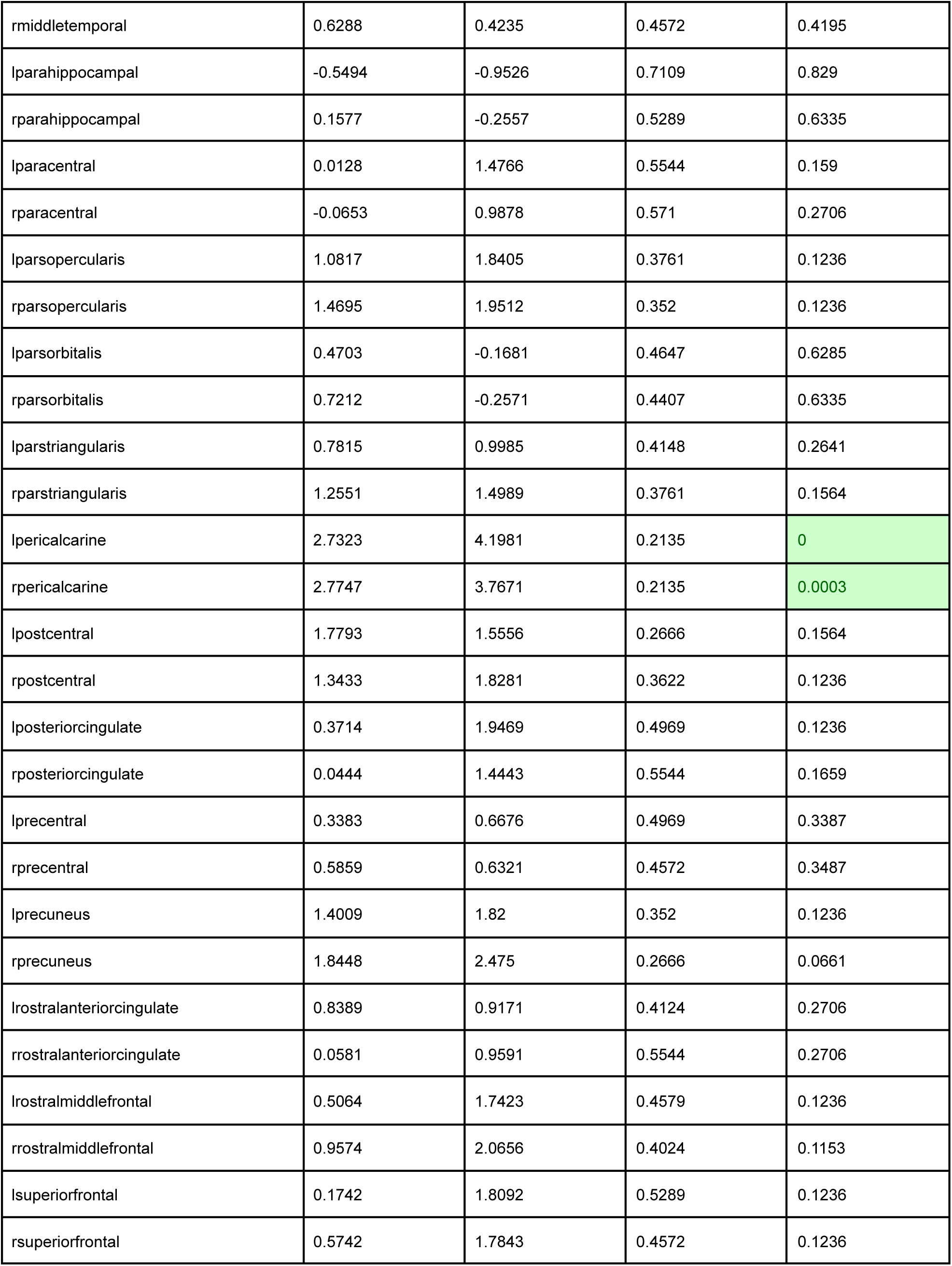
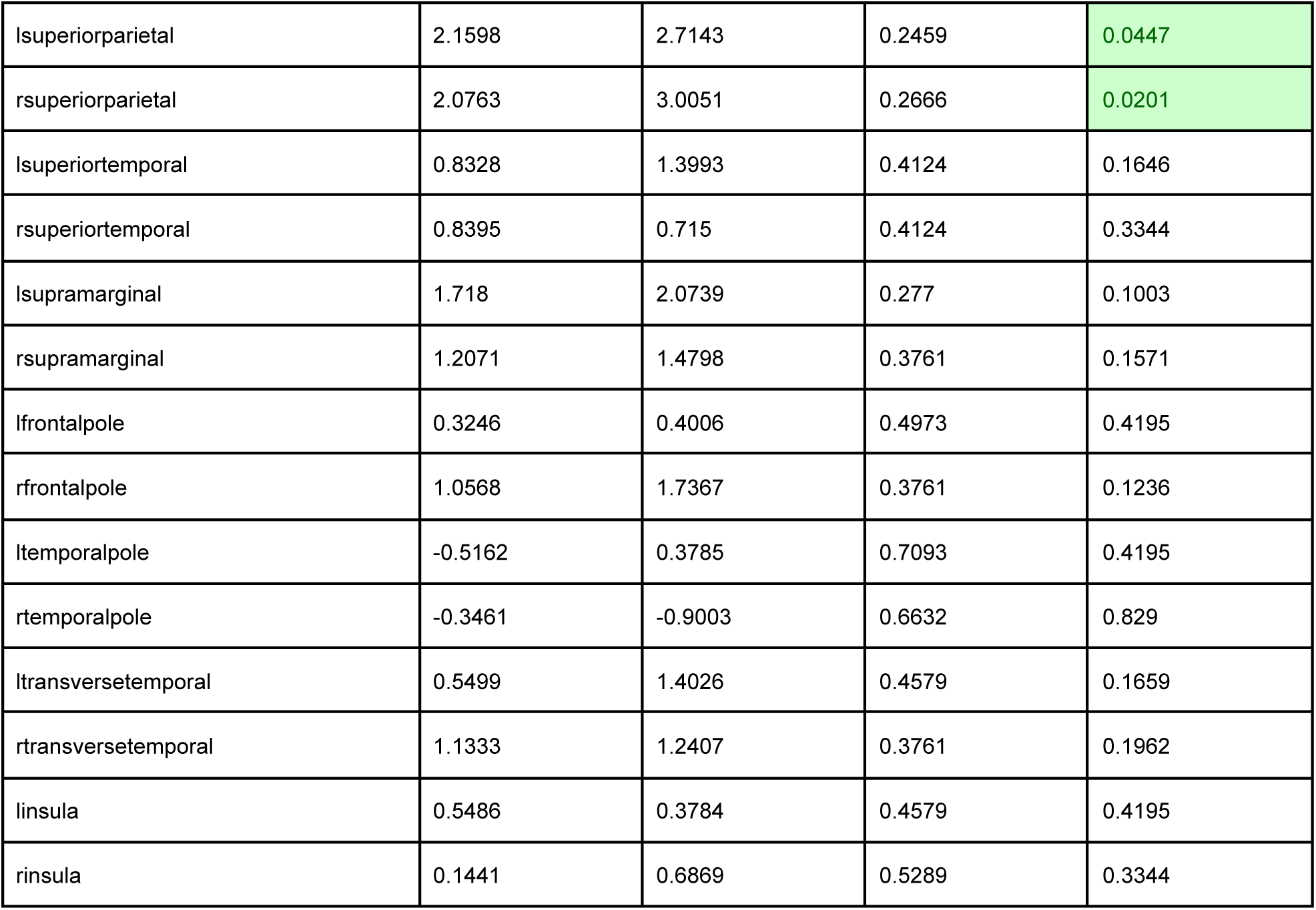

**Supplementary Material 6f.**

| thk_DK40_FastSurfer* | t_Before | t_After | q_Before | q_After |
| --- | --- | --- | --- | --- |
| lh_bankssts_thickness | 0.9928 | 2.1537 | 0.6994 | 0.0695 |
| lh_caudalanteriorcingulate_thickness | -0.6604 | 0.0196 | 0.9079 | 0.5407 |
| lh_caudalmiddlefrontal_thickness | -0.1206 | 2.5908 | 0.9079 | 0.0582 |
| lh_cuneus_thickness | -1.4156 | 0.2236 | 0.9893 | 0.4818 |
| lh_entorhinal_thickness | -0.3994 | 0.1544 | 0.9079 | 0.5063 |
| lh_fusiform_thickness | 0.4503 | 2.4612 | 0.8068 | 0.0582 |
| lh_inferiorparietal_thickness | 1.4271 | 2.8676 | 0.6994 | 0.0582 |
| lh_inferiortemporal_thickness | -0.5829 | 2.296 | 0.9079 | 0.0606 |
| lh_isthmuscingulate_thickness | 1.216 | 1.4598 | 0.6994 | 0.1453 |
| lh_lateraloccipital_thickness | 1.1208 | 1.7996 | 0.6994 | 0.1063 |
| lh_lateralorbitofrontal_thickness | -0.2396 | 1.6753 | 0.9079 | 0.1197 |
| lh_lingual_thickness | 0.464 | 1.8744 | 0.8068 | 0.1063 |
| lh_medialorbitofrontal_thickness | -2.0191 | 2.2921 | 0.9893 | 0.0606 |
| lh_middletemporal_thickness | 0.6086 | 2.9648 | 0.7913 | 0.0582 |
| lh_parahippocampal_thickness | 1.295 | 1.1349 | 0.6994 | 0.2153 |
| lh_paracentral_thickness | -0.7028 | -0.0156 | 0.9079 | 0.5407 |
| lh_parsopercularis_thickness | 0.531 | 2.3521 | 0.7913 | 0.0606 |
| lh_parsorbitalis_thickness | -0.1377 | 1.7261 | 0.9079 | 0.1223 |
| lh_parstriangularis_thickness | -0.2188 | 2.0559 | 0.9079 | 0.0815 |
| lh_pericalcarine_thickness | 0.0255 | 0.9018 | 0.9079 | 0.2646 |
| lh_postcentral_thickness | -0.6792 | 1.0145 | 0.9079 | 0.2462 |
| lh_posteriorcingulate_thickness | 1.0296 | 0.7885 | 0.6994 | 0.3006 |
| lh_precentral_thickness | -0.5937 | 0.3338 | 0.9079 | 0.4507 |
| lh_precuneus_thickness | 1.3178 | 1.8029 | 0.6994 | 0.1131 |
| lh_rostralanteriorcingulate_thickness | 0.8183 | 1.2194 | 0.6994 | 0.1912 |
| lh_rostralmiddlefrontal_thickness | -2.0856 | 1.6352 | 0.9893 | 0.1232 |
| lh_superiorfrontal_thickness | -0.2293 | 1.7632 | 0.9079 | 0.1142 |
| lh_superiorparietal_thickness | 1.2899 | 2.9824 | 0.6994 | 0.0582 |
| lh_superiortemporal_thickness | 0.3211 | 2.2065 | 0.8184 | 0.0597 |
| lh_supramarginal_thickness | 1.8286 | 2.3075 | 0.6994 | 0.0606 |
| lh_frontalpole_thickness | 0.5855 | 2.7286 | 0.7913 | 0.0582 |
| lh_temporalpole_thickness | -0.7682 | 1.5651 | 0.9079 | 0.1232 |
| lh_transversetemporal_thickness | -1.083 | 0.6724 | 0.9671 | 0.3534 |
| lh_insula_thickness | -1.5881 | -0.4735 | 0.9893 | 0.6682 |
| lh_MeanThickness_thickness | 0.1211 | 2.248 | 0.9079 | 0.0609 |
| rh_bankssts_thickness | 1.0731 | 1.5492 | 0.6994 | 0.127 |
| rh_caudalanteriorcingulate_thickness | -0.2956 | 0.9182 | 0.9079 | 0.2654 |
| rh_caudalmiddlefrontal_thickness | -0.313 | 1.3991 | 0.9079 | 0.1503 |
| rh_cuneus_thickness | -4.0738 | 0.1331 | 1 | 0.5063 |
| rh_entorhinal_thickness | -1.9391 | 0.6279 | 0.9893 | 0.3534 |
| rh_fusiform_thickness | 1.1915 | 2.0143 | 0.6994 | 0.0815 |
| rh_inferiorparietal_thickness | 1.7908 | 2.5601 | 0.6994 | 0.0582 |
| rh_inferiortemporal_thickness | -0.5182 | 0.4236 | 0.9079 | 0.4285 |
| rh_isthmuscingulate_thickness | 1.1555 | 0.7393 | 0.6994 | 0.3134 |
| rh_lateraloccipital_thickness | 1.1378 | 1.3586 | 0.6994 | 0.1503 |
| rh_lateralorbitofrontal_thickness | -0.4188 | 1.5732 | 0.9079 | 0.1232 |
| rh_lingual_thickness | -0.5267 | 1.6301 | 0.9079 | 0.1232 |
| rh_medialorbitofrontal_thickness | -1.5476 | 0.0896 | 0.9893 | 0.5082 |
| rh_middletemporal_thickness | 0.6525 | 2.0103 | 0.7913 | 0.0815 |
| rh_parahippocampal_thickness | 0.8957 | 0.4656 | 0.6994 | 0.4134 |
| rh_paracentral_thickness | -0.6145 | 1.0785 | 0.9079 | 0.2267 |
| rh_parsopercularis_thickness | 0.8766 | 2.6442 | 0.6994 | 0.0582 |
| rh_parsorbitalis_thickness | 0.5427 | 2.3317 | 0.7913 | 0.0606 |
| rh_parstriangularis_thickness | 0.8228 | 2.1074 | 0.6994 | 0.0609 |
| rh_pericalcarine_thickness | -1.6171 | 2.8623 | 0.9893 | 0.0582 |
| rh_postcentral_thickness | -1.3433 | 1.0101 | 0.9893 | 0.2359 |
| rh_posteriorcingulate_thickness | 1.2761 | 0.3697 | 0.6994 | 0.4333 |
| rh_precentral_thickness | -0.7776 | 0.0067 | 0.9079 | 0.5355 |
| rh_precuneus_thickness | 0.9493 | 2.3441 | 0.6994 | 0.0606 |
| rh_rostralanteriorcingulate_thickness | -0.7204 | -0.0801 | 0.9079 | 0.5564 |
| rh_rostralmiddlefrontal_thickness | -0.1914 | 1.3901 | 0.9079 | 0.1503 |
| rh_superiorfrontal_thickness | 0.0373 | 1.7492 | 0.9079 | 0.1146 |
| rh_superiorparietal_thickness | 1.7078 | 2.9987 | 0.6994 | 0.0582 |
| rh_superiortemporal_thickness | 0.3402 | 2.376 | 0.8068 | 0.0582 |
| rh_supramarginal_thickness | 0.9539 | 1.5786 | 0.6994 | 0.1232 |
| rh_frontalpole_thickness | 0.4332 | 2.5741 | 0.8068 | 0.0582 |
| rh_temporalpole_thickness | 0.3701 | 0.2273 | 0.8068 | 0.4818 |
| rh_transversetemporal_thickness | -0.1501 | -0.4376 | 0.9079 | 0.6682 |
| rh_insula_thickness | -0.1069 | -0.1169 | 0.9079 | 0.5565 |
| rh_MeanThickness_thickness | 0.0585 | 1.9677 | 0.9079 | 0.0943 |

**Supplementary Material 6g.**

| vol_DK40_FastSurfer* | t_Before | t_After | q_Before | q_After |
| --- | --- | --- | --- | --- |
| lh_bankssts_volume | 1.5236 | 2.8004 | 0.1448 | 0.0143 |
| lh_caudalanteriorcingulate_volume | 1.0668 | 1.1185 | 0.235 | 0.169 |
| lh_caudalmiddlefrontal_volume | 1.7696 | 2.5708 | 0.1185 | 0.0195 |
| lh_cuneus_volume | 1.1469 | 1.5562 | 0.2239 | 0.1054 |
| lh_entorhinal_volume | 0.4039 | 1.8156 | 0.4098 | 0.057 |
| lh_fusiform_volume | 1.0258 | 1.843 | 0.2739 | 0.057 |
| lh_inferiorparietal_volume | 2.5851 | 4.3203 | 0.0389 | 0.0011 |
| lh_inferiortemporal_volume | 0.8549 | 2.2732 | 0.2998 | 0.0286 |
| lh_isthmuscingulate_volume | 1.2935 | 1.3562 | 0.1958 | 0.1297 |
| lh_lateraloccipital_volume | 1.8641 | 2.3078 | 0.1045 | 0.0261 |
| lh_lateralorbitofrontal_volume | 0.7714 | 1.8443 | 0.3187 | 0.057 |
| lh_lingual_volume | 2.7146 | 3.3983 | 0.0334 | 0.0001 |
| lh_medialorbitofrontal_volume | -1.0075 | 0.8488 | 0.8308 | 0.2317 |
| lh_middletemporal_volume | 1.6018 | 2.9005 | 0.1288 | 0.0065 |
| lh_parahippocampal_volume | 0.5859 | 1.1237 | 0.3658 | 0.169 |
| lh_paracentral_volume | 1.8757 | 2.1809 | 0.1061 | 0.041 |
| lh_parsopercularis_volume | 0.8112 | 2.05 | 0.2998 | 0.0521 |
| lh_parsorbitalis_volume | 0.7772 | 1.7839 | 0.3285 | 0.057 |
| lh_parstriangularis_volume | 1.2799 | 2.3285 | 0.1958 | 0.0302 |
| lh_pericalcarine_volume | 2.5786 | 2.5031 | 0.0389 | 0.0226 |
| lh_postcentral_volume | 1.6425 | 2.9593 | 0.1298 | 0.0086 |
| lh_posteriorcingulate_volume | 2.6712 | 2.2112 | 0.0389 | 0.034 |
| lh_precentral_volume | 1.6423 | 2.3833 | 0.1288 | 0.0302 |
| lh_precuneus_volume | 2.3626 | 2.8008 | 0.0389 | 0.0092 |
| lh_rostralanteriorcingulate_volume | 1.0845 | 0.9494 | 0.2318 | 0.2026 |
| lh_rostralmiddlefrontal_volume | 1.9997 | 3.5034 | 0.0682 | 0.0001 |
| lh_superiorfrontal_volume | 1.7743 | 2.7116 | 0.1097 | 0.0135 |
| lh_superiorparietal_volume | 2.6841 | 3.5941 | 0.0364 | 0.0009 |
| lh_superiortemporal_volume | 1.2072 | 2.2415 | 0.1999 | 0.0228 |
| lh_supramarginal_volume | 2.7967 | 2.9188 | 0.0334 | 0.0065 |
| lh_frontalpole_volume | -0.0113 | 1.031 | 0.5565 | 0.1881 |
| lh_temporalpole_volume | 0.3426 | 1.5385 | 0.4098 | 0.094 |
| lh_transversetemporal_volume | 0.609 | 1.3372 | 0.3405 | 0.1297 |
| lh_insula_volume | -0.6982 | -1.3906 | 0.7733 | 0.9299 |
| rh_bankssts_volume | 2.9724 | 3.3538 | 0.0334 | 0.0016 |
| rh_caudalanteriorcingulate_volume | 0.6021 | 0.4496 | 0.3405 | 0.3482 |
| rh_caudalmiddlefrontal_volume | 1.2809 | 2.1165 | 0.1958 | 0.0421 |
| rh_cuneus_volume | 2.1472 | 4.0389 | 0.0749 | 0 |
| rh_entorhinal_volume | -0.8389 | 0.8018 | 0.8093 | 0.2397 |
| rh_fusiform_volume | 1.6564 | 1.9865 | 0.1144 | 0.0402 |
| rh_inferiorparietal_volume | 2.6718 | 3.3106 | 0.0389 | 0.0042 |
| rh_inferiortemporal_volume | 0.8184 | 1.0104 | 0.2998 | 0.1881 |
| rh_isthmuscingulate_volume | 0.59 | -0.0427 | 0.3588 | 0.5397 |
| rh_lateraloccipital_volume | 3.0822 | 3.3862 | 0.0334 | 0.0021 |
| rh_lateralorbitofrontal_volume | -0.2376 | 0.6009 | 0.6267 | 0.3143 |
| rh_lingual_volume | 2.0258 | 3.0053 | 0.072 | 0.0036 |
| rh_medialorbitofrontal_volume | 0.7266 | 1.3853 | 0.3187 | 0.1225 |
| rh_middletemporal_volume | 2.6801 | 3.603 | 0.0334 | 0 |
| rh_parahippocampal_volume | 0.5687 | 0.0739 | 0.3649 | 0.4953 |
| rh_paracentral_volume | 2.2451 | 2.9896 | 0.0682 | 0.0092 |
| rh_parsopercularis_volume | 0.5508 | 1.2762 | 0.3588 | 0.1478 |
| rh_parsorbitalis_volume | 0.897 | 1.7765 | 0.2998 | 0.064 |
| rh_parstriangularis_volume | 0.6912 | 1.1308 | 0.3405 | 0.169 |
| rh_pericalcarine_volume | 1.7294 | 3.7643 | 0.1189 | 0 |
| rh_postcentral_volume | 1.3108 | 2.9312 | 0.1967 | 0.0127 |
| rh_posteriorcingulate_volume | 2.3405 | 1.9373 | 0.0537 | 0.057 |
| rh_precentral_volume | 1.3356 | 2.3859 | 0.1958 | 0.0302 |
| rh_precuneus_volume | 1.9916 | 2.9871 | 0.0912 | 0.0076 |
| rh_rostralanteriorcingulate_volume | 0.1424 | -0.0535 | 0.4927 | 0.5397 |
| rh_rostralmiddlefrontal_volume | 2.2545 | 3.0855 | 0.0389 | 0.0027 |
| rh_superiorfrontal_volume | 2.5794 | 3.9108 | 0.0334 | 0 |
| rh_superiorparietal_volume | 2.5805 | 3.6488 | 0.0389 | 0.0011 |
| rh_superiortemporal_volume | 2.2543 | 3.4028 | 0.0389 | 0 |
| rh_supramarginal_volume | 2.2517 | 3.1772 | 0.0537 | 0.0027 |
| rh_frontalpole_volume | 1.4832 | 2.8978 | 0.1548 | 0.0103 |
| rh_temporalpole_volume | 0.8999 | 1.457 | 0.2883 | 0.1084 |
| rh_transversetemporal_volume | 1.986 | 1.2156 | 0.0762 | 0.1478 |
| rh_insula_volume | -0.2727 | -2.064 | 0.6392 | 0.9746 |

**Supplementary Material 6h.**

| SynthSeg* | t_Before | t_After | q_Before | q_After |
| --- | --- | --- | --- | --- |
| left cerebral white matter | 1.6167 | 1.7859 | 0.1088 | 0.0756 |
| left cerebral cortex | 4.5603 | 4.6303 | 0.0004 | 0 |
| left lateral ventricle | -2.82 | -2.8372 | 1 | 1 |
| left inferior lateral ventricle | -1.9072 | -1.9284 | 1 | 1 |
| left cerebellum white matter | 0.2298 | 0.1084 | 0.4786 | 0.5055 |
| left cerebellum cortex | 1.6144 | 2.0103 | 0.1088 | 0.0564 |
| left thalamus | 0.6907 | 0.7461 | 0.3316 | 0.2997 |
| left caudate | 2.4751 | 2.3802 | 0.0382 | 0.0366 |
| left putamen | 1.6581 | 1.8711 | 0.1088 | 0.0719 |
| left pallidum | 0.8694 | 1.3045 | 0.2779 | 0.1578 |
| 3rd ventricle | -2.7606 | -2.7622 | 1 | 1 |
| 4th ventricle | -3.9309 | -4.1813 | 1 | 1 |
| brain-stem | 2.6018 | 2.7102 | 0.0329 | 0.0212 |
| left hippocampus | 1.5035 | 1.4937 | 0.1332 | 0.1245 |
| left amygdala | 1.3075 | 0.7959 | 0.1654 | 0.2768 |
| csf | -2.9362 | -3.327 | 1 | 1 |
| left accumbens area | 3.2741 | 3.1859 | 0.0081 | 0.0076 |
| left ventral DC | 1.8798 | 2.0583 | 0.0894 | 0.0564 |
| right cerebral white matter | 1.8095 | 1.9468 | 0.0894 | 0.0593 |
| right cerebral cortex | 4.5196 | 5.0468 | 0 | 0 |
| right lateral ventricle | -2.5586 | -2.5589 | 1 | 1 |
| right inferior lateral ventricle | -1.8787 | -1.827 | 1 | 1 |
| right cerebellum white matter | -0.6779 | -1.1115 | 0.8007 | 0.9048 |
| right cerebellum cortex | 1.3219 | 1.7792 | 0.1614 | 0.0772 |
| right thalamus | 1.053 | 0.7524 | 0.2221 | 0.3007 |
| right caudate | 2.5321 | 2.3365 | 0.0329 | 0.0366 |
| right putamen | 2.3271 | 2.5335 | 0.0436 | 0.026 |
| right pallidum | 1.5362 | 1.468 | 0.1258 | 0.1283 |
| right hippocampus | 0.9464 | 0.9336 | 0.253 | 0.2453 |
| right amygdala | 1.4691 | 1.4156 | 0.1465 | 0.1439 |
| right accumbens area | 4.464 | 4.0394 | 0 | 0 |
| right ventral DC | 2.0492 | 2.3916 | 0.0652 | 0.0348 |
| ctx-lh-bankssts | 0.7205 | 1.1523 | 0.3053 | 0.1806 |
| ctx-lh-caudalanteriorcingulate | 0.5181 | 0.8387 | 0.3692 | 0.2538 |
| ctx-lh-caudalmiddlefrontal | 1.1537 | 1.1114 | 0.1993 | 0.2057 |
| ctx-lh-cuneus | 1.1797 | 1.5898 | 0.1878 | 0.101 |
| ctx-lh-entorhinal | 2.2698 | 2.9851 | 0.0436 | 0.0088 |
| ctx-lh-fusiform | 2.2508 | 2.2455 | 0.0519 | 0.0443 |
| ctx-lh-inferiorparietal | 2.6242 | 2.5355 | 0.0344 | 0.0348 |
| ctx-lh-inferiortemporal | 1.8357 | 1.9262 | 0.0883 | 0.0647 |
| ctx-lh-isthmuscingulate | 0.9325 | 1.0333 | 0.2367 | 0.2057 |
| ctx-lh-lateraloccipital | 0.9353 | 1.0516 | 0.2561 | 0.2107 |
| ctx-lh-lateralorbitofrontal | 0.4391 | 0.5075 | 0.3903 | 0.3659 |
| ctx-lh-lingual | 4.2117 | 4.1593 | 0 | 0 |
| ctx-lh-medialorbitofrontal | 0.5696 | 0.1205 | 0.3422 | 0.4889 |
| ctx-lh-middletemporal | 2.7801 | 2.7677 | 0.0253 | 0.0156 |
| ctx-lh-parahippocampal | 2.1008 | 1.9591 | 0.0509 | 0.0563 |
| ctx-lh-paracentral | 1.3636 | 1.5462 | 0.1522 | 0.1125 |
| ctx-lh-parsopercularis | 1.3223 | 1.187 | 0.1654 | 0.1837 |
| ctx-lh-parsorbitalis | 0.2047 | 0.3074 | 0.4786 | 0.4373 |
| ctx-lh-parstriangularis | 1.0645 | 1.4398 | 0.2039 | 0.1245 |
| ctx-lh-pericalcarine | 1.6248 | 1.7361 | 0.1064 | 0.0756 |
| ctx-lh-postcentral | 3.8901 | 3.9466 | 0.0065 | 0.0031 |
| ctx-lh-posteriorcingulate | 3.8948 | 4.0191 | 0 | 0 |
| ctx-lh-precentral | 1.868 | 2.034 | 0.0837 | 0.0564 |
| ctx-lh-precuneus | 1.1626 | 1.1978 | 0.1977 | 0.1837 |
| ctx-lh-rostralanteriorcingulate | 1.5694 | 1.5633 | 0.1163 | 0.1075 |
| ctx-lh-rostralmiddlefrontal | 2.7948 | 2.8813 | 0.0261 | 0.014 |
| ctx-lh-superiorfrontal | 2.0058 | 2.1242 | 0.0719 | 0.0517 |
| ctx-lh-superiorparietal | 0.1017 | 0.2152 | 0.5296 | 0.4774 |
| ctx-lh-superiortemporal | 1.7909 | 2.2491 | 0.0895 | 0.0398 |
| ctx-lh-supramarginal | 1.7086 | 1.8189 | 0.1018 | 0.0728 |
| ctx-lh-frontalpole | -0.4884 | -0.4795 | 0.7351 | 0.7316 |
| ctx-lh-temporalpole | 1.6941 | 1.1278 | 0.1018 | 0.1837 |
| ctx-lh-transversetemporal | 0.3847 | 0.4793 | 0.4144 | 0.3794 |
| ctx-lh-insula | -1.8203 | -0.9627 | 1 | 0.8846 |
| ctx-rh-bankssts | 1.6664 | 1.7074 | 0.1088 | 0.0904 |
| ctx-rh-caudalanteriorcingulate | 0.6541 | 1.0014 | 0.3189 | 0.2104 |
| ctx-rh-caudalmiddlefrontal | 1.1594 | 1.1091 | 0.1975 | 0.2057 |
| ctx-rh-cuneus | 2.2871 | 2.4963 | 0.0496 | 0.0315 |
| ctx-rh-entorhinal | -0.7415 | -0.6253 | 0.8109 | 0.7832 |
| ctx-rh-fusiform | 2.719 | 3.1125 | 0.0288 | 0.0064 |
| ctx-rh-inferiorparietal | 3.4516 | 3.8689 | 0.0078 | 0.0021 |
| ctx-rh-inferiortemporal | 1.0869 | 1.0583 | 0.2039 | 0.2057 |
| ctx-rh-isthmuscingulate | -0.1407 | 0.1076 | 0.6234 | 0.5055 |
| ctx-rh-lateraloccipital | 2.4576 | 2.6827 | 0.0329 | 0.0156 |
| ctx-rh-lateralorbitofrontal | 0.1787 | 0.6019 | 0.4844 | 0.3306 |
| ctx-rh-lingual | 1.602 | 1.6232 | 0.1088 | 0.0958 |
| ctx-rh-medialorbitofrontal | 0.769 | 0.7197 | 0.2865 | 0.2905 |
| ctx-rh-middletemporal | 3.9457 | 4.4878 | 0.0006 | 0 |
| ctx-rh-parahippocampal | 0.1415 | 0.405 | 0.4903 | 0.3811 |
| ctx-rh-paracentral | 2.0229 | 2.251 | 0.0729 | 0.0443 |
| ctx-rh-parsopercularis | -0.0775 | 0.2788 | 0.5853 | 0.4423 |
| ctx-rh-parsorbitalis | 1.6867 | 1.9495 | 0.1064 | 0.0656 |
| ctx-rh-parstriangularis | 0.453 | 0.4739 | 0.3903 | 0.3811 |
| ctx-rh-pericalcarine | 1.422 | 1.2217 | 0.1422 | 0.1758 |
| ctx-rh-postcentral | 2.7152 | 2.9993 | 0.0269 | 0.0076 |
| ctx-rh-posteriorcingulate | 1.655 | 2.0219 | 0.1064 | 0.0519 |
| ctx-rh-precentral | 3.388 | 3.6099 | 0.0078 | 0.0027 |
| ctx-rh-precuneus | 1.3329 | 1.5909 | 0.1654 | 0.1125 |
| ctx-rh-rostralanteriorcingulate | 0.591 | 1.3406 | 0.3463 | 0.1575 |
| ctx-rh-rostralmiddlefrontal | 2.692 | 3.2274 | 0.0288 | 0.0064 |
| ctx-rh-superiorfrontal | 2.2116 | 2.2518 | 0.0514 | 0.0418 |
| ctx-rh-superiorparietal | 0.4519 | 0.8095 | 0.3885 | 0.2697 |
| ctx-rh-superiortemporal | 1.1853 | 2.072 | 0.1875 | 0.0505 |
| ctx-rh-supramarginal | 2.8967 | 3.4743 | 0.0156 | 0.0021 |
| ctx-rh-frontalpole | 0.4304 | 0.3066 | 0.3903 | 0.4278 |
| ctx-rh-temporalpole | 1.3908 | 1.8204 | 0.1453 | 0.069 |
| ctx-rh-transversetemporal | -0.2262 | 0.1285 | 0.6375 | 0.4914 |
| ctx-rh-insula | -1.7665 | -1.1112 | 1 | 0.9119 |

**Supplementary Material 6i.**

| CAT_AAL3_subcortical* | t_Before | t_After | q_Before | q_After |
| --- | --- | --- | --- | --- |
| Caudate_left | 2.5828 | 2.7093 | 0.0175 | 0.0086 |
| Caudate_right | 2.6491 | 2.5388 | 0.0126 | 0.0131 |
| Putamen_left | 0.5831 | 1.6688 | 0.3535 | 0.0766 |
| Putamen_right | 0.3162 | 1.3458 | 0.4299 | 0.1282 |
| Pallidum_left | -1.082 | -0.479 | 0.8685 | 0.6478 |
| Pallidum_right | -0.4637 | 0.2338 | 0.6636 | 0.4386 |
| Thal_AV_left | 1.0798 | -0.5471 | 0.1819 | 0.7112 |
| Thal_AV_right | 1.7516 | 0.7708 | 0.0772 | 0.2608 |
| Thal_LP_left | -0.5536 | 1.514 | 0.7365 | 0.0975 |
| Thal_LP_right | 1.5005 | 1.5945 | 0.1044 | 0.0897 |
| Thal_VA_left | -0.3056 | -0.9698 | 0.6611 | 0.8354 |
| Thal_VA_right | -1.0666 | -0.074 | 0.8634 | 0.538 |
| Thal_VL_left | 0.252 | 1.0642 | 0.4498 | 0.1786 |
| Thal_VL_right | -0.206 | 0.6432 | 0.6162 | 0.2939 |
| Thal_VPL_left | -0.4281 | 0.3807 | 0.6998 | 0.389 |
| Thal_VPL_right | -1.1768 | -0.2658 | 0.8859 | 0.6271 |
| Thal_IL_left | -0.7111 | 0.7498 | 0.7851 | 0.2718 |
| Thal_IL_right | 0.0374 | 0.2901 | 0.5304 | 0.4153 |
| Thal_MDM_left | 2.492 | 1.5222 | 0.0218 | 0.0962 |
| Thal_MDM_right | 2.6827 | 1.4378 | 0.0168 | 0.1134 |
| Thal_MDL_left | 2.2047 | 1.0242 | 0.0372 | 0.1937 |
| Thal_MDL_right | 1.9547 | 1.2254 | 0.0614 | 0.1553 |
| Thal_LGN_left | -0.416 | 0.7479 | 0.6913 | 0.2608 |
| Thal_LGN_right | -0.0363 | 0.9107 | 0.5647 | 0.2229 |
| Thal_MGN_left | -3.2297 | -0.7453 | 0.9998 | 0.7619 |
| Thal_MGN_right | -2.092 | 0.1059 | 0.9834 | 0.4721 |
| Thal_PUA_left | 0.3949 | 1.7052 | 0.3961 | 0.0721 |
| Thal_PUA_right | 1.6112 | 1.8543 | 0.0885 | 0.0565 |
| Thal_PUM_left | 1.3142 | 1.475 | 0.1357 | 0.1038 |
| Thal_PUM_right | 2.2372 | 1.8445 | 0.0296 | 0.0565 |
| Thal_PUL_left | 1.2616 | 1.0729 | 0.1441 | 0.1797 |
| Thal_PUL_right | 1.9691 | 1.2723 | 0.0579 | 0.1468 |
| Thal_PUI_left | -0.246 | 1.1199 | 0.6421 | 0.1786 |
| Thal_PUI_right | 0.8503 | -0.0901 | 0.2318 | 0.5552 |
| N_Acc_left | 2.2976 | 2.0658 | 0.0338 | 0.0417 |
| N_Acc_right | 2.9041 | 1.9248 | 0.0122 | 0.0584 |
| Raphe_Dorsal | 0.4511 | 0.7365 | 0.3758 | 0.2709 |
| Raphe_Median | 1.2177 | 0.6865 | 0.1575 | 0.2832 |

**Supplementary Material 6j.**

| CAT_AAL3_cerebellum* | t_Before | t_After | q_Before | q_After |
| --- | --- | --- | --- | --- |
| Cerebellum_CRU1_left | 4.1739 | 4.373 | 0.0005 | 0 |
| Cerebellum_CRU1_right | 4.1518 | 5.1526 | 0.0012 | 0 |
| Cerebellum_CRU2_left | 3.5539 | 4.0316 | 0.0019 | 0.0006 |
| Cerebellum_CRU2_right | 3.911 | 5.4901 | 0.0013 | 0 |
| Cerebellum_3_left | 2.8466 | 3.2481 | 0.0105 | 0.0037 |
| Cerebellum_3_right | 2.9347 | 2.9448 | 0.0098 | 0.0084 |
| Cerebellum_4_5_left | 3.7319 | 4.2693 | 0.0008 | 0.0002 |
| Cerebellum_4_5_right | 3.5824 | 4.3068 | 0.0013 | 0.0007 |
| Cerebellum_6_left | 4.3585 | 5.0619 | 0.0001 | 0 |
| Cerebellum_6_right | 4.4978 | 5.2407 | 0.0001 | 0 |
| Cerebellum_7b_left | 3.9338 | 3.7769 | 0.0014 | 0.0013 |
| Cerebellum_7b_right | 3.7865 | 4.4548 | 0.0031 | 0.0002 |
| Cerebellum_8_left | 4.5323 | 4.0236 | 0.0008 | 0.0005 |
| Cerebellum_8_right | 4.1548 | 4.0764 | 0.0019 | 0.0004 |
| Cerebellum_9_left | 4.0987 | 1.1957 | 0.0001 | 0.1625 |
| Cerebellum_9_right | 3.3274 | 1.0976 | 0.0033 | 0.1802 |
| Cerebellum_10_left | 4.272 | 0.3853 | 0.0005 | 0.3904 |
| Cerebellum_10_right | 3.0964 | 0.1172 | 0.0069 | 0.4721 |
| Vermis_1_2 | 0.4998 | 1.2171 | 0.3604 | 0.1553 |
| Vermis_3 | 1.9555 | 2.6379 | 0.0574 | 0.0156 |
| Vermis_4_5 | 2.3617 | 2.5523 | 0.0215 | 0.0164 |
| Vermis_6 | 2.7539 | 3.9246 | 0.0125 | 0.0003 |
| Vermis_7 | 3.4607 | 2.2323 | 0.0019 | 0.0342 |
| Vermis_8 | 3.6813 | 2.1107 | 0.0025 | 0.0417 |
| Vermis_9 | 3.6405 | 3.6941 | 0.003 | 0.0002 |
| Vermis_10 | 1.6358 | 1.9943 | 0.0867 | 0.051 |

**Supplementary Material 6k.**
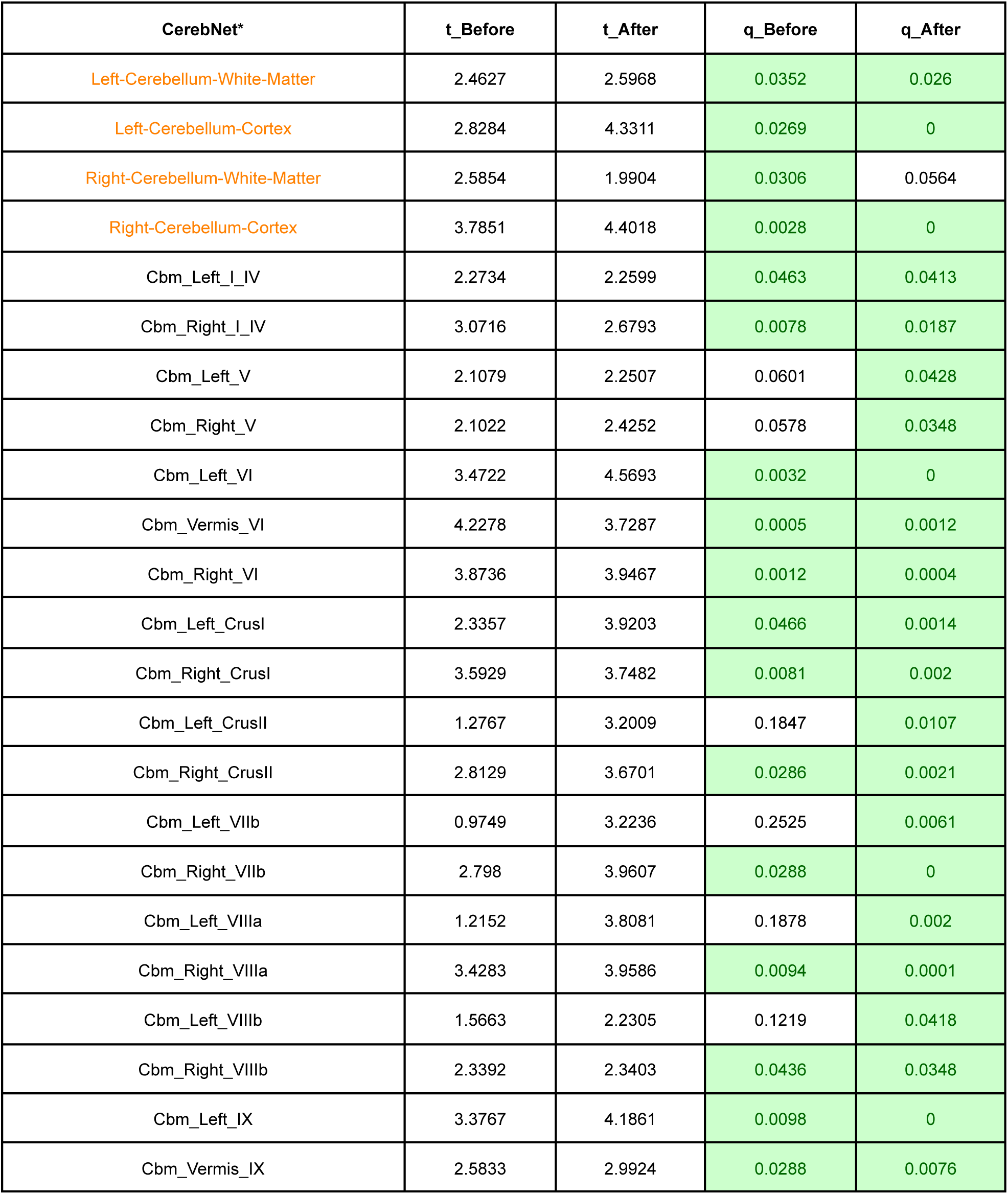
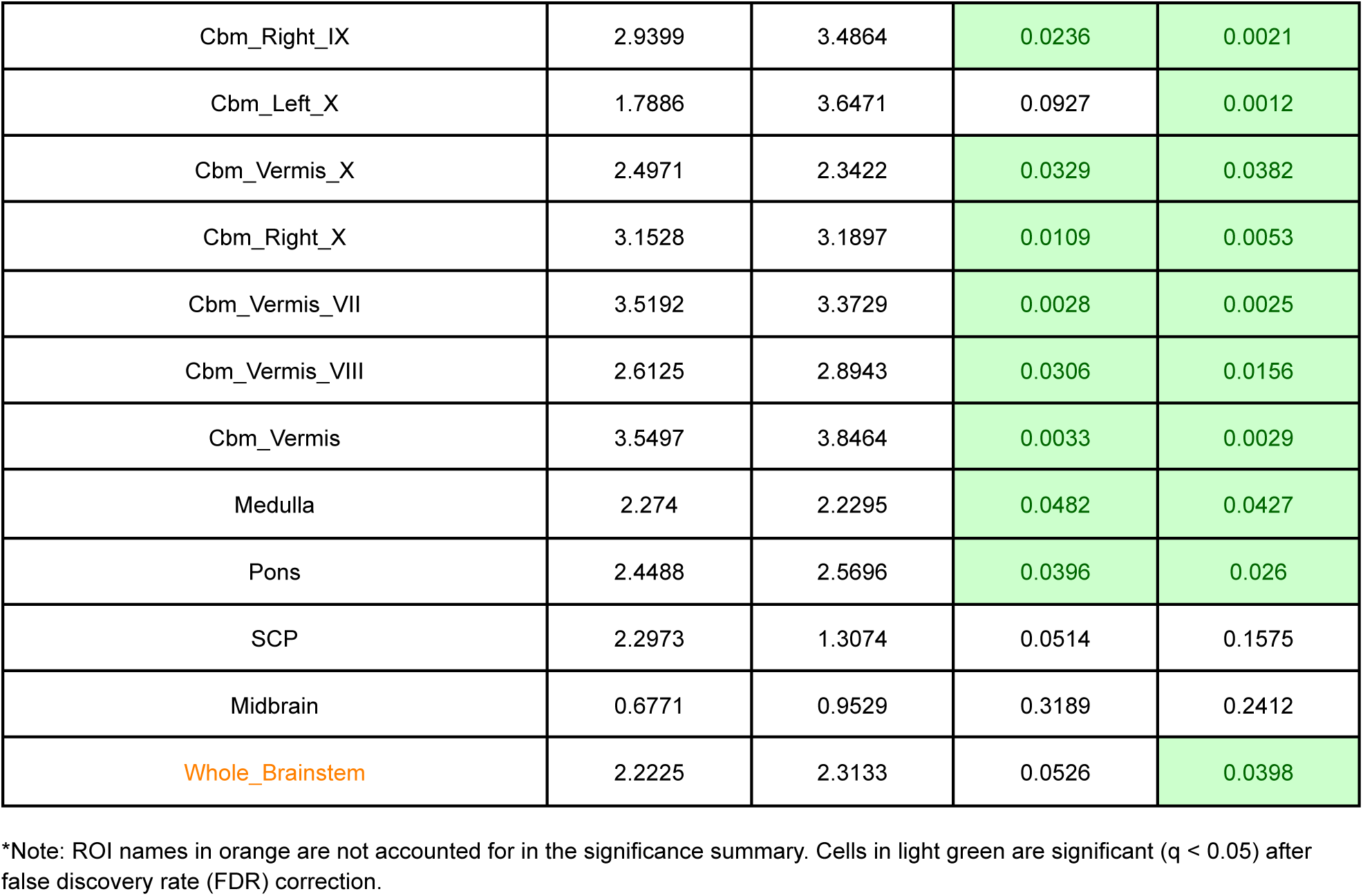

| CerebNet* | t_Before | t_After | q_Before | q_After |
| --- | --- | --- | --- | --- |
| Left-Cerebellum-White-Matter | 2.4627 | 2.5968 | 0.0352 | 0.026 |
| Left-Cerebellum-Cortex | 2.8284 | 4.3311 | 0.0269 | 0 |
| Right-Cerebellum-White-Matter | 2.5854 | 1.9904 | 0.0306 | 0.0564 |
| Right-Cerebellum-Cortex | 3.7851 | 4.4018 | 0.0028 | 0 |
| Cbm_Left_I_IV | 2.2734 | 2.2599 | 0.0463 | 0.0413 |
| Cbm_Right_I_IV | 3.0716 | 2.6793 | 0.0078 | 0.0187 |
| Cbm_Left_V | 2.1079 | 2.2507 | 0.0601 | 0.0428 |
| Cbm_Right_V | 2.1022 | 2.4252 | 0.0578 | 0.0348 |
| Cbm_Left_VI | 3.4722 | 4.5693 | 0.0032 | 0 |
| Cbm_Vermis_VI | 4.2278 | 3.7287 | 0.0005 | 0.0012 |
| Cbm_Right_VI | 3.8736 | 3.9467 | 0.0012 | 0.0004 |
| Cbm_Left_CrusI | 2.3357 | 3.9203 | 0.0466 | 0.0014 |
| Cbm_Right_CrusI | 3.5929 | 3.7482 | 0.0081 | 0.002 |
| Cbm_Left_CrusII | 1.2767 | 3.2009 | 0.1847 | 0.0107 |
| Cbm_Right_CrusII | 2.8129 | 3.6701 | 0.0286 | 0.0021 |
| Cbm_Left_VIIb | 0.9749 | 3.2236 | 0.2525 | 0.0061 |
| Cbm_Right_VIIb | 2.798 | 3.9607 | 0.0288 | 0 |
| Cbm_Left_VIIIa | 1.2152 | 3.8081 | 0.1878 | 0.002 |
| Cbm_Right_VIIIa | 3.4283 | 3.9586 | 0.0094 | 0.0001 |
| Cbm_Left_VIIIb | 1.5663 | 2.2305 | 0.1219 | 0.0418 |
| Cbm_Right_VIIIb | 2.3392 | 2.3403 | 0.0436 | 0.0348 |
| Cbm_Left_IX | 3.3767 | 4.1861 | 0.0098 | 0 |
| Cbm_Vermis_IX | 2.5833 | 2.9924 | 0.0288 | 0.0076 |
| Cbm_Right_IX | 2.9399 | 3.4864 | 0.0236 | 0.0021 |
| Cbm_Left_X | 1.7886 | 3.6471 | 0.0927 | 0.0012 |
| Cbm_Vermis_X | 2.4971 | 2.3422 | 0.0329 | 0.0382 |
| Cbm_Right_X | 3.1528 | 3.1897 | 0.0109 | 0.0053 |
| Cbm_Vermis_VII | 3.5192 | 3.3729 | 0.0028 | 0.0025 |
| Cbm_Vermis_VIII | 2.6125 | 2.8943 | 0.0306 | 0.0156 |
| Cbm_Vermis | 3.5497 | 3.8464 | 0.0033 | 0.0029 |
| Medulla | 2.274 | 2.2295 | 0.0482 | 0.0427 |
| Pons | 2.4488 | 2.5696 | 0.0396 | 0.026 |
| SCP | 2.2973 | 1.3074 | 0.0514 | 0.1575 |
| Midbrain | 0.6771 | 0.9529 | 0.3189 | 0.2412 |
| Whole_Brainstem | 2.2225 | 2.3133 | 0.0526 | 0.0398 |

\*Note: ROI names in orange are not accounted for in the significance summary. Cells in light green are significant ( $q < 0.05$ ) after false discovery rate (FDR) correction.

### Supplementary Material 7

#### List of abbreviations

AAL3: Automated Anatomical Labeling Atlas 3
AFNI: Analysis of Functional NeuroImages
AMHEN: Arithmetic Mean of Hemispheric Euler Numbers
ANTsX: Advanced Normalization Tools Ecosystem
BIDS: Brain Imaging Data Structure
CAT: Computational Anatomy Toolbox
CNNs: Convolutional Neural Networks
CSF: Cerebrospinal Fluid
DICOM: Digital Imaging and Communications in Medicine
DK40: Desikan-Killiany Atlas
FAST: FMRIB Automated Segmentation Tool
FMRIB: Functional Magnetic Resonance Imaging of the Brain
GM: Gray Matter
INU: Intensity Non-Uniformity
ITK: Insight Segmentation and Registration Toolkit
MP2RAGE: Magnetization Prepared 2 Rapid Acquisition Gradient Echoes
MPRAGE: Magnetization Prepared Rapid Acquisition Gradient Echo
MRI: Magnetic Resonance Imaging
MRIQC: Magnetic Resonance Image Quality ControlQuality control (QC)
N3: Non-Parametric Non-Uniform Intensity Normalization
N4ITK: Nick’s Non-Parametric Non-Uniform Intensity Normalization for ITK
NIfTI: Neuroimaging Informatics Technology Initiative
NON: Normative Outlier Number
QC: Quality control
SA2RAGE: Saturation-Prepared with 2 Rapid Gradient Echoes
SIQR: Structural Image Quality Rating
SPM: Statistical Parametric Mapping
T1w: T1-weighted
TIV: Total Intracranial Volume Ventral
DC: Ventral Diencephalon
UHF: Ultra-High Field
VINN: Voxel-size Independent Neural Network
WM: White Matter

## Notes

### Competing Interest Statement

The authors have declared no competing interest.

### Author Declarations

The Medical Ethics Review Committee of the Maastricht University Medical Center gave ethical approval for this work.

